# Subnational tailoring of malaria interventions to prioritize the malaria response in Guinea

**DOI:** 10.1101/2024.06.26.24309532

**Authors:** Ousmane Oumou Diallo, Abdourahamane Diallo, Kok Ben Toh, Nouman Diakité, Mohamed Dioubaté, Manuela Runge, Tasmin Symons, Elhadj Marouf Diallo, Jaline Gerardin, Beatriz Galatas, Alioune Camara

**Author notes:** Denotes equal contribution (Ousmane Oumou Diallo, Abdourahamane Diallo, Kok Ben Toh).

## Abstract

**Background:** In the context of high malaria burden yet limited resources, Guinea’s national malaria program adopted an innovative subnational tailoring (SNT) approach, including engagement of stakeholders, data review, and data analytics, to update their malaria operational plan for 2024-2026 and identify the most appropriate interventions for each district considering the resources available.

**Methods:** Guinea’s malaria program triggered the SNT exercise with a list of decisions that could be informed with local data. The program established an SNT team, which determined intervention targeting criteria; identified, assembled, and reviewed relevant data sources; stratified malaria risk and its determinants to inform geographical targeting for each intervention; and used mathematical modeling to predict the impact of different intervention mix scenarios. The SNT analysis was performed at the district level, excluding the urban area of Conakry.

**Results:** Malaria incidence, malaria prevalence, and all-cause under-5 mortality were used for the epidemiological stratification of Guinea. Additional indicators relevant for decisions-making including seasonality patterns, insecticide resistance or malaria interventions and vaccine coverage were also stratified. Stratified layers were used to inform the targeting criteria for each intervention to identify districts to prioritize for indoor residual spray, dual-action insecticide-treated nets, seasonal malaria chemoprevention (SMC) including number of cycles for each eligible district, malaria vaccine, and perennial malaria chemoprevention. Results of the SNT analysis were used to mobilize funding from the Global Fund for scale-up of dual-action nets and expansion of SMC.

**Conclusions:** SNT allowed Guinea’s national malaria program to adapt their intervention strategy at the health district level, an unprecedented approach in the country. The use of local data to inform eligibility and prioritization allowed the program to identify the optimal mix of interventions for each district and to successfully mobilize resources to support their plans.

## Background

Malaria is the primary cause of outpatient visits and hospitalizations in children under five years of age in Guinea despite years of intense malaria control efforts (1,2). Guinea’s national malaria program, the Programme National de Lutte contre le Paludisme (PNLP), developed a national strategic plan for malaria covering the period 2023 to 2027, which includes the deployment of pyrethroid-based insecticide-treated bednets (ITNs), seasonal malaria chemoprevention (SMC), intermittent preventive treatment in pregnancy (IPTp), introduction of a malaria vaccine, and treatment of symptomatic cases. The national strategy development process also considered second-generation ITNs, indoor residual spraying (IRS), and perennial malaria chemoprevention (PMC, then known as intermittent preventive treatment for infants) as interventions of interest should additional resources become available.

Main funders for the malaria response in Guinea include the Global Fund against AIDS, Tuberculosis and Malaria (50%), the US President’s Malaria Initiative (PMI) (40%), with World Bank (3%), and domestic sources (7%). As is the case globally (3), the total resources available for malaria response in Guinea are insufficient to fund all interventions necessary to achieve key targets.

In 2023, the PNLP underwent a thorough prioritization exercise to review the malaria control plan for 2024 to 2026, aligned with the Global Fund funding cycle (GC7). This exercise aimed to maximize the impact of the resources available for the fight against malaria in Guinea, following the subnational tailoring of interventions (SNT) approach recommended by the World Health Organization (WHO) (3,4).

In brief, SNT consists of using local data and additional contextual information available to malaria control programs to inform decision-making. In the context of strategic planning and prioritization, SNT is used to determine the most appropriate mix of interventions and strategies for a given area. Through SNT, intervention plans are defined that achieve optimum impact on malaria burden either resource-agnostically in a strategic plan, or within a specific resource envelope. SNT can also be used to inform how new tools (such as malaria vaccines) can be integrated most effectively within existing plans, or for dynamic review of plans as additional funding opportunities become available (3).

Guinea’s PNLP implemented SNT to respond to six questions related to the prioritization and implementation of five prevention interventions contemplated in their strategy, within the available domestic and international resource envelope, (Figure 1, Table 1). The questions were: which districts to prioritize for more effective vector control (1) IRS or 2) dual-action Interceptor G2 (IG2) bednets); 3a) which districts to target with SMC, and 3b) how many cycles of SMC should targeted districts receive; 4) which districts to prioritize for the malaria vaccine RTS,S/AS01 (RTS,S) given the limited supply of doses available in 2023; and 5) which districts to prioritize for PMC. An additional question was raised regarding the tailoring of the malaria response in the capital city Conakry, where the malaria transmission has significantly decreased and become increasingly more heterogeneous and focalized. Given the differential approaches and units of analysis required to address the first six questions and the last question, the SNT team split the exercise into two. This manuscript presents the activities and results obtained from the prioritization of interventions in areas outside of Conakry.

**Figure 1:**
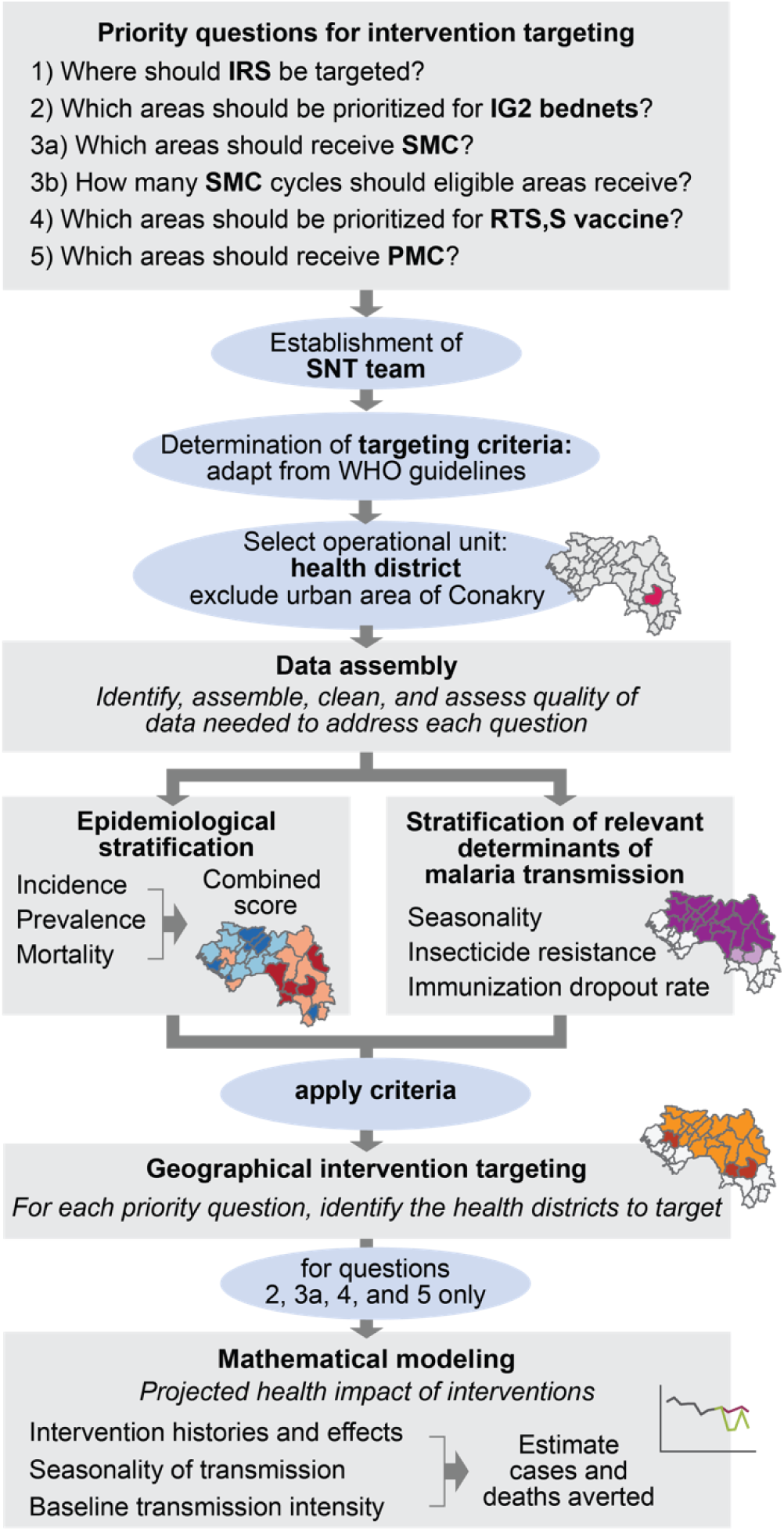
Overview of the SNT intervention prioritization and modeling process. The Guinea PNLP identified priority questions to inform the prioritization of several interventions within their strategy. An SNT team was established and targeting criteria determined. Data were assembled for each health district. Districts were stratified according to malaria risk and relevant determinants for decision-making. Targeting criteria were applied to identify priority districts per intervention. Mathematical modeling was used to provide further evidence for a subset of questions.

**Table 1.** Priority questions for intervention targeting in Guinea and data needs for addressing each question.

| Question | Indicator | All-age malaria clinical case incidence | Malaria infection prevalence estimates in children under 5 | Estimates of all-cause mortality in children under 5 | Case and rainfall seasonality | Entomological and insecticide resistance indicators | Immunization coverage | Intervention history |
| --- | --- | --- | --- | --- | --- | --- | --- | --- |
|  | Data needs | Routine, demographic, and care-seeking behavior information | Household surveys and geospatial modeled estimates | Household surveys and geospatial modeled estimates | Monthly routine case data and rainfall data | Entomological data and vector mortality after 24h of exposure to pyrethroids and other insecticides | Routine EPI data | Historical intervention implementation data |
| 1: Where should IRS be targeted? |  | X | X | X |  | X |  | X |
| 2: Which areas should be prioritized with IG2 nets? |  | X | X | X |  | X |  | X |
| 3a: Which areas should receive SMC? |  | X | X |  | X |  |  | X |
| 3b: How many SMC cycles should eligible areas receive? |  | X | X |  | X |  |  | X |
| 4: Which areas should be prioritized for RTS,S vaccine? |  | X | X | X |  |  | X |  |
| 5: Which areas should receive PMC, and where should it be piloted? |  | X | X |  | X |  | X |  |

To address these questions, the SNT process was implemented by the PNLP through the following steps (Figure 1):

1) **Establishment of an SNT team.** The PNLP created and led a local SNT team under direction of the PNLP program manager. The SNT team was responsible for the oversight of the entire SNT exercise, including ensuring that consensus was reached by all relevant stakeholders at each step of the process and all actors were ultimately aligned under a single, collectively-discussed plan. The team included members from the PNLP, WHO, Northwestern University, the Global Fund, PMI, Catholic Relief Services, and RTI-Notre Santé. The PNLP regularly convened the SNT team until completion of the exercise.
2) **For each intervention under consideration, determination of criteria to identify areas to target or prioritize.** The first task of the SNT team was to determine the criteria that would inform the targeting of each of the interventions under consideration by the PNLP and to determine the operational unit for SNT. The SNT team took WHO’s available guidance and practical information for the implementation of each intervention during the period of this exercise (5) as the basis for the criteria, and adapted them to their specific context. These criteria then established the types of information and data required for review and analysis (Table 1). The SNT team identified the health district (Figure 2) as the lowest operationally feasible administrative unit at which interventions ought to be prioritized. Together, the criteria and the identification of the unit for SNT analysis established the data needs for the SNT exercise and the analytical outputs required to inform decisions.
3) **Data collection and stratification of indicators required for decision-making.** The SNT team identified and assembled all appropriate databases for analysis, then thoroughly assessed their quality. Next, the key indicators to inform about malaria transmission intensity patterns (clinical malaria incidence, prevalence, and mortality), and its determinants (seasonality patterns, insecticide resistance, coverage of implemented malaria interventions, immunization coverage, etc.) were estimated and stratified into relevant categories for decision-making.
4) **Geographic targeting of each intervention at subnational level.** Using the relevant stratified layers obtained in the previous step, the SNT team applied the specified criteria to identify the districts in most need for each intervention. This process led to the development of various scenarios of intervention mixes under different resource conditions.
5) **Mathematical modeling the impact of each intervention at subnational level.** Transmission models were used to predict the impact on future malaria transmission and burden of the different scenarios, for those questions where mathematical modeling was appropriate.

**Figure 2:**
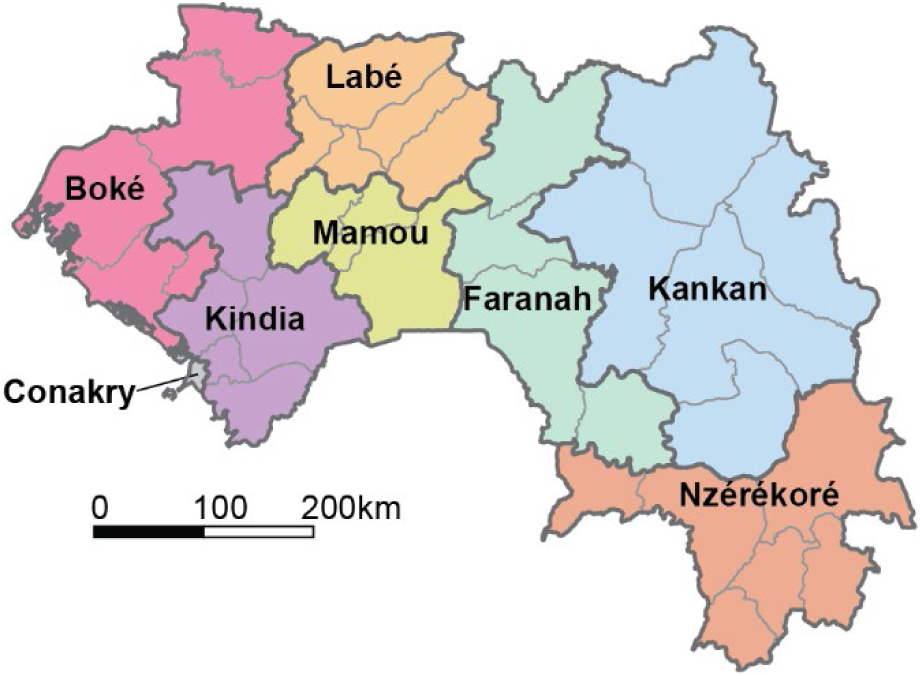
Regions (bold lines and colors) and health districts (thin lines) of Guinea. Conakry was excluded from all analyses presented here.

The resulting prioritized mix of interventions was incorporated into Guinea’s 2023 funding application to the Global Fund against AIDS, Tuberculosis, and Malaria, and was used to inform the activities considered under PMI’s annual Malaria Operational Plans (MoPs). The SNT exercise also identified knowledge gaps for the understanding of the malaria epidemic and its determinants in specific parts of the country and highlighted the need for improved surveillance, data structuring and review systems, and data quality.

## Methods

Thirty-three health districts were included in the SNT analysis to prioritize the malaria response in Guinea (Figure 2).

### Data assembly

Data needs for each priority question were identified (Table 1), and data were assembled from various sources within and outside the PNLP to estimate malaria risk and additional information of interest.

Routine surveillance data were obtained from the District Health Information System 2 (DHIS2) of Guinea for the period 2018 to 2022 to estimate clinical malaria incidence and to inform case seasonality patterns. Indicators extracted per health facility included: all-cause outpatient visits, suspected malaria cases, tested malaria cases, confirmed malaria cases, treated malaria cases, all-cause hospitalizations, malaria hospitalizations, treated severe malaria cases, all-cause deaths, and malaria deaths. In total 694 health facilities that received outpatient consultations and hospitalizations, (563 public and 131 private), were included in this analysis. Health facility level data were reviewed for quality and completeness (see Supplementary File 1) and aggregated to the health district level for analysis. Population estimates per health district were obtained from the Institut National de la Statistique of Guinea (6).

Regional-level estimates of malaria infection prevalence in children under the age of five (U5) were obtained from the 2012 and 2018 Demographic Health Surveys (DHS) or 2021 Malaria Indicator Surveys (MIS), hereafter referred to collectively as DHS (7). The DHS of 2012 and 2018 also provided regional-level estimates of all-cause under-five deaths at the community-level. Both of these indicators were used to estimate annual prevalence and all-cause mortality estimates at the district-level.

Monthly rainfall data to inform seasonality patterns per district were obtained from ERA5 (8) for the period between January 2015 to December 2021, using the coordinates of the district centroid. Entomological data on pyrethroid resistance in 2022 were obtained from entomological sentinel surveillance sites throughout Guinea (9,10).

Historical intervention implementation information available to the PNLP included: for SMC, history of SMC implementation per district from 2018 to 2022, including number and timing of SMC rounds; for ITNs, number distributed in the three most recent mass distributions of 2016, 2019 and 2022, number of nets distributed routinely, and type of nets distributed; for immunizations in 2021, number of children vaccinated for each of the three Diphtheria-Tetanus-Pertussis (DTP) doses and for the first measles dose. Data on treatment rate of fever and access to and use of mosquito nets were extracted from the DHS or MIS of 2012, 2018, and 2021 (11–13). Care-seeking behavior rates for a fever were calculated per sector of care (public or private) as the proportion of children U5 who had fever within the two weeks prior to the survey and sought care for the fever. Use of mosquito nets was calculated as the proportion of people who slept under a bednet on the night before the survey.

### Epidemiological stratification

#### Clinical malaria incidence estimates

To estimate community-level clinical malaria incidence and thereby measure malaria transmission per health district and year, four incidence estimates were considered following a standard approach used by the WHO in the World Malaria Report (3): This approach adjusts crude incidence by the main factors that affect the final number of monthly confirmed cases reported per health facility in an additive fashion. The first adjustment aims to correct for varying testing rates of suspected cases. The resulting number of cases is further adjusted by the lack of reporting of data to DHIS2 for some health facilities and months. The final adjustment considers varying levels of care seeking of fevers outside of the public health sector (14). All adjustments were made separately for each health district between 2018 and 2022 and presented to the SNT team for review. See Supplementary File 1 for full methods on incidence adjustments.

Maps per incidence indicator were produced per year and reviewed by the SNT team with the aim of identifying the most appropriate clinical malaria incidence metric and period of time for decision-making. Upon review of the assumptions and limitations of each estimate, a decision was made to use the median clinical malaria incidence adjusted for testing and reporting rates (second adjustment level) between 2018 and 2022. This was decided due to the geographical and temporal resemblance of the resulting estimates to the local understanding of transmission patterns after expert review.

#### *Plasmodium* falciparum infection prevalence estimates in children under 5

Estimates of *Plasmodium falciparum* parasite rate (*Pf*PR) at health-district aggregation were provided by the Malaria Atlas Project (MAP). These estimates were generated using a geospatial model taking as response data the routine case data described above, augmented by geo-located cross-sectional data on malaria parasitaemia by RDT, non-malarial fever incidence in the two weeks prior to survey, and care-seeking for fever from MIS 2021 (15). After adjusting the routine data for missingness and care-seeking in steps logically consistent with the approach outlined above, monthly estimates of *Pf*PR January 2015-December 2021 were inferred by reconciling the cross-sectional observations of RDT-positivity in children with the adjusted health-district-aggregated case counts via a semi-mechanistic Bayesian model accounting for (a) varying environmental and socio-economic receptivity using high-resolution remotely-sensed covariates; (b) the non-linear prevalence-incidence relationship; and (c) the role of non-malarial febrile illness as a driver of care-seeking for incidental fevers. This joint inference procedure ensures biological consistency between estimates of *Pf*PR and clinical incidence, in addition to informing the interpolation of *Pf*PR trends between surveys with routine case data.

#### All-cause under-five mortality rate

Estimates of the all-cause under-5 mortality rate (U5MR) by health district were obtained from the Institute for Health Metrics and Evaluation (IHME) for the period 2000-2017 using a model that draws on birth history information collected in household surveys, census data, and other various mortality risk factors (16). Ideally, estimates of malaria mortality would have been more appropriate for the stratification process. However, the precise estimation of malaria mortality in the community is difficult and uncertain due to under-reporting of deaths and low sensitivity of the public health and surveillance system to capture the malaria deaths that occur in the community. For this analysis, the distribution and relative intensity of mortality are more important than the magnitude. Therefore, for the purpose of this exercise, the SNT team assumed that areas with higher relative U5MR rates coincided with those with high malaria mortality rates.

#### Risk categorization and combining indicators

A joint indicator of *Pf*PR, clinical malaria incidence adjusted for testing and reporting rates, and U5MR was used to obtain an estimate of overall malaria risk. This approach was carried out in two steps.

First, scores of 1 to 4 were assigned to the relevant categories of prevalence (<10%, 10-20%, 20-40% or >40%) and incidence (<100, 100-250, 250-450 or >450 cases per 1000 population at risk).

The sum of the prevalence and incidence scores were reclassified into four groups on the basis of the combined scores, ranked from "lowest" (score of 4) to "highest" (score of 7).

Second, the morbidity indicator categories were assigned to the scores from 1 (low) to 4 (high); and combined with the scored categories of U5MR (<9.5, 9-5-12.5, 12.5-15, or >15 deaths per 1000 live births). Health districts were reclassified into four groups on the basis of the combined morbidity and mortality scores, ranked from "lowest" (score of 6) to "highest" (score of 9). The resulting maps were reviewed and discussed with all SNT team members.

### Analysis of other indicators relevant for malaria decision-making

#### SMC eligibility and timing

Malaria case trends are generally seasonal and strongly dependent on rainfall, and SMC is recommended by WHO in areas of seasonal and moderate to high transmission (17). Ideally, identification of areas with short, intense seasonality suitable for SMC would be done through analysis of seasonality patterns in the number of monthly or weekly cases reported. The preliminary analysis of seasonality of cases revealed ambiguous patterns of seasonality that did not align with the country’s reality. The SNT team agreed that routine data was biased by factors such as care-seeking behavior, reporting rates, and the impact of SMC and other interventions implemented right before the rainy season. Therefore, rainfall trends were used instead to evaluate suitability of seasonality for SMC in each health district. Districts with *Pf*PR of 5% or higher were considered moderate to high transmission, which included all districts in the country.

A health district was considered seasonal when at least 60% of cases (or rainfall) occur during four months of the year. To evaluate which health districts met this definition, the same algorithm was applied to monthly confirmed malaria cases and monthly rainfall data from CHIRPS (18). For each indicator, a sliding window of four months was applied to each consecutive 4-month block of data. Seasonality peaks were defined as those 4-month blocks where the sum of cases (or rainfall) within the block was at least 60% of the sum of cases (or rainfall) in a 12-month block beginning with the same month. A district was considered seasonal if at least 50% of evaluated years contained one or more consecutive seasonal month-blocks. Results of the case and rainfall evaluations for each district were reviewed by the PNLP prior to final determination of SMC eligibility.

Given that rainfall was used as a proxy to identify malaria case peaks, the trends in rainfall and cases were evaluated among districts with >5% prevalence with suitable seasonality patterns for analysis. Districts with clear disparities between cases and rainfall trends were discussed to identify additional determinants of malaria trends, such as agricultural practices or presence of other environmental factors, that could explain said disparities. SMC eligible districts according to rainfall trends with additional determinants that contribute to longer or no seasonal trends in cases were considered ineligible for SMC. Finally, districts with previous history of SMC implementation were also considered eligible for SMC.

In Guinea, SMC is targeted to children U5, and monthly routine data on confirmed malaria cases in children U5 were used to determine the timing of SMC cycles for eligible districts. The beginning of the seasonal peak was defined as the month with an increase in cases of over 60% compared with the previous month. The proportion of cases that would be covered by four cycles, i.e. the fraction of annual cases that occur during months with SMC cycles, with SMC beginning in either June or July, was evaluated for all SMC-eligible districts.

Given a 4-cycle schedule beginning in July, two options were explored for the month in which to introduce a 5th cycle in districts where the rainfall analysis found a peak of five months. Ideally, the 5th cycle would be placed in the month before (June) or after (November) the July-October period that would maximize the number of cases averted. Thus, for each district eligible for five cycles, the proportion of cases occurring in June and November was assessed.

#### Immunization dropout rate

Number of children receiving the first, second and third doses of DTP vaccine and the first dose of measles vaccine was obtained for each district for 2021 using routine data from DHIS2. To identify districts with strong immunization implementation where the malaria vaccine or PMC could be successfully implemented, dropout rates were calculated for DTP1 to DTP2, DTP2 to DTP3, and DTP3 to measles. Each dropout rate was calculated by dividing the difference between the number of children receiving the later dose and earlier dose by the number of children receiving the earlier dose. After review, the DTP1 to DTP2 and DTP2 to DTP3 dropout rates were determined to have greater data quality and were considered when determining priority districts for malaria vaccine.

### Intervention targeting

The criteria for each intervention, adapted from WHO guidelines, were then applied to Guinea’s health districts according to the stratifications of epidemiology and malaria determinants described above to generate intervention mix maps relevant for each priority question.

### Mathematical modeling for impact prediction

Impact predictions were made with EMOD v2.20, an agent-based mathematical model of malaria transmission (19). The general approach used for adapting EMOD to Guinea was based on an approach used for Nigeria (20). Full details of the modeling methods are available in Supplementary file 2.

EMOD was parameterized separately for each of the 33 health districts considered for intervention targeting, using district-level data where possible. A four-step process was used for the modeling. First, past and present interventions in each district were added, including each intervention’s schedule, coverage, and effect size. Second, the modeled case seasonality was matched to confirmed case seasonality in the routine data. Third, the modeled baseline transmission intensity in each district was fit to malaria prevalence and incidence data. These three steps are described in detail in Supplementary File 2. Last, impact predictions of intervention scenarios under consideration by the PNLP were modeled for the period 2023-2027 for uncomplicated malaria cases and malaria deaths in children U5 and in individuals of all ages.

The baseline scenario to which other possible intervention mixes were compared was selected by the PNLP as the intervention mix that was, in their opinion, likely to be funded. This base mix assumed that case management, SMC, and ITN usage would remain at 2020 levels. Districts designated as Group 1 priority (see Results) received IG2 nets, whereas remaining districts received standard pyrethroid nets. SMC was implemented only in districts that were already receiving SMC in 2022, and no PMC or vaccine was implemented in any district.

For a subset of the questions of interest to the PNLP, the base intervention mix was compared to an alternate mix as follows:

- For Question 2: Compare base intervention mix to 1) base mix with pyrethroid nets in all districts and 2) base mix with IG2 nets in all districts.
- For Question 3a: Compare base intervention mix to base mix with SMC implementation in additional districts.
- For Question 4: Compare base intervention mix to base mix with PMC implemented in selected districts, with or without RTS,S vaccine distribution in Yomou district.

For each district, malaria transmission was simulated from 1960 to 2022, including all historical interventions as described earlier, for 15 stochastic realizations of each of the 20 selected parameter sets, a total of 300 model runs per district. For each intervention scenario under consideration, these 300 model runs were continued from 2023 to 2027.

Annual clinical incidence and deaths, for all ages and for children U5, were extracted for each simulation. Modeled incidence included both treated and untreated cases. A set of 1000 national-level incidence and death rates were generated from the 300 model runs of each district with a weighted average accounting for the district population (see Supplementary file 2). Mean and 95% predicted intervals were calculated from the 1000 national estimates for each scenario.

## Results

Results are presented for the data quality assessment, epidemiological stratification of Guinea, and intervention targeting plans developed in response to each of the priority questions. Impact predictions from mathematical models are shown for scenarios of interest to the PNLP.

### Data quality assessment

The reporting rate was greater than or equal to 80% in most health districts and improved between 2018 and 2022 (Figure S1.1 in Supplementary File 1). The proportion of missing data for total confirmed cases, suspected cases, tested cases, and treated cases was very low (Figure S1.2 in Supplementary File 1). The internal consistency check found that in almost all health facilities, the number of suspected cases was equal to the number of tested cases and the number of confirmed cases was lower than the number of tested cases (Figure S1.4 in Supplementary File 1), indicating good coherency across these key indicators. However, other comparisons, such as the number of treated cases and the number of confirmed cases, suggested instances of presumptive treatment, which was highlighted as an area for case management improvement.

### Epidemiological stratification

Crude and adjusted incidence per health district are shown in Figure 3A for the year 2022 (see Supplementary file 1 for other years). Crude incidence was between 100 and 300 cases per thousand per year for most health districts and greater than 300 for only two health districts.

**Figure 3:**
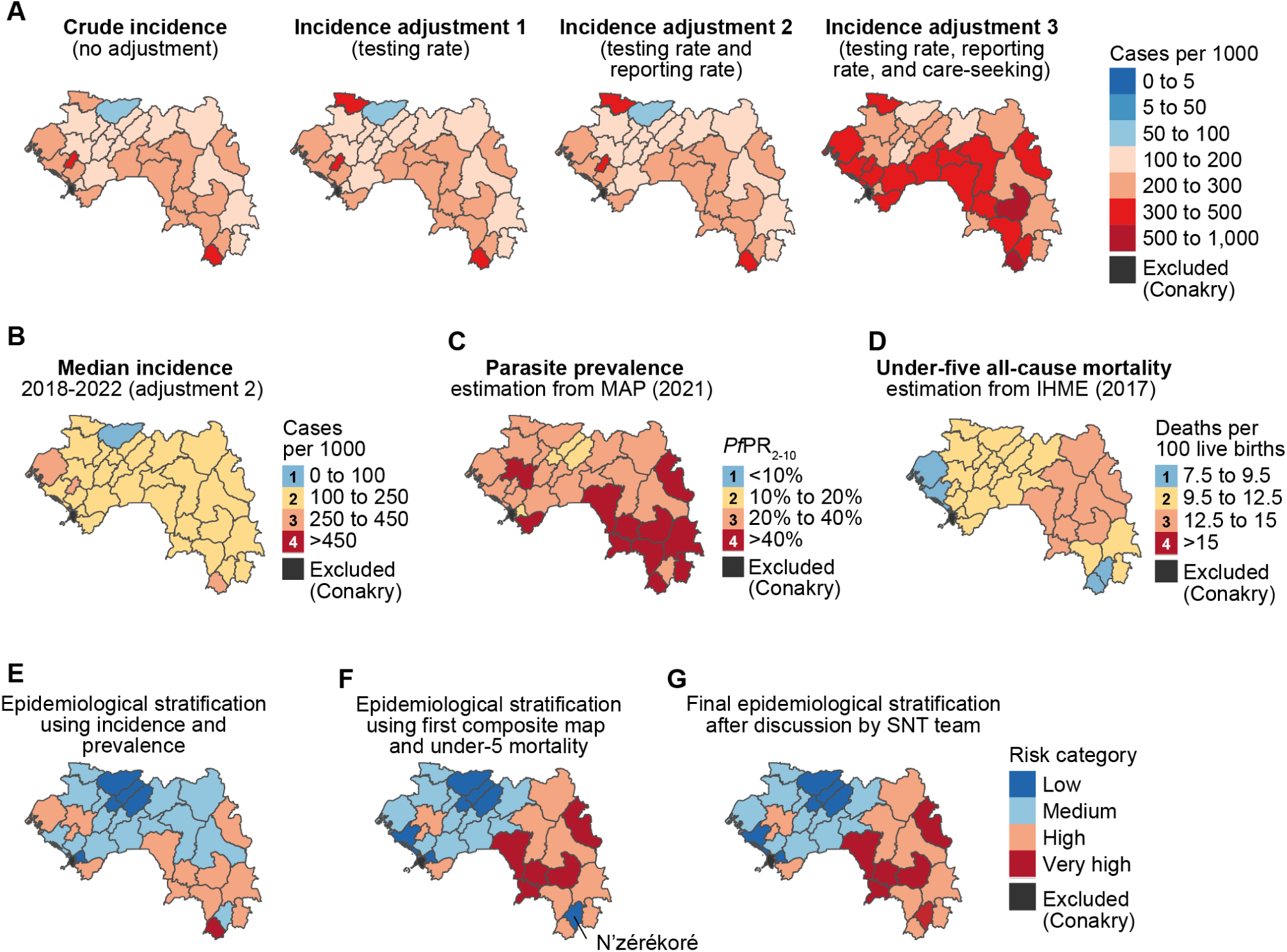
Malaria epidemiological stratification of Guinea to inform targeting of interventions. (A) The four estimated incidences using routine data for 2022. (B) Median incidence between 2018 and 2022 adjusted for testing and reporting rates, categorized into 4 strata. (C) Parasite prevalence in children under five estimated by MAP for the year 2021, categorized into 4 strata. (D) All-cause mortality in children under five estimated by IHME for the year 2017, categorized into 4 strata. (E) Malaria morbidity stratification from combining the maps in B and C. (F) Malaria morbidity and mortality stratification from combining the maps in E and D. (G) Final epidemiological stratification after review by the SNT team.

Adjusting for testing and reporting rates did not have a large impact on estimated incidence (Figure 3A and Figure S1.7 in Supplementary file 1), as testing and reporting rates were high in most health districts (Figures S1.1 and S1.10 in Supplementary file 1). Adjusting for care-seeking increased the estimated incidence to over 300 cases per thousand per year in 16 out of 33 health districts, as care-seeking in the public sector was reported at 39.6-69.4% in the 2021 MIS, depending on the region (Figure S1.5 in Supplementary file 1). Care-seeking in the public sector increased between the 2018 DHS and 2021 MIS, and therefore the magnitude of the adjustment for care-seeking was larger for earlier years.

After review of the four incidence estimates, the PNLP made the choice to use the median incidence adjusted for testing and reporting rates when stratifying health districts by level of malaria risk (Figure 3B). This choice was driven by the quality of routine data and concerns about the accuracy and district-level representability of the region-level care-seeking rates from the surveys. The selected median adjusted incidence was classified into three categories of transmission intensity following WHO definitions (21): one health district was very low malaria transmission (less than 100 cases per 1000 per year); 29 health districts were low malaria transmission (between 100 cases and 250 cases per 1000 per year); and three health districts were moderate malaria transmission (between 250 cases and 450 cases per 1000 per year).

In 2021, the district-level infection prevalence estimates in children under five was over 40% (high transmission) in 11 health districts, concentrated in the regions of N’zérékoré, Kankan, and Faranah (Figure 3C). Prevalence was <10% (low transmission) in only four health districts, three of which were in the region of Labé. All-cause under-five mortality estimates were very high (over 12.5 per 100 live births) in 8 districts in the regions of N’zérékoré, Kankan, and Faranah.

Figure 3E shows a composite map of malaria risk based on morbidity indicators (median adjusted incidence and prevalence in under-fives). In this stratification, five health districts, mainly in the Labé region, were classified in the lowest transmission stratum. Only the district of Yomou, located in the N’zérékoré region, was classified in the very high risk stratum.

The morbidity composite map (Figure 3E) was combined with stratified mortality (Figure 3D) to produce a composite risk stratification based on both morbidity and mortality (Figure 3F). Seven health districts were classified in the lowest risk stratum, in Labé, Boké, and Kindia regions. Districts in the low and medium strata were mostly in central or western Guinea, with the exception of N’zérékoré district in southeast Guinea being classified as low risk. Other districts in eastern and southern Guinea were in the high or very high risk strata. Routine data in N’zérékoré district had previously been shown to be of low quality and not representative of actual malaria incidence in the region (22) Thus the SNT team decided that N’zérékoré district was more likely to belong to the very high transmission stratum as per their knowledge of the area (Figure 3G).

### Targeting of IRS

WHO recommends IRS in areas of high receptivity, defined as *Pf*PR of at least 1% in 2000; all districts in Guinea met this criterion. Since IRS is a costly intervention, the PNLP considered IRS only in districts with very high morbidity and mortality (Figure 4), where it would have the highest potential impact, and districts in the region of N’zérékoré. Ultimately the PNLP decided not to include IRS in the 2023 funding request, although should resources become available, these ten districts would be the ones prioritized.

**Figure 4:**
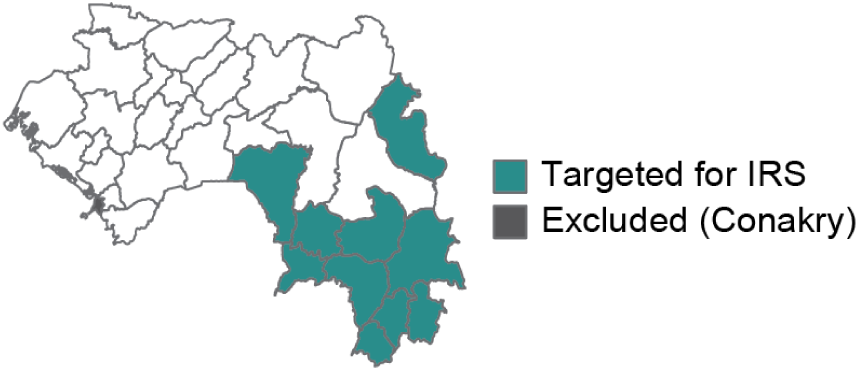
Target districts for IRS, if it were to be included in Guinea’s vector control strategy.

### Targeting and potential impact of IG2 bednets

Insecticide-treated nets distributed through mass campaigns and routine distribution are the main form of vector control implemented in Guinea. Entomological surveillance suggests that insecticide resistance is widespread across Guinea (9). Thus, all districts would benefit from the distribution of new-generation mosquito nets, particularly those with a dual-action insecticide, such as IG2 nets. In the context of limited resources, the PNLP prioritized IG2 distribution for districts with high or very high morbidity and mortality (Figure 3G), as well as the district of Forecariah, which had already received IG2 nets in the past as part of a pilot evaluation project. These districts were prioritized as Group 1 for IG2 nets, and the remaining districts formed Group 2 (Figure 5A).

**Figure 5:**
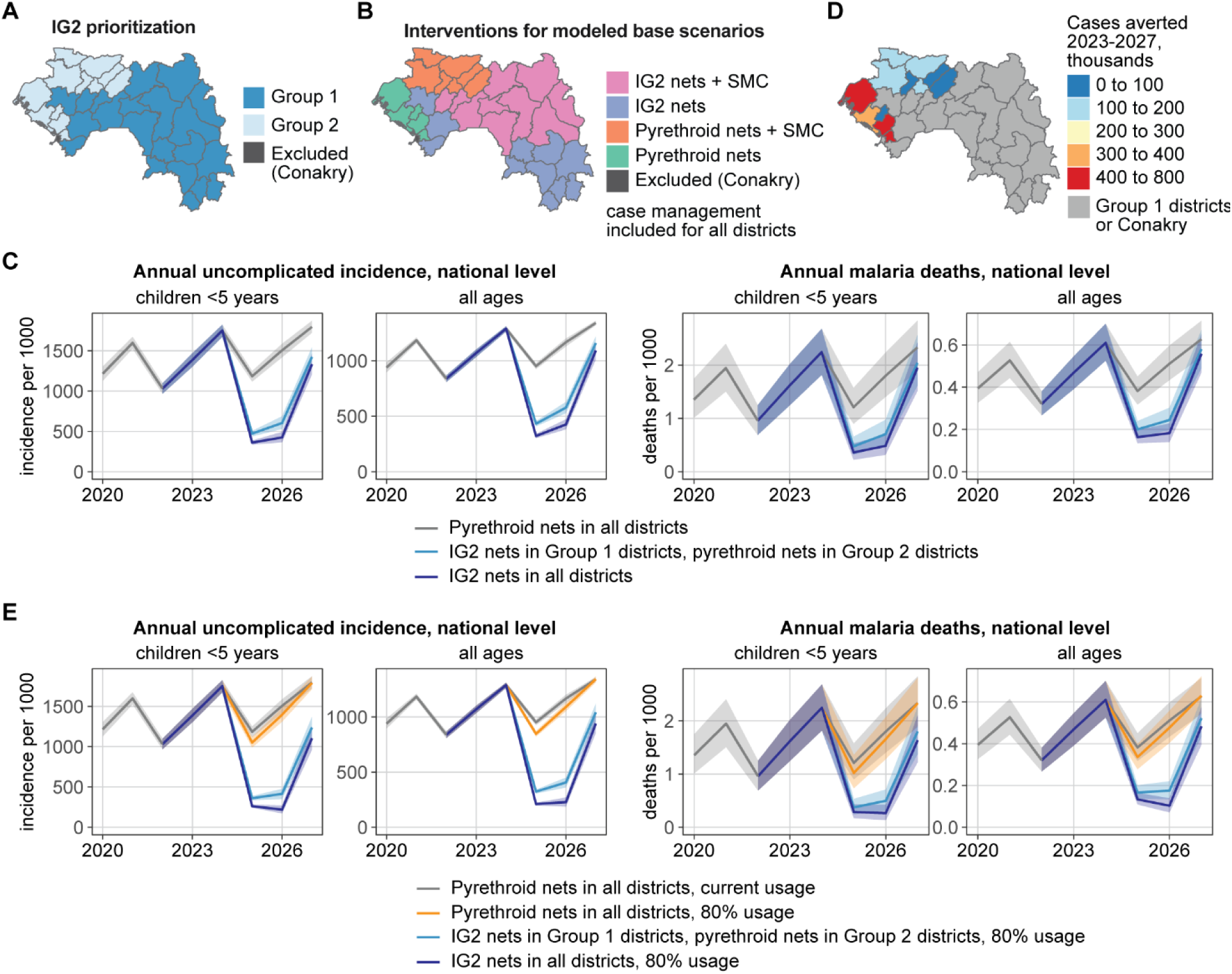
Prioritization of districts for IG2 implementation and predicted impact of IG2 bednets in Guinea. (A) Group 1 districts were prioritized for IG2 nets, and Group 2 districts would receive pyrethroid nets unless additional resources became available. (B) Base fundable package of interventions for modeled scenarios, to which additional interventions were added to assess impact. (C) Mean predicted incidence and death rate (solid line) and their 95% interval (shaded area) at the national level from 2020 to 2027 for IG2 in no districts, Group 1 districts only, and all districts. (D) Total cases averted from 2023 to 2027 in Group 2 districts if IG2 nets were implemented instead of pyrethroid nets. (E) Mean predicted incidence and death rate (solid line) and their 95% PI (shaded area) at the national level from 2020 to 2027 under various IG2 implementation and usage scenarios.

Districts in Group 2 for prioritization would receive pyrethroid nets unless additional resources were to become available for IG2 nets. The Group 1 targeting scheme for IG2 bednets, with SMC only in districts that have already been receiving SMC, formed the base scenario for mathematical modeling to which other intervention scenarios were compared (Figure 5B).

Distribution of IG2 nets in the mass campaign of 2025 was projected to reduce a substantial number of cases in all age groups (Figure 5C, Table 2), averting 22% of cases (95% PI: 21% to 24%) and 19% of deaths (95% PI: 14% to 25%) across all ages over the period 2023-2027 when distributed in Group 1 districts alone.

**Table 2:** Predicted impact of IG2 nets from 2023 to 2027, compared to the base scenario (Figure 4B) with pyrethroid nets in all districts, and ITN usage based on the 2021 MIS. The 95% prediction interval is indicated in parentheses.

|  | IG2 nets in Group 1 districts only | IG2 nets in all districts |
| --- | --- | --- |
| Children U5 years |  |  |
| Relative reduction in cases<br>Cases averted (millions) | 26% (25% – 28%)<br>3.5 (3.3 - 3.7) | 31% (29% – 33%)<br>4.1 (3.9 - 4.3) |
| Relative reduction in deaths<br>Deaths averted (thousands) | 23% (16% – 30%)<br>3.7 (2.5 - 5.0) | 28% (20% – 35%)<br>4.4 (3.1 - 5.9) |
| All ages |  |  |
| Relative reduction in cases<br>Cases averted (millions) | 22% (21% – 24%)<br>12.4 (11.6 - 13.1) | 28% (26% – 30%)<br>15.5 (14.6 - 16.4) |
| Relative reduction in deaths<br>Deaths averted (thousands) | 19% (14% – 25%)<br>5.0 (3.5 - 6.6) | 24% (19% – 29%)<br>6.2 (4.7 - 7.8) |

Implementation of IG2 nets also in Group 2 districts was simulated to further reduce the number of cases by 6% (95% PI: 5% to 6%) and the number of deaths by 5% (95% PI: 2% to 8%) in all ages, a smaller proportional reduction due to the lower malaria risk in Group 2 districts. Among individual districts in Group 2, the projected number of cases averted varied from a mean of 47 thousand to 630 thousand (Figure 5D) due to differences in population size, transmission intensity, and ITN usage. Districts in the regions of Boké and Kindia were predicted to have the most cases averted if IG2 were to be implemented.

Modeled ITN usage ranged from 42% to 80% (Figure S2.6 in Supplementary File 2). Under the implementation of pyrethroid nets in all districts, increasing net usage to at least 80% in all districts was predicted to avert 3% of cases (95% PI: 2% to 4%) in all ages (Figure 5E, Table 3). Under IG2 implementation in Group 1 districts, increasing usage from current to at least 80% was also beneficial: an additional 7% of cases (95% PI: 5% to 8%) and 6% of deaths (95% PI: 0.4% to 12%) averted compared with IG2 implementation under current usage. At the national level, a similar impact was predicted if IG2 was also implemented in Group 2 or if IG2 was limited to Group 1 but usage was increased to 80%.

**Table 3:**
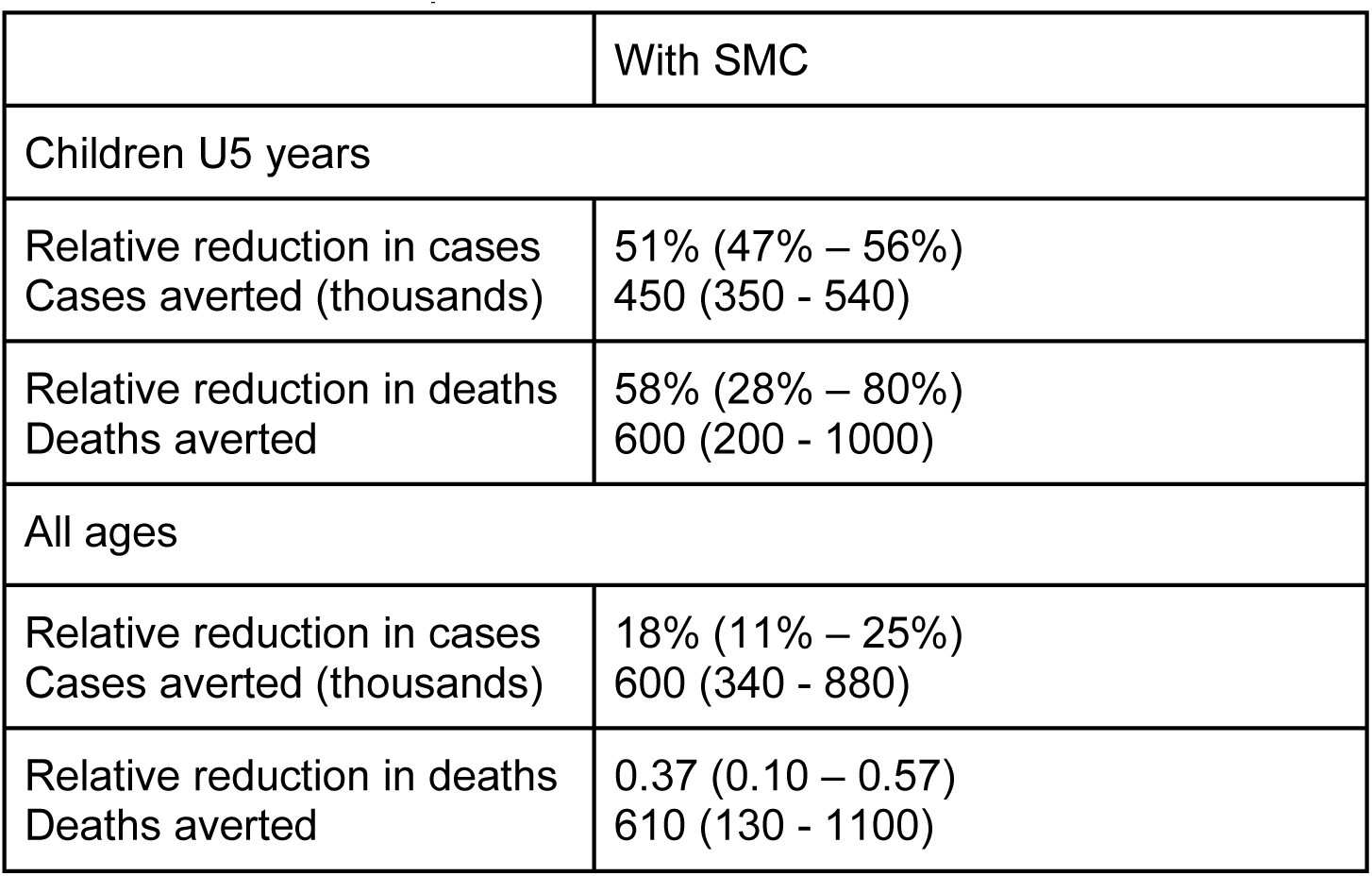
Predicted impact of IG2 deployment if bednet usage were at least 80%, from 2023 to 2027, compared to the base scenario (Figure 4B) with pyrethroid nets in all districts and ITN usage based on the 2021 MIS. The 95% prediction interval is indicated in parentheses.

### SMC eligibility and potential impact of geographic expansion of SMC

Districts eligible for SMC were those with parasite prevalence over 5% in children under 5 years of age (Figure 3C) and seasonal transmission suitable for this intervention. 27 districts met these criteria (Figure 6A). Three observations were made after a visual comparison of trends in cases and rainfall (Figure 6B, Figure S1.14-S1.20 in Supplementary File 1): first, there were nearly always case peaks that occurred after the onset of rain even if those peaks were not detected by the seasonality algorithm; second, case peaks were sometimes longer than rainfall peaks; and last, there were districts with no clear case seasonality but clear rainfall peaks.

**Figure 6:**
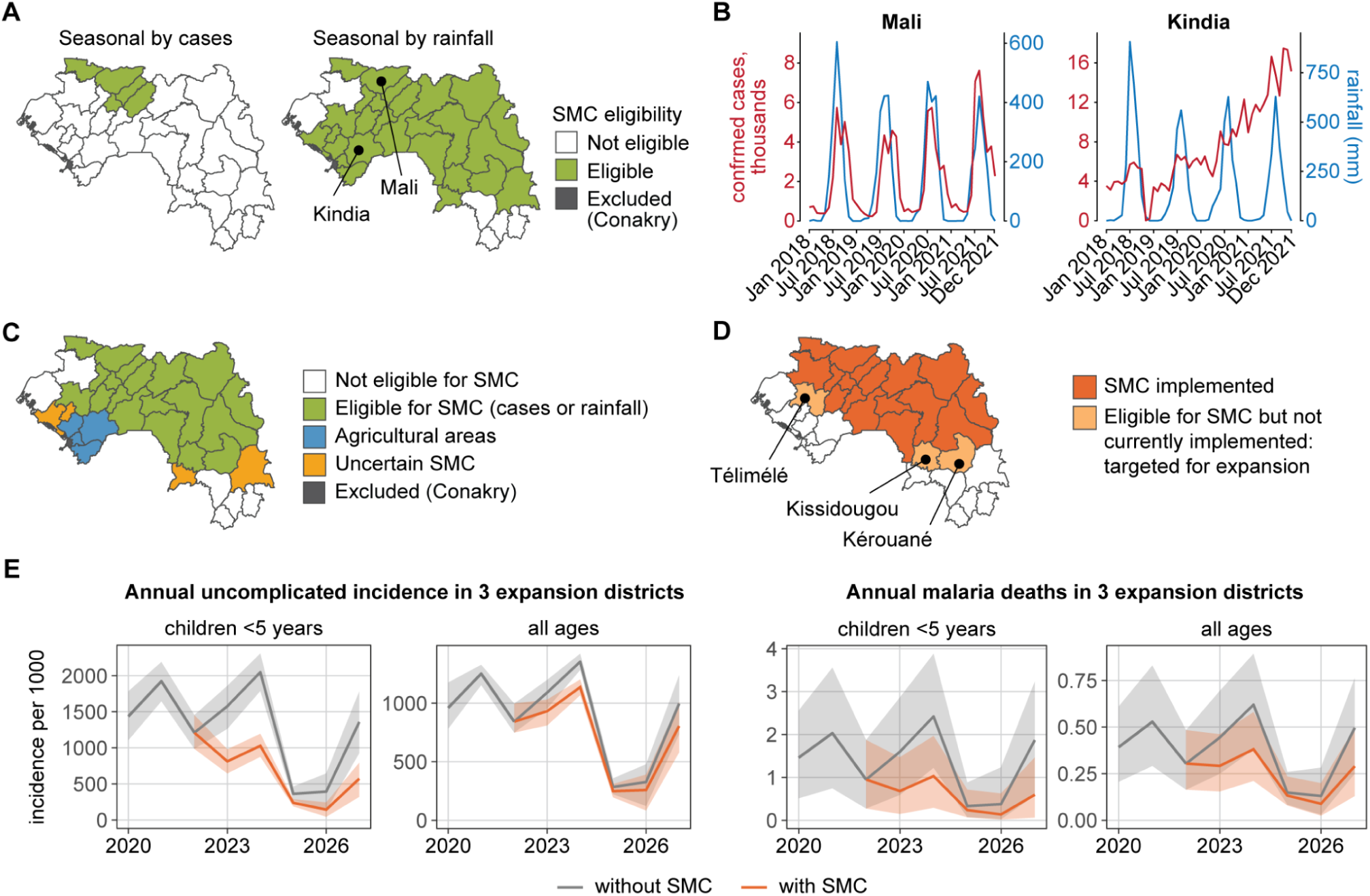
Assessing eligibility for SMC and potential impact of SMC expansion to three additional districts. (A) Health districts eligible for SMC according to analysis of routine cases and of rainfall data. (B) Examples of districts with similar (Mali) and different (Kindia) seasonality in cases and rainfall. (C) Final seasonality classification for SMC eligibility after review of rainfall and cases trends. (D) 22 Districts where SMC had been already implemented as of 2022, and three additional districts selected for SMC implementation. (E) Mean predicted incidence and death rates (solid lines) and their 95% PI (shaded area) from 2020 to 2027 in the three districts targeted for expansion of SMC.

The district-by-district review of seasonality identified a few districts with highly seasonal rainfall, but where the seasonality of cases was not evident. In some districts of Kindia, this phenomenon could be explained by agricultural activities and persistent puddles that remain after rainfall, sustaining transmission and widening the seasonal case peaks. In other districts, seasonality could be affected by the presence of mangroves and swamps. These factors were taken into account when determining the final assignment of SMC eligibility, leaving 19 districts eligible for SMC (Figure 6C).

Three new districts (Télimélé, Kissidougou, and Kérouané) were selected for SMC in addition to the districts where SMC had already been implemented (Figure 6D). Implementation of SMC over five years in the three expansion districts was predicted to avert 51% of cases (95% PI: 47% to 56%) and 58% of deaths (95% PI: 28% to 80%) in children U5 in these districts (Figure 6E, Table 4). Additional cases were predicted to be averted in individuals over 5 years, as some children over 5 nonetheless receive SMC (see Methods).

**Table 4:**
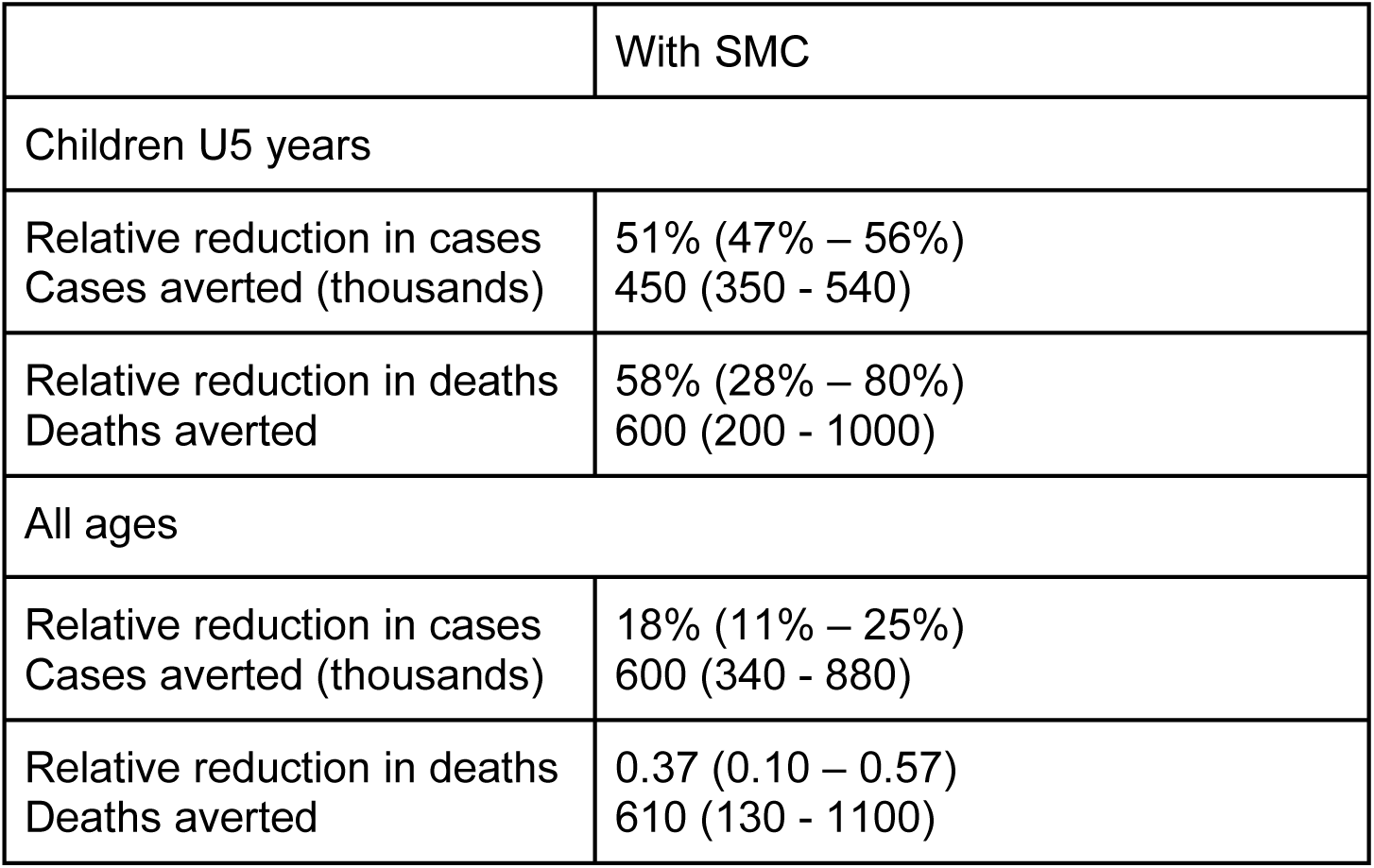
Predicted impact of SMC from 2023 to 2027 in the three additional districts targeted for expansion of SMC, compared with the base scenario without SMC (Figure 5B). The 95% prediction interval is indicated in parentheses.

### Selection of the number and timing of SMC cycles

SMC in Guinea has historically consisted of four monthly cycles. In areas determined to be eligible for SMC (Figure 6D), the median number of confirmed cases in children U5 between 2018 and 2022 was used to determine the duration of the seasonal peak (Figure 7A) by identifying the 4-month or 5-month window when >60% of cases were reported, which translated into 4 or 5 rounds, respectively. Health districts eligible for SMC where <60% of cases were reported in a 5-month window were also considered for 5 rounds acknowledging the previously mentioned limitations of the case data to accurately depict seasonality. The median of the confirmed cases in children U5 was also used to determine the month of peak onset per district (Figure 7B). The impact of adding an additional cycle was evaluated in all health districts targeted for SMC eligible for 5 rounds. The month of peak onset could be detected in the majority of eligible districts as May or June, with the exception of districts in the Kankan region where issues with the quality of routine data made onsets of case peak not detectable with the standard approach.

**Figure 7:**
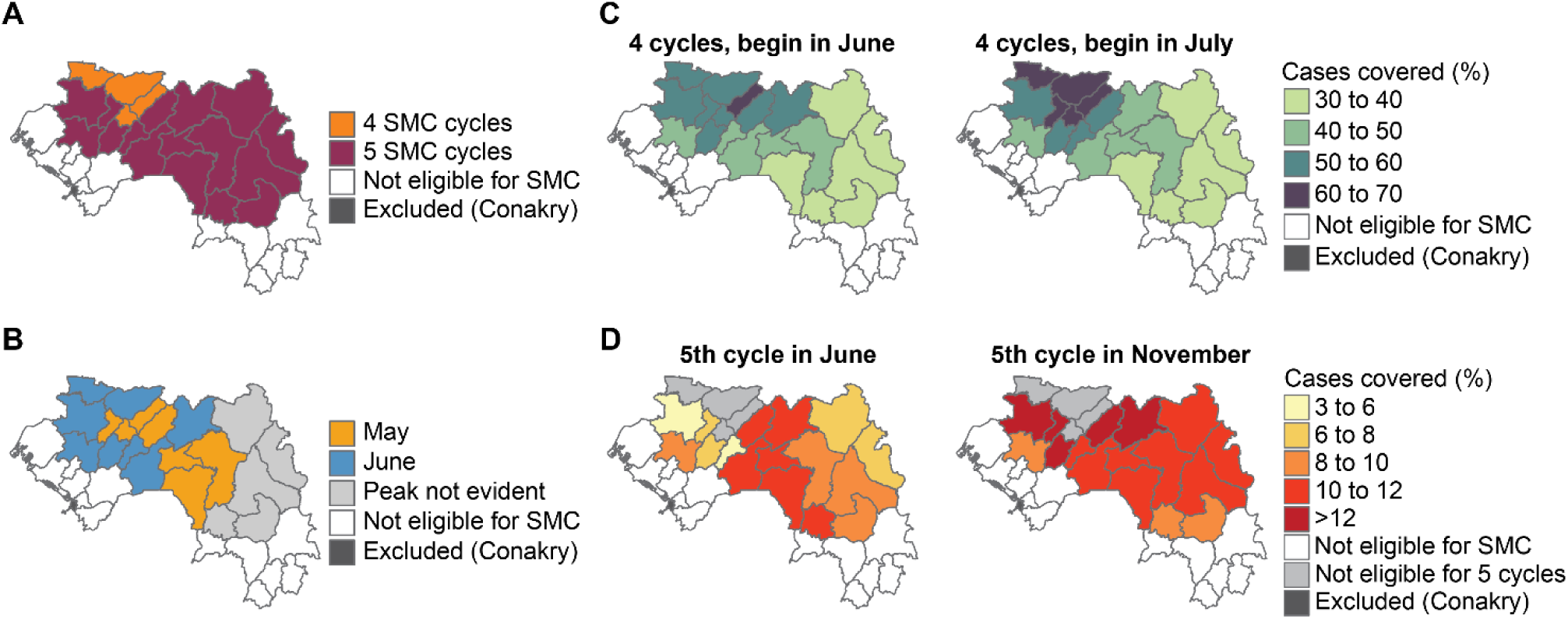
Selection of the number and timing of SMC cycles in Guinea. (A) Number of SMC cycles required to cover the peak transmission season, based on the median confirmed cases U5 between 2018 and 2022. (B) Month in which the median number of cases increased by more than 60% compared with the previous month. (C) Proportion of cases covered by four cycles if the first cycle begins in June. (D) Proportion of cases covered by four cycles if the first cycle begins in July. (E) Proportion of additional cases covered by the 5th cycle of SMC if the cycle is implemented in June. (F) Proportion of additional cases covered by the 5th cycle of SMC if the cycle is implemented in November.

Considering the predominant month of peak onset and for operational purposes, the most appropriate date for the first SMC cycle in a 4-cycle calendar was identified to be either June or July. In most districts, more cases would be covered with SMC cycles between July and October than between June and September (Figure 7C). If a fifth SMC cycle were to be implemented in eligible districts, assuming an existing 4-cycle schedule of July to October, placing the fifth cycle in November would cover more cases than a fifth cycle in June (Figure 7D). The PNLP decided to have five cycles in all eligible districts, although implementation would be subject to availability of resources.

### Targeting of the malaria vaccine under vaccine supply constraints

WHO recommends prioritizing the implementation of either of the two recommended malaria vaccines in areas where the intensity of transmission is moderate or high (23). In Guinea, the majority of health districts met this criterion. At the time when this exercise was conducted, only the RTS,S vaccine was recommended with the availability of very limited supply for Africa (18 million doses). For this purpose, the PNLP followed WHO’s Framework for the allocation of limited malaria vaccine supply (24). Health districts were categorized based on malaria transmission and mortality, where areas with parasite prevalence of at least 20% (Figure 3C) and mortality of at least 9.5 per 1000 live births (Figure 3D), or parasite prevalence of at least 40% and mortality of at least 7.5 per 1000 live births, were identified as the those most in need of vaccine (category 1 areas). 26 health districts met the criteria for the highest level of prioritization (Figure 8). The remaining seven health districts met criteria for second-tier prioritization.

**Figure 8:**
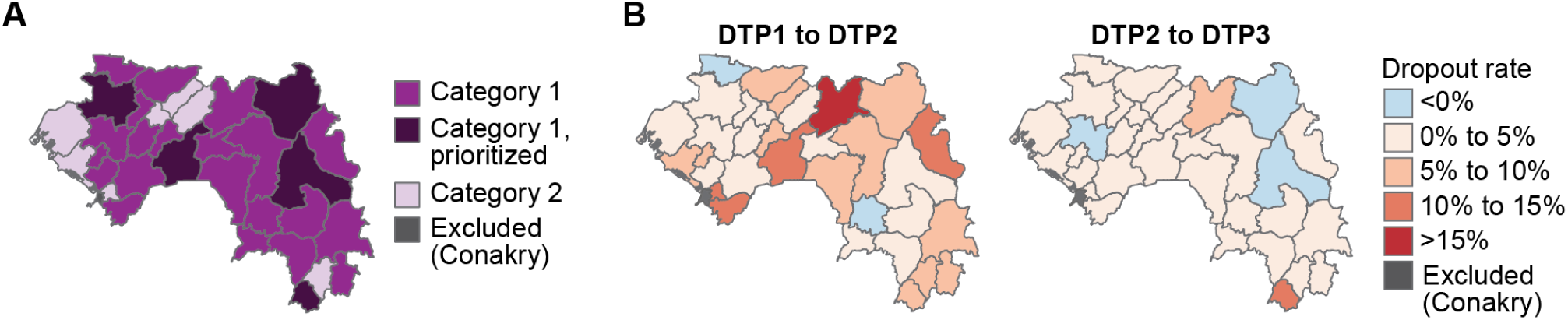
Prioritization of districts due to limited availability of the RTS,S vaccine in 2022. (A) Categorization of Guinea’s health districts according to their malaria prevalence and all-cause mortality following the WHO framework on allocation of limited doses of malaria vaccine, and final districts chosen for vaccine implementation under supply constraints. (B) Dropout rates of DTP1 to DTP2 and DTP2 to DTP3 used to inform targeting of malaria vaccine.

At the time of this analysis, a maximum number of 1 million RTS,S vaccine doses was available per country (to cover approximately 250 thousand children), which needed to be prioritized within the 26 highest-need districts. The PNLP initially intended to use DTP1 to DTP2 and DTP2 to DTP3 drop-out rates to identify the districts with higher-functioning immunization programs that would be more likely to achieve successful implementation of RTS,S with high coverage. However, due to the limited quality of the immunization data the PNLP decided to choose 5 districts from category 1 for pilot malaria vaccine implementation (Gaoual, Mamou, Kankan, Siguiri and Yomou). The districts were selected due to their regional spread, high incidence, and their status as PNLP programmatic priority districts. This plan was since revised to replace RTS,S with R21 after the recommendation of R21, targeting the same districts, with national scale-up to occur subsequently

### Targeting and potential impact of PMC with or without RTS,S

PMC was targeted to districts with a prevalence of at least 10% and not targeted with SMC. Thirteen districts in the regions of N’zérékoré, Kindia, and Boké met these criteria (Figure 9A). The PNLP identified the districts in the N’zérékoré region for priority implementation (Figure 9B) as these were the eligible districts with high or very high transmission according to the risk stratification (Figure 3G).

**Figure 9:**
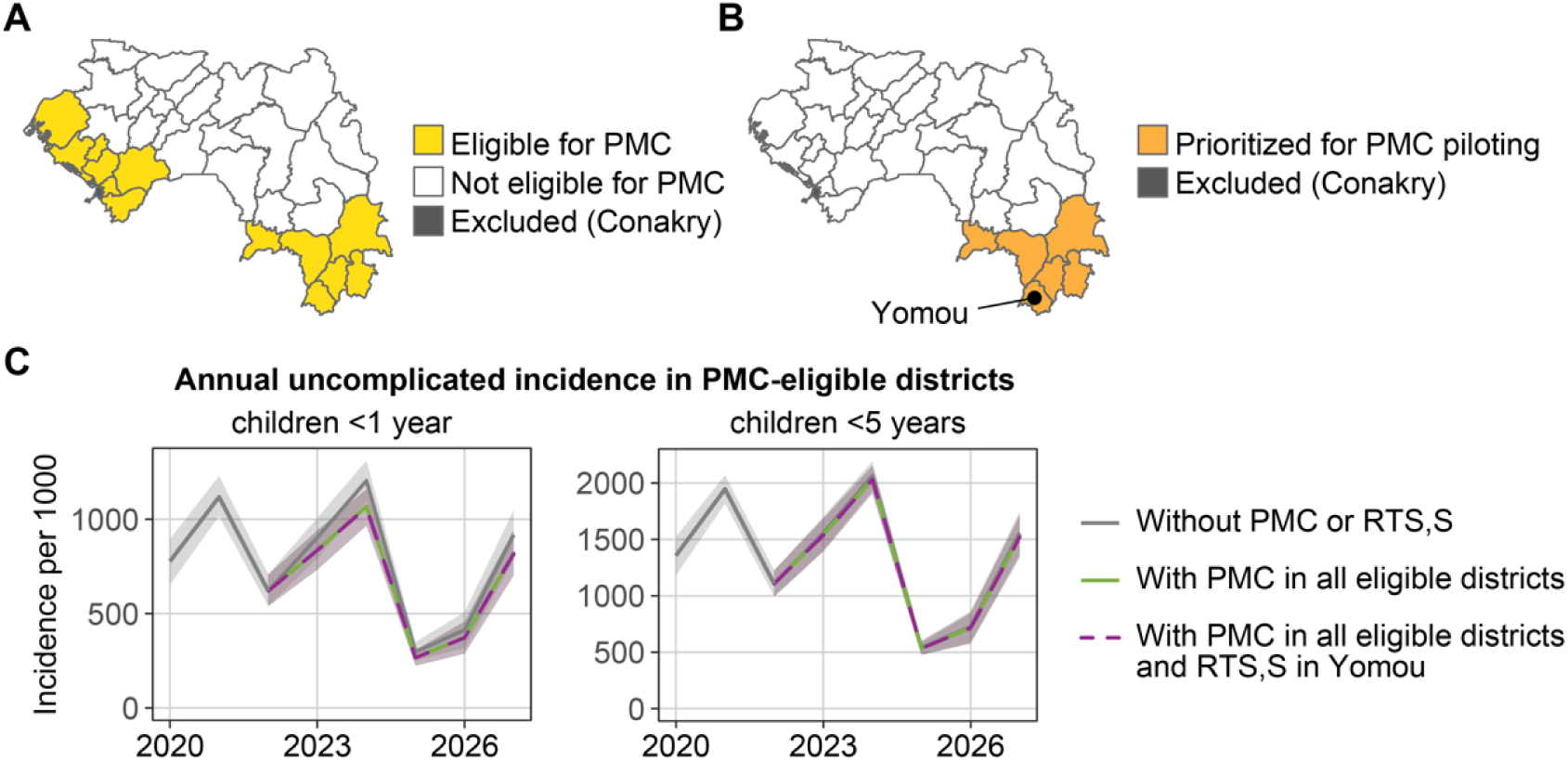
Targeting and potential impact of PMC in Guinea. (A) Districts eligible for PMC. (B) Priority districts for PMC piloting. (C) Mean predicted incidence (solid line) and their 95% PI (shaded area) from 2020 to 2027 for all 13 districts eligible for PMC. In addition to PMC, malaria vaccine RTS,S may be available to the Yomou district and is factored into the prediction here.

The potential impact of PMC if it were to be implemented in all eligible districts (Figure 9A), with or without co-implementation of RTS,S in Yomou District (one of the selected districts for the piloting of the malaria vaccine), was predicted using mathematical modeling. Other districts prioritized for RTS,S were not also eligible for PMC. Since PMC doses are given at ten weeks, fourteen weeks, and nine months of age, and the protective effect of PMC lasts for around one month, PMC was not expected to have any effect in children older than one year of age, and thus impact was assessed for cases averted in children under the age of 1 year (U1) in addition to children U5. The impact on deaths was not predicted due to high uncertainty in events with very small numbers.

The model predicted that implementation of PMC from 2023 to 2027 could avert 10% of cases (95% PI: 5% to 14%) in children U1 in the 13 districts eligible for PMC (Figure 9, Table 5). In Yomou district, addition of RTS,S averted an additional 7% of cases (95% PI: -9% to 21%) in children U1. PMC averted no additional cases in children above the age of one year, as expected due to the duration of protection of PMC. Addition of RTS,S averted 6% of cases (95% PI: -11% to 20%) in children between 1 and 5 years of age. The model stochasticity was larger than the impact of RTS,S, resulting in negative values in the predictive interval.

**Table 5:** Predicted impact of PMC with or without RTS,S from 2023 to 2027, in all 13 districts eligible for PMC and in Yomou district only, compared to the base scenario (Figure 4B) with no PMC and no RTS,S. The 95% prediction interval is indicated in parentheses.

|  | All 13 districts | Yomou district |  |
| --- | --- | --- | --- |
|  | With PMC only | With PMC only | With PMC and RTS,S |
| Children U1 |  |  |  |
| Relative reduction in cases | 10% (5% – 14%) | 11% (-4% – 27%) | 18% (3% – 31%) |
| Cases averted (thousands) | 53 (27 - 78) | 2.6 (-0.8 - 6.4) | 4.0 (0.5 - 7.6) |
| Children U5 |  |  |  |
| Relative reduction in cases | 1% (-2% – 4%) | 1% (-14% – 13%) | 7% (-7% – 18%) |
| Cases averted (thousands) | 62 (-90 - 202) | 1.8 (-22.8 - 21.3) | 12.7 (-11.6 - 32.4) |

## Discussion

Application of the SNT process resulted in improved understanding of recent dynamics in malaria transmission, identification of specific areas to prioritize and to maximize the impact of interventions, and an assessment of the availability and quality of data needed for clear decision-making. SNT provided solid support to justify programmatic decisions in Guinea’s Global Fund funding request by demonstrating the expected significant impact of the expanded intervention package on malaria transmission. Guinea’s Global Fund funding request was ultimately successful at securing funding for IG2 nets in all 33 districts, expanding SMC to three new districts, and implementing five cycles of SMC in all districts eligible for five cycles.

The use of data for sound decision-making relies on the availability of sufficient data of appropriate quality. A surveillance assessment would allow the PNLP to systematically assess the quality of its surveillance system and identify areas for improvement (14), but insights were also gained during the SNT process. Missingness of key malaria indicators in the routine data was low, testing and reporting rates were high, and there was good internal consistency between indicators. This level of quality demonstrates the long-standing efforts made by the PNLP and its partners to avoid stock-outs for diagnostic and treatment products and to improve data collection and reporting via the DHIS2 platform. However, other indicators such as data on inpatient admissions or deaths were not well-reported, and alternative proxies such as modelled all-cause under-five mortality rates had to be used. Furthermore, a substantial effort was required during the SNT process to assemble all routine and non-routine data. The PNLP would benefit from having a more structured malaria data repository containing all relevant data for decision-making.

In Guinea, most facilities in the DHIS2 are in the public sector, and these facilities report more information than those in the private sector. A similar pattern is observed in other countries (25,26). In some health districts in Guinea, private health facilities may send their data through public health facilities located in their geographical area. Non-reporting or missing data from those private facilities would therefore not be visible during data quality assessments.

Adjusting for care-seeking behavior outside of the public health sector substantially increased the estimated incidence. Nevertheless, accurately estimating care-seeking remains difficult: fine-scale spatio-temporal data is lacking, including at the district level, and estimates for care-seeking in individuals above five years of age are often unavailable (27). Furthermore, the relationship between care-seeking reported in surveys and actual patterns of care-seeking for symptomatic malaria remains unclear. In Guinea, the PNLP did not find the incidence estimated after adjusting for care-seeking to reflect actual epidemiological conditions, and instead chose to use incidence adjusted only for testing and reporting rates to identify relative differences in burden between districts while acknowledging that this incidence was still likely to be an underestimate of the actual incidence. Better data and refined estimation approaches are needed to understand the true burden of malaria.

Until measurements of malaria prevalence or mortality are sufficiently frequent and available at relevant spatial resolution in Guinea, spatio-temporal modelled estimates will be crucial for decision-making. To improve acceptability and utility from the PNLP, estimating these metrics should shift from a global to a local approach, incorporating locally collected subnational routine and non-routine information and adjusting methodologies to fit the country’s context through strong partnerships that focus on developing local capacity.

The SNT process highlighted the limited availability and quality of data on determinants of malaria transmission. Under constrained resources, vector control with dual-action insecticide ITNs or with IRS should be targeted at areas where vectors are resistant to lower-cost pyrethroid insecticides (23). Ideally, entomological surveillance would inform the SNT team of the degree of insecticide resistance in each district. However, in practice entomological data is collected in a few sentinel sites across Guinea, so while it is broadly informative of regional levels of insecticide resistance, it is not powered to inform at the district level. Good data on aspects of intervention implementation that could inform effectiveness at the routine and community level were also lacking. This limited the PNLP’s ability to interpret epidemiological data, assess gaps, and make accurate predictions of local intervention impact.

Impact predictions from the mathematical model have several key limitations, in addition to the limitations due to data inputs to the model already described above. Model results are limited by uncertainty around intervention coverages and effect sizes, especially at the district level; uncertainty of vector behavior and susceptibility to insecticides; uncertainty in the true burden of malaria in each district; and uncertainty in case fatality rates of treated and untreated malaria. Due to the time constraints imposed by funding request deadlines, it was not possible to fully explore the sensitivity of model predictions to uncertainties in key parameters. However, future work would benefit from a quantitative understanding of impact of uncertainties, particularly to inform future data collection, via sensitivity analysis or more complex and comprehensive methods to estimate joint uncertainties.

Consensus-building and constant review were essential elements of the SNT process, and consequently, SNT can only be automized to a certain extent. For example, Guinea’s analysis of seasonality found that seasonality of rainfall and cases were not always the same, and districts with similar rainfall could have very different case patterns. The SNT team reviewed and discussed the seasonality by rainfall and cases on a district-by-district basis, bringing in local knowledge to guide the interpretation of the analysis results, and together made a determination for each district. A full automation of the SNT process would not have resulted in such a nuanced and informed determination of SMC eligibility, nor would it have given the PNLP and its partners a greater understanding of the utility and limitations of key data sources. Similarly, automation of the epidemiological stratification without adaptation after PNLP review would have misclassified the district of N’zérékoré as low risk.

By including partners at every step of the SNT process, under the leadership of the PNLP, there was team ownership of the results by the entire SNT team. The SNT team reviewed and discussed the data at every step, and decisions for whether and how to include data in the decision-making process were undertaken together, with the ultimate decisions made by the PNLP. Issues with quality or availability of certain data were identified for follow-up and future data collection. Bringing together all partners from the beginning enabled the SNT team to discuss and resolve sensitive issues together, and partners acquired a greater understanding of why decisions were made, resulting in greater buy-in for the decisions.

Capacity does not yet exist within the PNLP to conduct every step of SNT without external support. Strong leadership, capacity to translate evidence and analytical outputs into policy, and thorough understanding of data availability and quality were already present at the initiation of this SNT exercise. For data management and analysis, the PNLP relied on external partners. The PNLP grew its capacity in data management and simpler analysis such as the epidemiological stratification during this SNT, as well as during workshops conducted by WHO after the SNT, and plans to be able to conduct these steps internally in future rounds of SNT. However, other analytical steps such as geostatistical modeling and mathematical modeling may take much longer before capacity is present within the PNLP.

Prior to the use of SNT, Guinea’s planning process used a homogeneous approach that applied interventions evenly in all districts regardless of suitability. The SNT process enabled the PNLP to, for the first time, use a targeted approach in their intervention planning and focus resources where they could have the most impact. The success of the process spurred the PNLP to adapt the principles of SNT to new questions after the submission of their funding request, including a microstratification of Conakry, a retrospective analysis of trends in incidence, and an exercise to reprioritize bednets under further resource constraints.

## Conclusions

In the face of continued high burden of malaria, Guinea’s national malaria program adopted an innovative data-informed process, guided by local expertise and engaging multiple partners, to prioritize malaria interventions at the district level and successfully obtain funding for expanded intervention plans. This analytic approach was unprecedented in Guinea and allowed the PNLP to effectively decide how to prioritize their limited resources.

## Supporting information

Supp File 1: Data Management and Stratification

Supp File 2: Mathematical Modeling

## List of abbreviations

ACT: artemisinin-based combination therapy
DHIS2: District Health Information System 2
DHS: Demographic and Health Survey
DTP: Diphtheria-Tetanus-Pertussis vaccine
EPI: Expanded Programme on Immunization
IG2: Interceptor G2
IHME: Institute for Health Metrics and Evaluation
IPTp: Intermittent Preventive Treatment in pregnancy
IRS: indoor residual spraying
ITN: insecticide-treated net
MAP: Malaria Atlas Project
MIS: Malaria Indicator Survey
*Pf*PR: *Plasmodium falciparum* parasite rate
PMC: Perennial Malaria Chemoprevention
PMI: US President’s Malaria Initiative
PNLP: Programme National de Lutte contre le Paludisme
RDT: Rapid Diagnostic Test
SMC: Seasonal Malaria Chemoprevention
SNT: subnational tailoring of interventions
SP-AQ: Sulfadoxine-Pyrimethamine with Amodiaquine
U1: Under-1 (year of age)
U5: Under-5 (years of age)
U5MR: All-cause under-5 Mortality Rate
WHO: World Health Organization

## Declarations

### Ethics approval and consent to participate

This study was determined by the Northwestern University Institutional Review Board to not constitute human subjects research (STU00220182).

### Consent for publication

Not applicable.

### Availability of data and materials

All code used for data analysis and modeling is available online at https://github.com/numalariamodeling/snt_gin , as are all model inputs. DHS and MIS data are available online at dhsprogram.org. Inquiries regarding routine malaria surveillance data in Guinea may be directed to the PNLP at.

### Competing interests

The authors declare that they have no competing interests.

### Funding

OOD, KBT, MR, and JG were supported by a grant from the Bill & Melinda Gates Foundation (INV-048410).

### Authors’ contributions

Study conceptualization: AD, BG, AC. Data assembly and management: OOD, AD, ND, MD, EMD. Data review: OOD, AD, ND, MD, EMD, BG, AC. Data analysis: OOD, JG, BG, AC. Modeling: OOD, KBT, MR, TS, JG. Interpretation: all authors. First draft manuscript: OOD, AD, KBT, MR, TS, JG, BG. All authors have read and approved the final manuscript.

## Data Availability

https://dhsprogram.org

## Acknowledgements

The authors would like to thank PMI Guinea, country teams of the Global Fund and Catholic Relief Services, and RTI/Notre Sante for their support of this work. The authors would also like to thank Northwestern University Research Computing for their assistance with high-performance computing needs. All simulations were run on the Quest computing platform at Northwestern University.

## Disclaimer

Beatriz Galatas is a staff member of the World Health Organization. This author alone is responsible for the views expressed in this article and does not necessarily represent the decisions, policy or views of the World Health Organization.

## Notes

### Competing Interest Statement

The authors have declared no competing interest.

## References

1. Division Information Sanitaire et Recherche. Annuaire Statistique Sanitaire 2021. Conakry, Guinée, 2022.

2. Programme National de Lutte Contre le Paludisme de Guinée. Bulletin annuel de surveillance du paludisme. Conakry, Guinée, 2023.

3. WHO. World malaria report 2023. Geneva, World Health Organization, 2024.

4. WHO. Guiding principles for prioritizing malaria interventions in resource-constrained country contexts to achieve maximum impact. 2024. Geneva, World Health Organization, 2024.

5. MAGIC Evidence Ecosystem Foundation. Une Plateforme de creation et de publication numérique pour l’écosystème des preuves. https://app.magicapp.org/%23/guidelines. Accessed 6 June 2024

6. Institut National de la Statistique. Recensement Général de la Population et de l’Habitat 2014. Conakry, Guinée, 2014.

7. Demographic and Health Surveys (DHS) Program. https://dhsprogram.com Accessed 6 June 2024

8. Hersbach H, Bell B, Berrisford P, Biavati G, Horányi A, Muñoz Sabater J, et al. ERA5 hourly data on single levels from 1940 to present. Copernicus Climate Change Service (C3S) Climate Data Store (CDS). Ecmwf. 2023;147.

9. PMI VectorLink Project Guinea. Guinea entomological monitoring annual report, June 2022. Rockville, MD, PMI VectorLink Project, Abt Associates Inc., USA; 2022.

10. WHO. Malaria Threats Map: tracking the spread of biological threats to malaria 2023. Geneva, World Health Organization, 2023.

11. Institut National de la Statistique et ICF International. Enquête Démographique et de Santé et à Indicateurs Multiples (EDS-MICS). Ministère de la santé publique et Institut national de la statistique, Conakry, Guinée, 2012.

12. Institut National de la Statistique et ICF International. Enquête sur les indicateurs du paludisme et de l’anémie en Guinée (EIPAG). Ministère de la santé publique et Institut national de la statistique, Conakry, Guinée, 2021.

13. Institut National de la Statistique et ICF International. Enquête Démographique et de Santé (EDS). Ministère de la santé publique et Institut national de la statistique, Conakry, Guinée, 2018.

14. WHO. Malaria surveillance, monitoring, and evaluation: a reference manual. Geneva, World Health Organization, 2018.

15. Tasmin L, Symons JJM, et al. Informing stratified malaria control for impact: a semi-mechanistic, multi-metric, Bayesian geostatistical model for Mozambique. In prep. 2024.

16. GBD 2019 Diseases and Injuries Collaborators. Global burden of 369 diseases and injuries in 204 countries and territories, 1990–2019: a systematic analysis for the Global Burden of Disease Study 2019. Lancet. 2020; 396:1204–22.

17. WHO. Consolidated guidelines for malaria. Geneva, World Health Organization, 2022.

18. Funk C, Peterson P, Landsfeld M, Pedreros D, Verdin J, Shukla S, et al. The climate hazards infrared precipitation with stations - A new environmental record for monitoring extremes. Sci Data. 2015;2.

19. Bershteyn A, Gerardin J, Bridenbecker D, Lorton CW, Bloedow J, Baker RS, et al. Implementation and applications of EMOD, an individual-based multi-disease modeling platform. Pathog Dis. 2018;76(5).

20. Ozodiegwu ID, Ambrose M, Galatas B, Runge M, Nandi A, Okuneye K, et al. Application of mathematical modelling to inform national malaria intervention planning in Nigeria. Malar J. 2023;22(1).

21. WHO. A Framework for Malaria Elimination. Geneva, World Health Organization, 2017.

22. Camara A, Guilavogui T, Keita K, Dioubaté M, Barry Y, Camara D, et al. Rapid epidemiological and entomological survey for validation of reported indicators and characterization of local malaria transmission in Guinea, 2017. American Journal of Tropical Medicine and Hygiene. 2018;99(5).

23. WHO. Guidelines for malaria. Geneva, World Health Organization. https://app.magicapp.org/%23/guideline/LwRMXj/section/LpOA4j. Accessed 6 June 2024

24. WHO. Framework for the allocation of limited malaria vaccine supply. Geneva, World Health Organization, 2022.

25. Githinji S, Oyando R, Malinga J, Ejersa W, Soti D, Rono J, et al. Completeness of malaria indicator data reporting via the District Health Information Software 2 in Kenya, 2011-2015. Malar J. 2017;16(1).

26. Muhoza P, Tine R, Faye A, Gaye I, Zeger SL, Diaw A, et al. A data quality assessment of the first four years of malaria reporting in the Senegal DHIS2, 2014– 2017. BMC Health Serv Res. 2022;22(1).

27. Ozodiegwu ID, Ambrose M, Battle KE, Bever C, Diallo O, Galatas B, et al. Beyond national indicators: adapting the Demographic and Health Surveys’ sampling strategies and questions to better inform subnational malaria intervention policy. Malar J. 2021;20(1).

