## Supplementary material for "Subnational tailoring of malaria interventions to prioritize the malaria response in Guinea": Supp File 1: Data Management and Stratification

Supplementary file 1: Data management, stratification, and interventions

### Data management

#### Defining active health facilities

Due to the absence of a master list of health facility functionality, activity was defined as the point at which the health facility reported at least some information on confirmed cases. Thus, a health facility is considered active when it has started to send a report. This activity is used to calculate the health district's completeness rate by month and year (Figure S1.1).

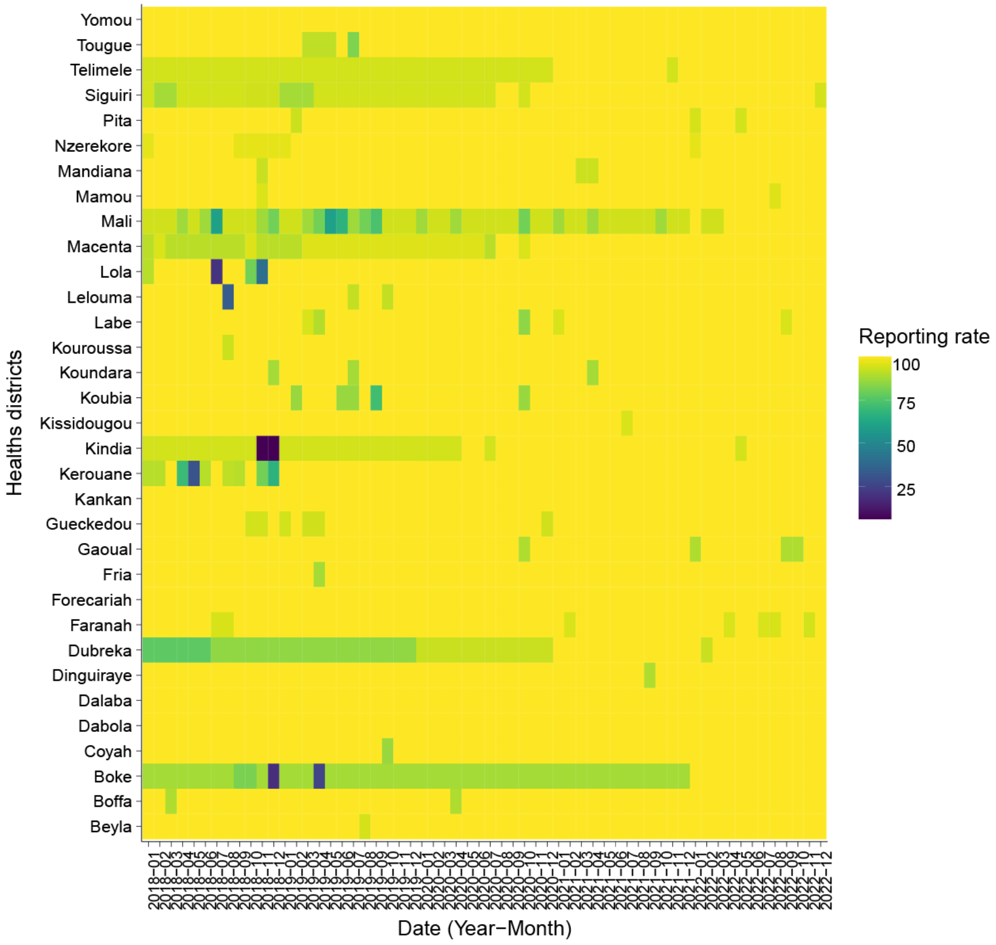

Figure S1.1: Reporting rate of confirmed cases, by health district and month.

#### Missing data and outlier detection

We estimated the proportion of missing data for the 2018-2022 period for each of the following indicators: all-cause outpatients visits, suspected cases, tested cases, confirmed cases, treated cases, hospitalization and deaths due to malaria. We found that malaria deaths and all-cause outpatient visits were the variables with the most missing data in the health facility reports (~13% of missing values) (Figure S1.2). Limited reporting of malaria deaths was explained by the fact that not all health facilities included in the denominator of this calculation are hospitals with inpatient services. Reporting of all-cause outpatient visits improved between 2018 and 2022.

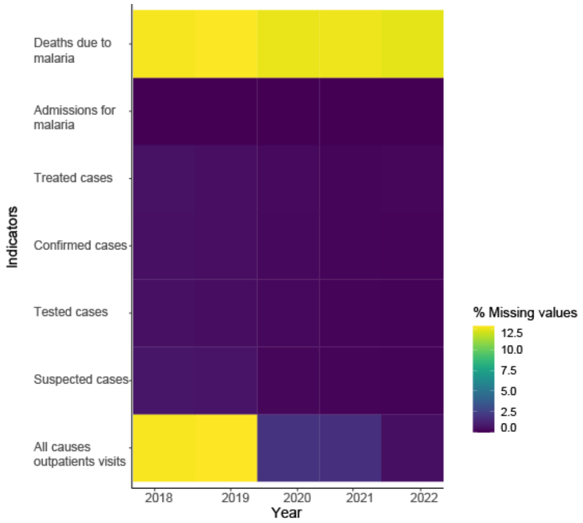

Figure S1.2: Proportion of health facilities missing specific data for each type of variable per year.

Outliers in key indicators at the health facility-month level were identified with a visual examination during discussion with the PNLP (Figure S1.3). Outliers were corrected using the moving average of the values before and after the outlier (sum of previous and next month values divided by 2). 14 outlier values for different indicators (3, 3, 3, 3, 1, 1 respectively for suspected cases, tested cases, confirmed cases, treated cases, all malaria admissions and all outpatient visits) were corrected over the study period. A list of outliers and corrected values was prepared for the PNLP in case the values could also be corrected in DHIS2.

A

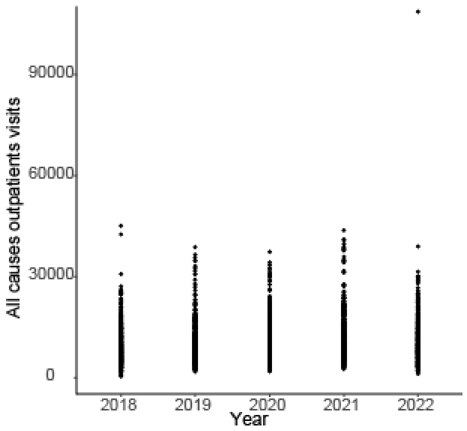

B

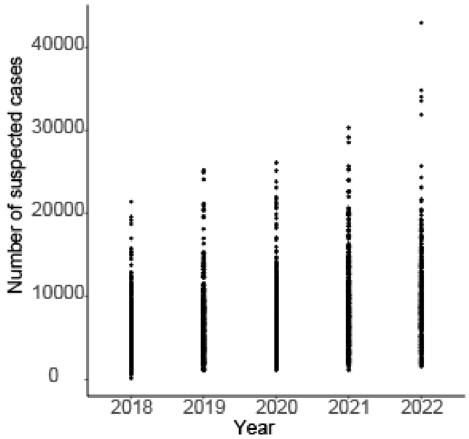

C

D

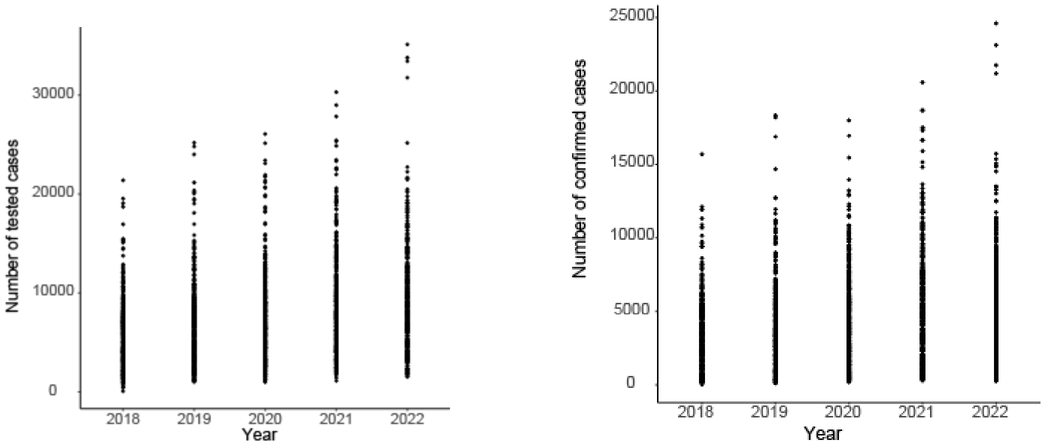

E

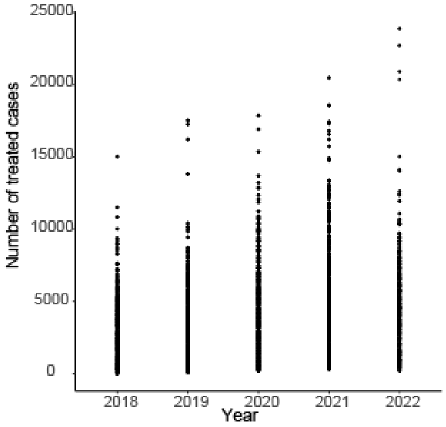

Figure S1.3: Outlier detection using graphical representation. A) All-cause outpatient visits. B) Number of suspected cases. C) Number of tested cases. D) Number of confirmed cases. E) Number of treated cases.

#### Review of data coherence

A consistency check after outlier replacement was performed on various indicators in the routine surveillance data by examining scatterplots where each point represents a health district (HD)-month record (Figure S1.4).

1. Tested cases vs all-cause outpatient visits

Logically, the number of patients tested should not exceed the number of all-cause outpatient visits, as the number of patients tested comes from all-causes outpatient visits. While this is true in most cases (for most points, the number of all-cause visits exceeds the number tested), however, we find that some health facilities report more patients tested than all-cause consultations. This suggests problems with the quality of the data and highlights the need to improve the surveillance system and data quality control in the future.

2. Suspected vs tested cases

In general, all suspected cases (cases with fever) of malaria should be tested at the health facility level. If there are stockouts of tests, tested cases may be fewer than suspected cases. We found that the number of suspected cases was similar to the number of cases tested, with the exception of one health facility, indicating a strong practice of testing cases in most facilities.

3. Tested cases vs confirmed cases

Because confirmed cases are suspected cases with a positive test, the number of confirmed cases should not exceed the number of tested cases. Reassuringly, all health facilities reported a higher number of tested cases than confirmed cases.

4. Confirmed cases vs treated cases

Ideally, the number of confirmed cases and number of treated cases (including for severe treatment) should be equal: every diagnosed case of malaria should receive treatment. However, that these two values were generally not equal. In some cases, the number of treated cases was higher than the number of confirmed cases; in other cases, the number of treated cases was substantially lower than the number of confirmed cases. This reflects poor adherence to management protocols and a high number of cases treated on the basis of presumption, as well as data quality issues.

5. All-cause admissions vs admissions for malaria

Only one health facility reported a higher number of patients admitted for malaria than patients admitted for all causes for the years. This shows excellent consistency but the need remains for data verification at facility and hospital level.

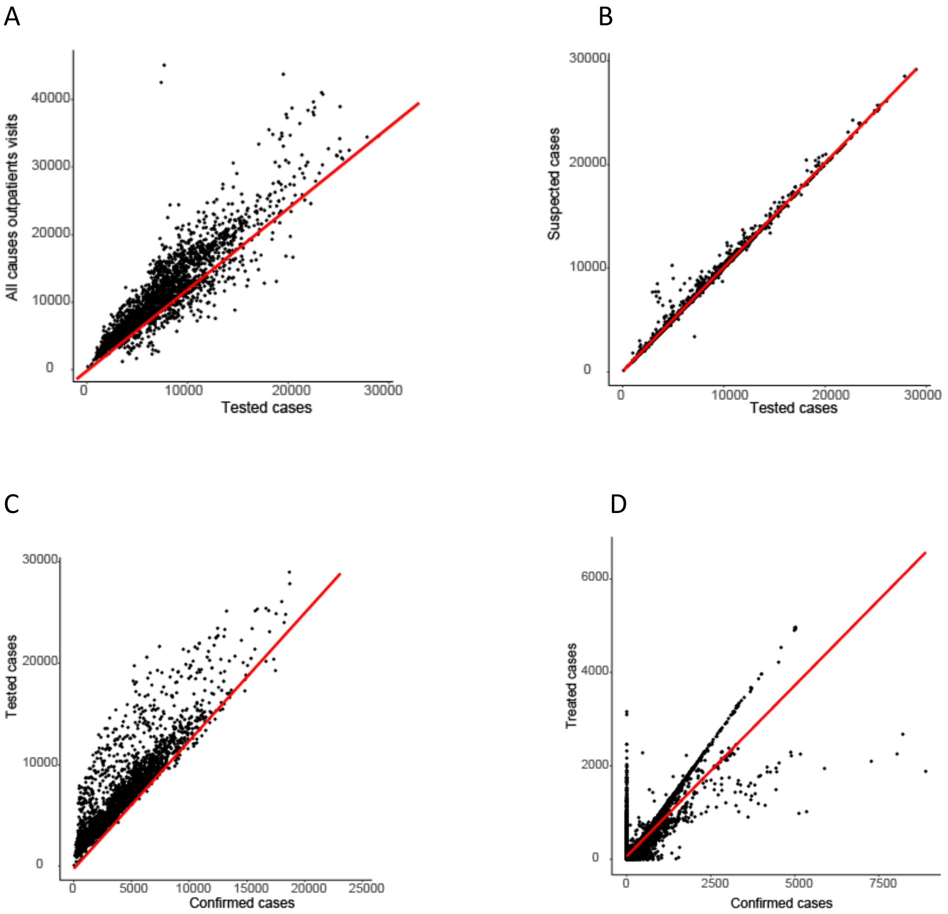

E

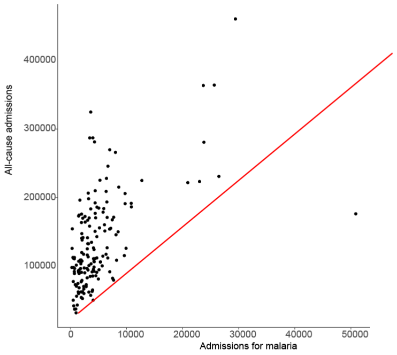

Figure S1.4: Coherence checks on key indicators in routine surveillance data. The red line indicates the identity line. A) All-cause outpatient visits vs tested cases. B) Suspected cases vs tested cases. C) Tested cases vs confirmed cases. D) Treated cases vs confirmed cases. E) All-cause admissions vs admissions for malaria.

### Stratification

#### Incidence adjustment methods

These methods follow the standard methods used by WHO to estimate incidence of malaria cases from routine data (WHO 2023).

Annual crude incidence was estimated by dividing the aggregated crude and adjusted number of confirmed cases (C) by the official population per health district and multiplying by 1000.

As rapid diagnostic tests (RDTs) have become increasingly acceptable and available over the years, the number of suspected cases tested by RDT continues to rise. Changing testing practices can also influence case numbers. To account for incomplete testing rate, the number of additional cases per health district and month that would have been confirmed had all suspected cases been tested was estimated by multiplying the number of presumed cases (P) by the test positivity rate for the same district and month. The presumed cases were calculated by subtracting the number of tested cases from the number of suspected cases given that this information is not directly captured by the PNLP. The test positivity rate was calculated by dividing the number of confirmed cases by the number of tested cases (T). The number of expected additional cases was added to the number of confirmed cases for the same district and month (equation 1) to generate a new estimate of cases (N1). N1 was aggregated to the year and divided by the district population and multiplied by 1000 to obtain the first level of adjusted incidence. This adjustment assumes that the test positivity rate in the presumed cases would have been similar to the test positivity rate in the tested cases.

$$N1 = C + [P * C/T] \qquad (1)$$

To account for incomplete reporting rate, N1 was divided by the estimated reporting rate per district-month (R) (equation 2). The monthly reporting rate for confirmed cases was calculated by dividing the number of reports received by the number of reports expected (Figure S1.1 in Supplementary File 1). A health facility was considered active after the first month of reported confirmed malaria cases. The resulting number of cases adjusted for varying testing and reporting rates (N2) were aggregated to the year and divided by the district population and multiplied by 1000 to obtain the second level of incidence adjustment.

$$N2 = N1/R \qquad (2)$$

To account for care-seeking behavior for a fever outside of the public health sector, the number of additional cases that would have been reported if all malaria-attributable fevers were treated in this sector was estimated by adding the number of additional cases expected from the private health sector (by multiplying N2 by the ratio of the care-seeking rate in the private sector (CSpr) to the care-seeking rate in the public sector (CSpu)) and those fevers that do not seek care (by multiplying N2 by the ratio of half of the rate of not seeking care for a fever (CSn) to the care-seeking rate in the public sector). This adjustment assumes that the infection rate is the same for those seeking care in the public sector, in the private sector, or among those who do not seek care. It also assumes that care-seeking behavior for any fever represents that of a malaria-attributable fever, and that the patterns observed in children are similar in the over-five population.

$$N3 = N2 + (N2 * CSpr/CSpu) + (N2 * CSn/CSpu) \quad (3)$$

Care-seeking rates were extracted from the DHS 2018 and MIS 2021 surveys. The care-seeking rates were defined as the proportion of children U5years who had fever within the last two weeks and 1) sought care in the public sector, 2) sought care in the private sector (REF), or 3) did not seek care (Figures S1.5 and S1.). Children who reported seeking care in both the public and private sectors were included only in the public health sector category. Care-seeking rates were extracted at the regional level by aggregating information by cluster, accounting for cluster weights [REF]. The same rates were applied to all districts in a region. A linear interpolation between the two available surveys of 2018 and 2021 was used to estimate the care-seeking behavior patterns per province in the year without surveys (2019, 2020 and 2022) (Figure S1.6).

#### Treatment-seeking rates

We estimated care-seeking rates at regional level using the 2018 DHS and 2021 MIS through 3 indicators: the proportion of children with fever who reported seeking care from the public sector, the proportion of children with fever who reported seeking care from the private sector, and the proportion of children with fever who reported not seeking care (Figure S1.5). Children seeking care in both the private and public sectors were considered only in the public count, as presumably their case would be recorded at the health facility in routine surveillance data. For years without surveys, for each source of care, a linear interpolation was performed by fitting a linear regression between year and care-seeking rate, adjusting for the region, and also extrapolated to 2022 (Figure S1.6).

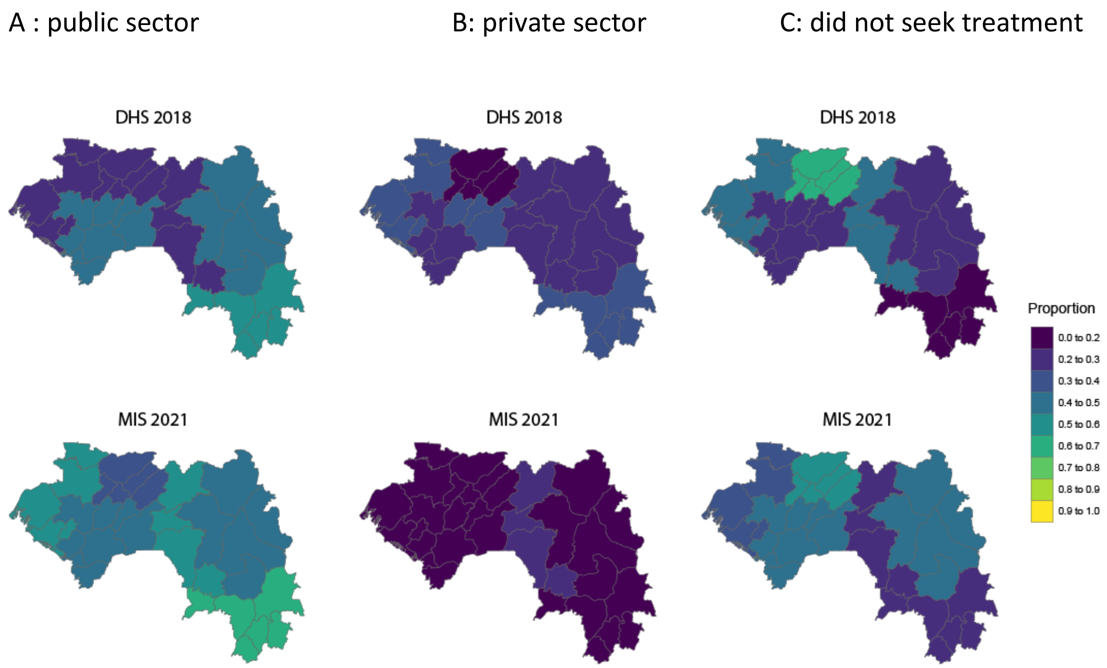

Figure S1.5: Care-seeking rates at regional level in the 2018 DHS and the 2021 MIS. A) Proportion of children with recent fever who reported seeking care from the public sector. B) Proportion of children with fever who reported seeking care from the private sector. C) Proportion of children with fever who reported not seeking care.

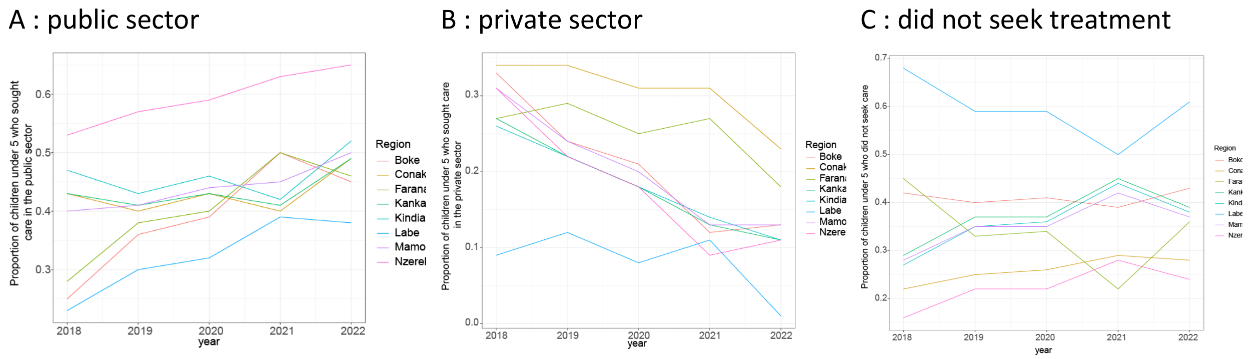

Figure S1.6: Linear interpolation of care-seeking rates at regional level. A) Proportion of children with recent fever who reported seeking care from the public sector. B) Proportion of children with fever who reported seeking care from the private sector. C) Proportion of children with fever who reported not seeking care.

#### Crude and adjusted incidence for 2018-2022

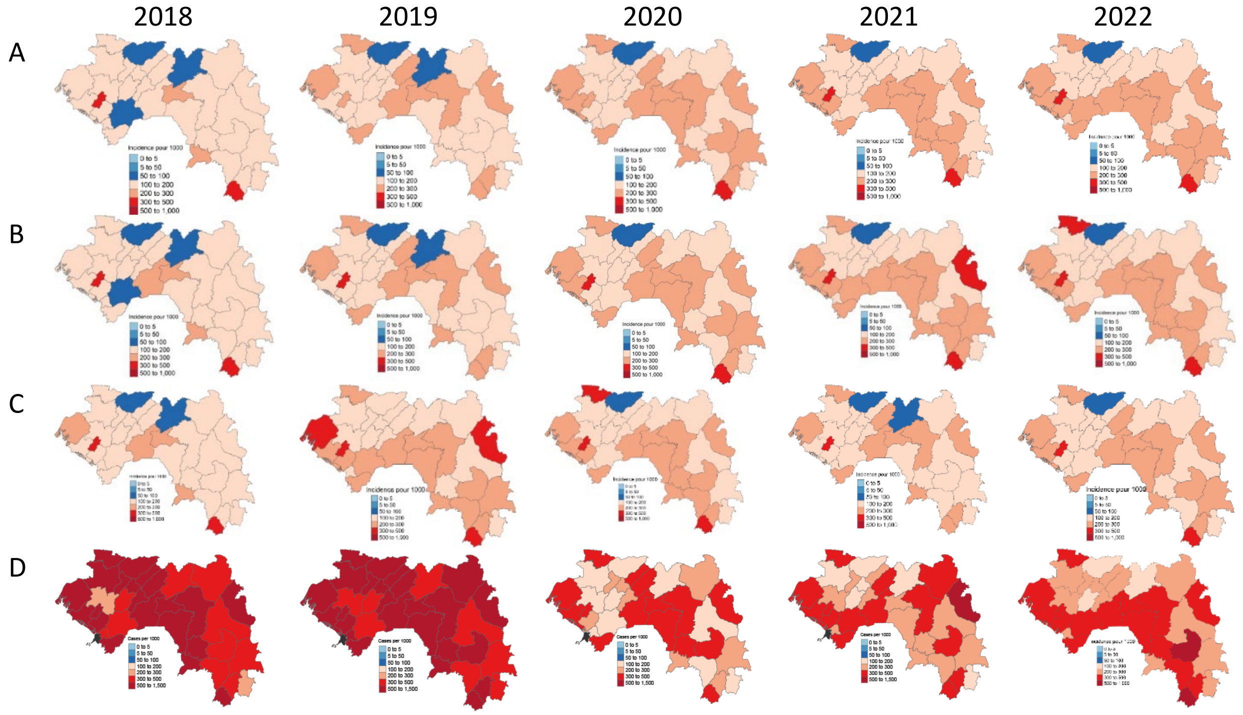

Figure S1.7: Crude and adjusted incidence for routine data from 2018-2022. A) Crude incidence per 1000. B) Incidence adjusted for testing rate. C) Incidence adjusted for testing rate and reporting rate. D) Incidence adjusted for testing rate, reporting rate, and treatment-seeking rate.

#### Adjustment 3 without dividing non-treatment-seeking by 2

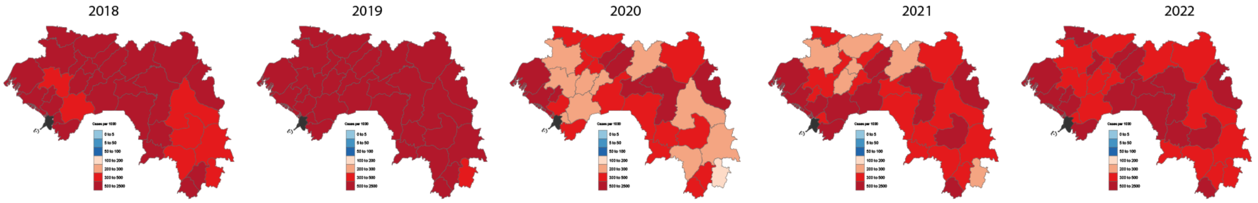

Figure S1.8: Incidence adjusted for testing rate, reporting rate, and treatment-seeking rate, without dividing the non-treatment-seeking rate by 2, for 2018-2022.

#### Parasite prevalence for 2018-2020

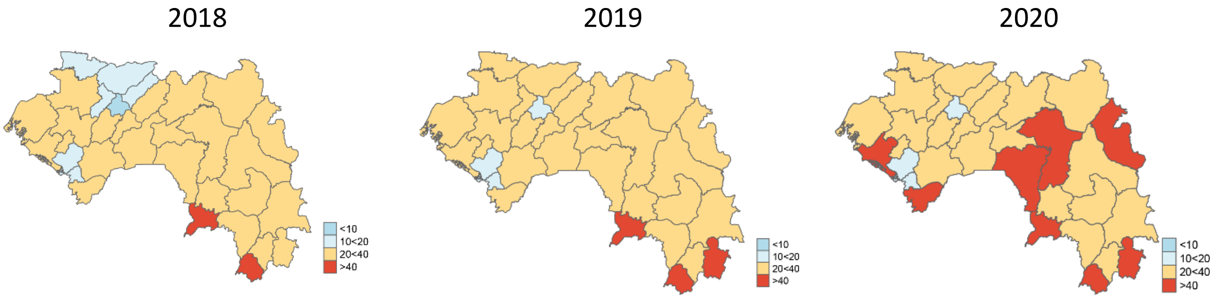

Figure S1.9: District-level estimates of parasite prevalence in children 2-10 years of age.

### Interventions

#### Therapeutic efficacy of ACTs

A literature review was carried out to examine the results of studies on the therapeutic efficacy of antimalarial drugs in Guinea. Beavogui et *al*. had carried out a study at 2 sites (Maférinya and Labé) with the aim of focusing on the efficacy and safety of artesunate-amodiaquine (ASAQ) and artemether-lumefantrine (AL). They concluded that the efficacy at day 28 of ASAQ and AL was between 99-100%.

#### Quality of care

A preliminary assessment of the main indicators of quality of care was carried out using routine data from the health districts. Quality was estimated separately for uncomplicated and severe malaria from 2018 to 2022. For uncomplicated malaria, the following 3 indicators were estimated: the testing rate, the proportion of suspected cases that were presumed, and the treatment rate (Figure S1.10). The testing rate was defined as the fraction of suspected cases that were tested. Presumed cases were divided by suspected cases to estimate the % of malaria cases that were not confirmed. The treatment rate was defined as treated cases divided by confirmed cases.

For the quality of management of severe malaria, the proportion of deaths due to malaria was reviewed (Figure S1.11). This proportion often exceeded 100%, suggesting major issues with data quality. The proportion of deaths due to malaria declined between 2018 and 2021, which could be due to improved data quality.

Overall, the quality of these data was judged insufficient for rigorous use to assess the quality of care in Guinea.

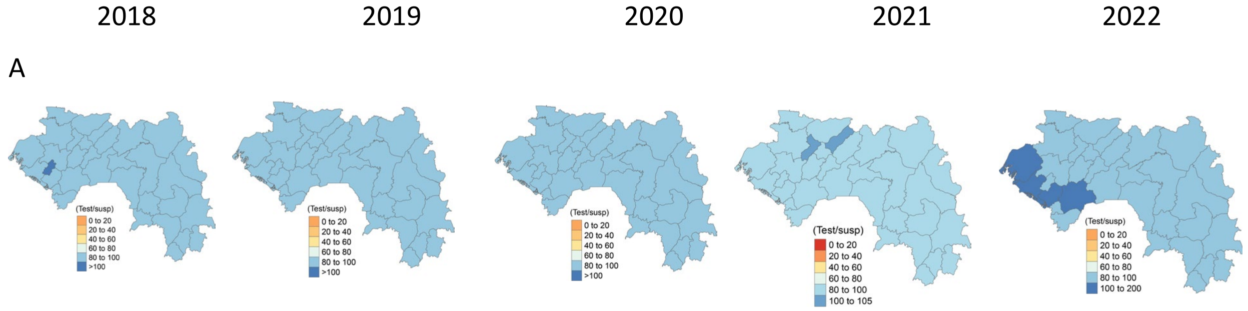

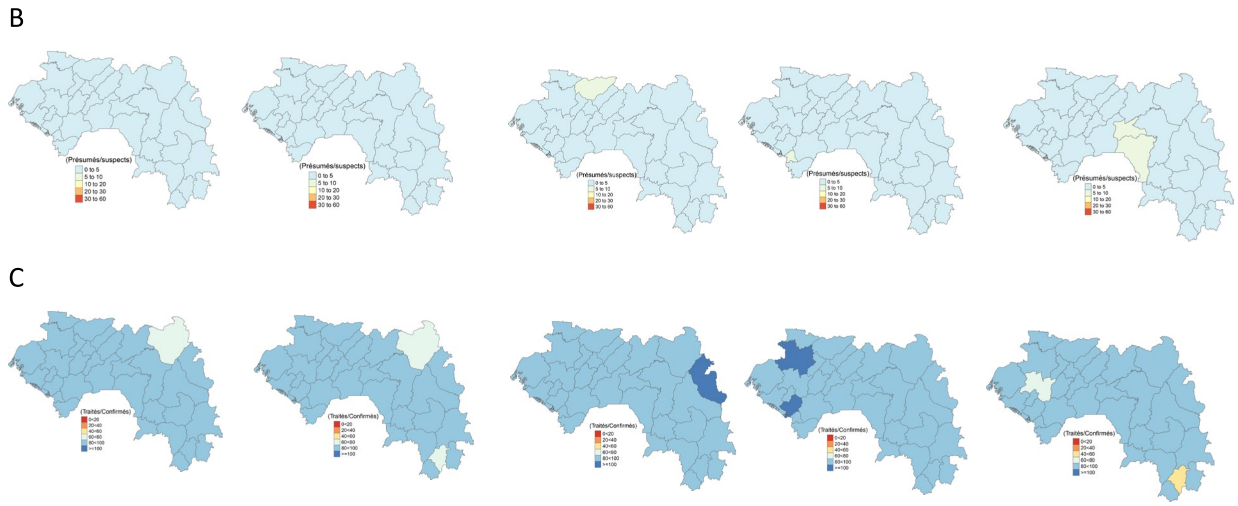

Figure S1.10. Quality of care for uncomplicated malaria. A) testing rate ; B) proportion of suspected cases that are presumed ; C) treatment rate.

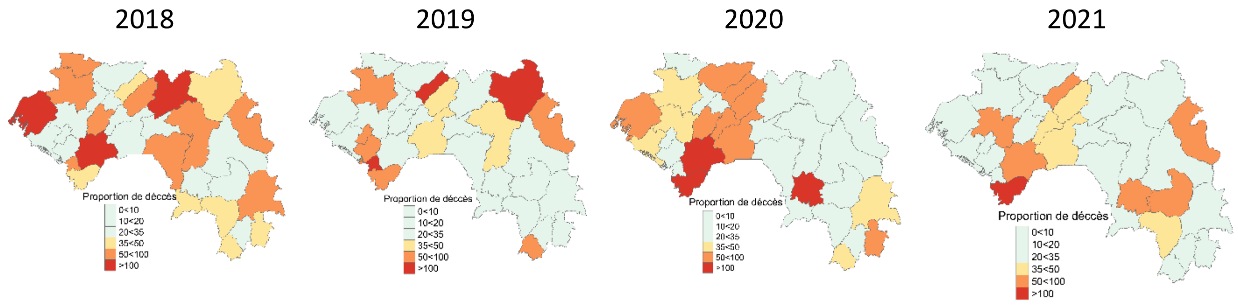

Figure S1.11. Quality of care for severe malaria: proportion of deaths due to malaria.

#### ITN access and usage

The main vector control tool used in Guinea is long-acting insecticide-treated mosquito nets (LLINs), which are distributed through mass campaigns and routinely during prenatal consultations and the Expanded Program on Immunization (EPI).

Current coverage and use of LLINs was measured at regional level through the 2021 MIS. Estimates at district level were obtained using the following procedure: given the GAPSS sampling design, an estimation function for estimates generated by complex surveys was used to estimate usage at the DS level. This analysis revealed that access and use at district level is not high (Figure S1.12).

A recent analysis of possession, access and use of insecticide-treated nets from the 2018 Demographic and Health Survey (Diallo 2023) revealed that possession was strongly influenced by household size and socio-economic factors, and that better allocation strategies could improve access. However, risk factors for possession varied from region to region. Use among those with access to nets was associated with age and socioeconomic factors, and risk factors also varied by region. These results suggest that strategies to increase ownership and use could benefit from region-specific adaptation.

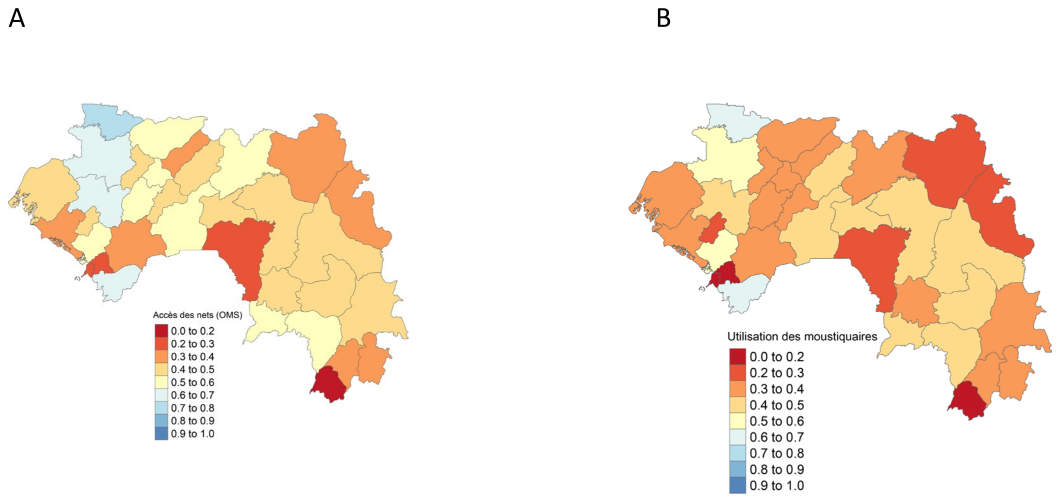

Figure S1.12. Access (A) and use (B) of LLINs in children under 5, according to Guinea's 2021 MIS.

#### Historical SMC

The number of health districts receiving SMC in Guinea has increased, with 17 districts receiving 4 cycles of SMC in 2022.

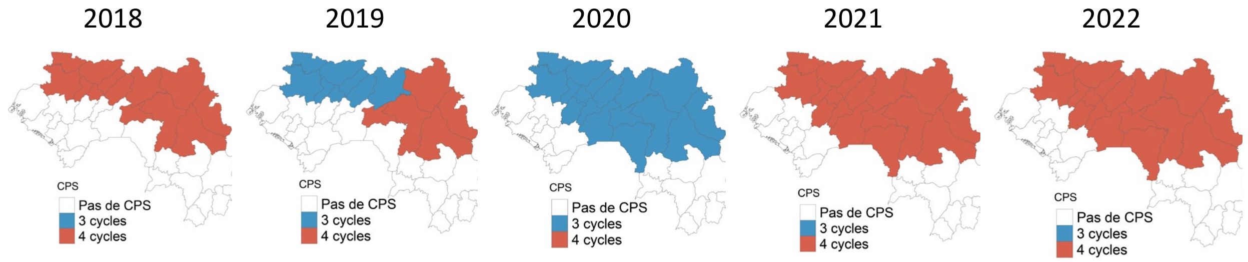

Figure S1.13. Districts receiving SMC by number of cycles, 2018-2022.

#### Case and rainfall seasonality

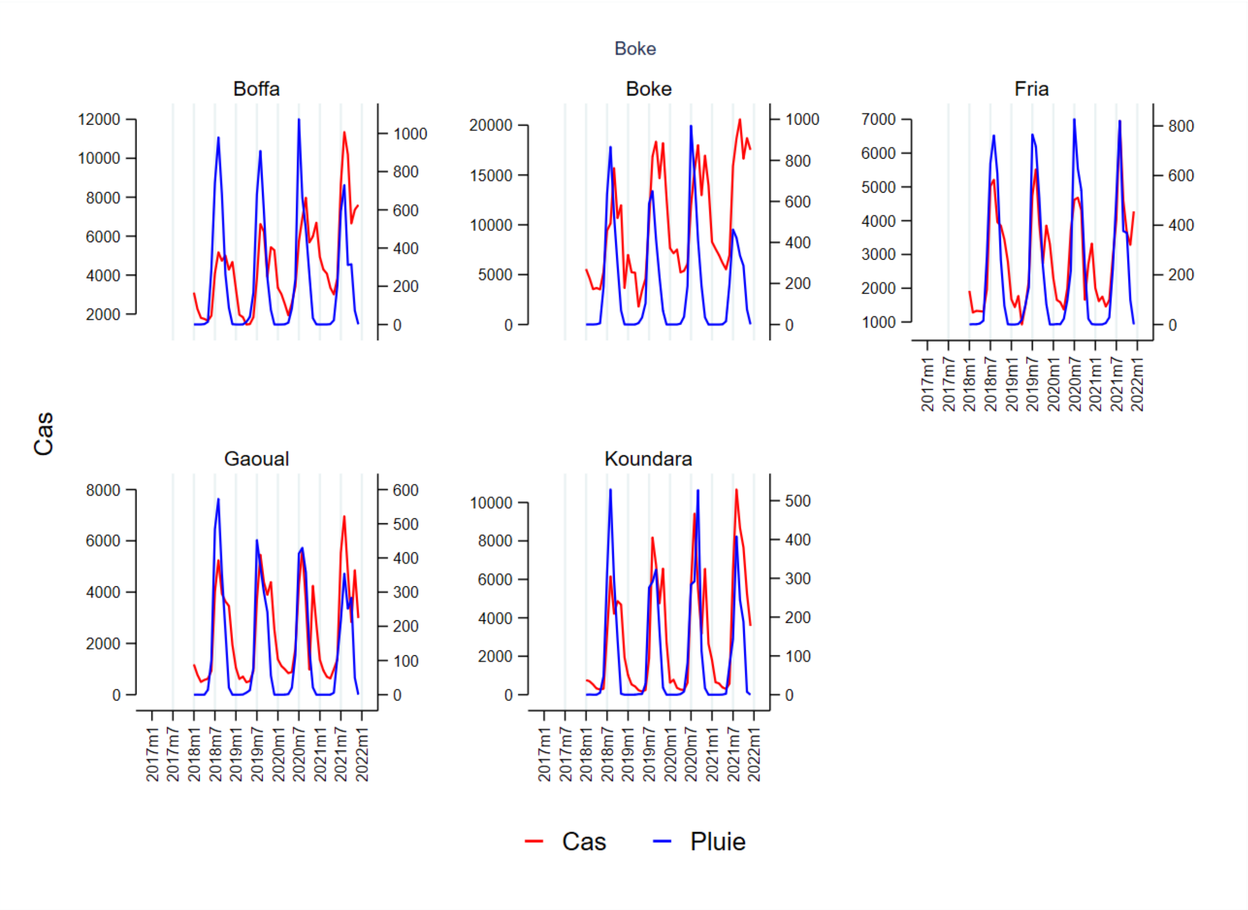

Figure S1.14. Monthly malaria cases (red) and rainfall (blue) in the region of Boké.

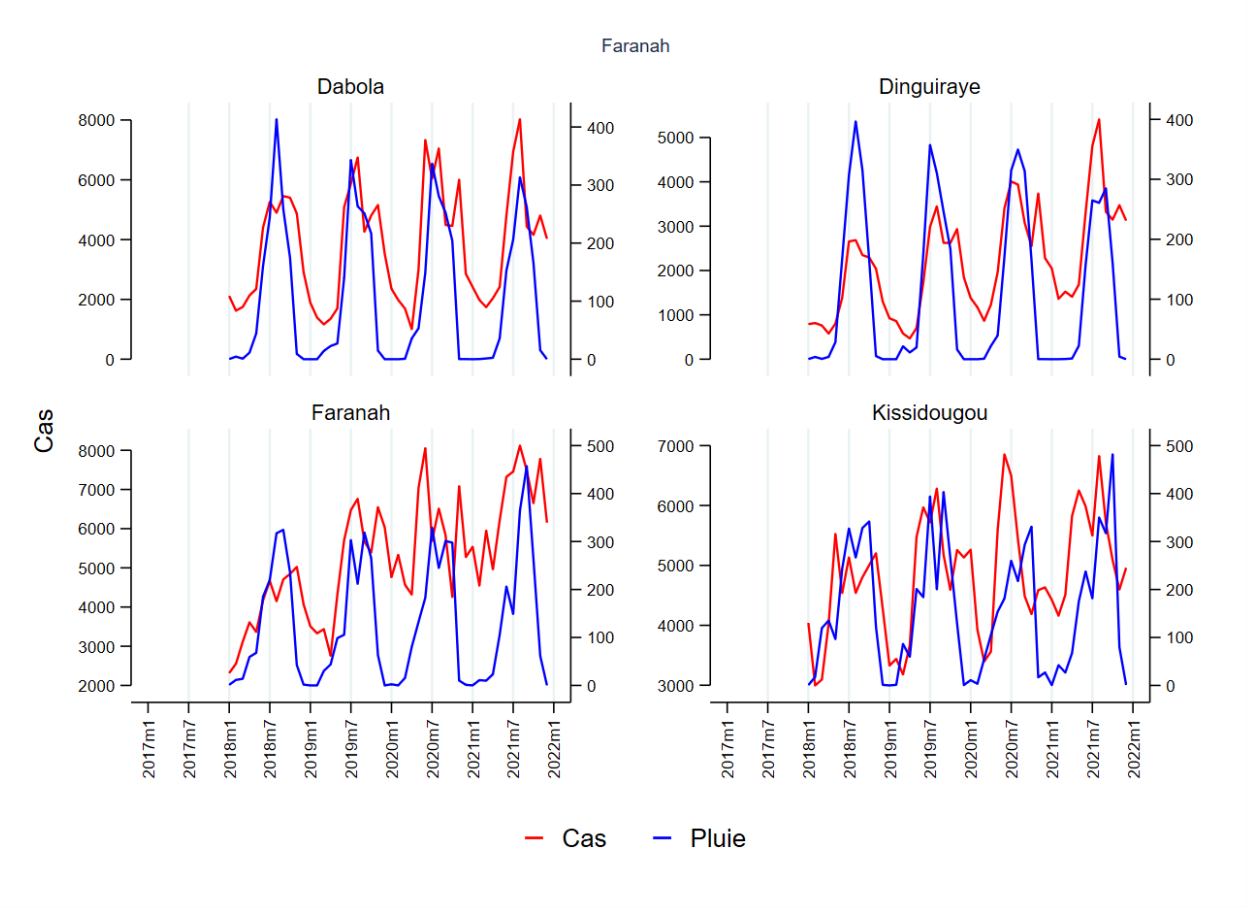

Figure S1.15. Monthly malaria cases (red) and rainfall (blue) in the region of Faranah.

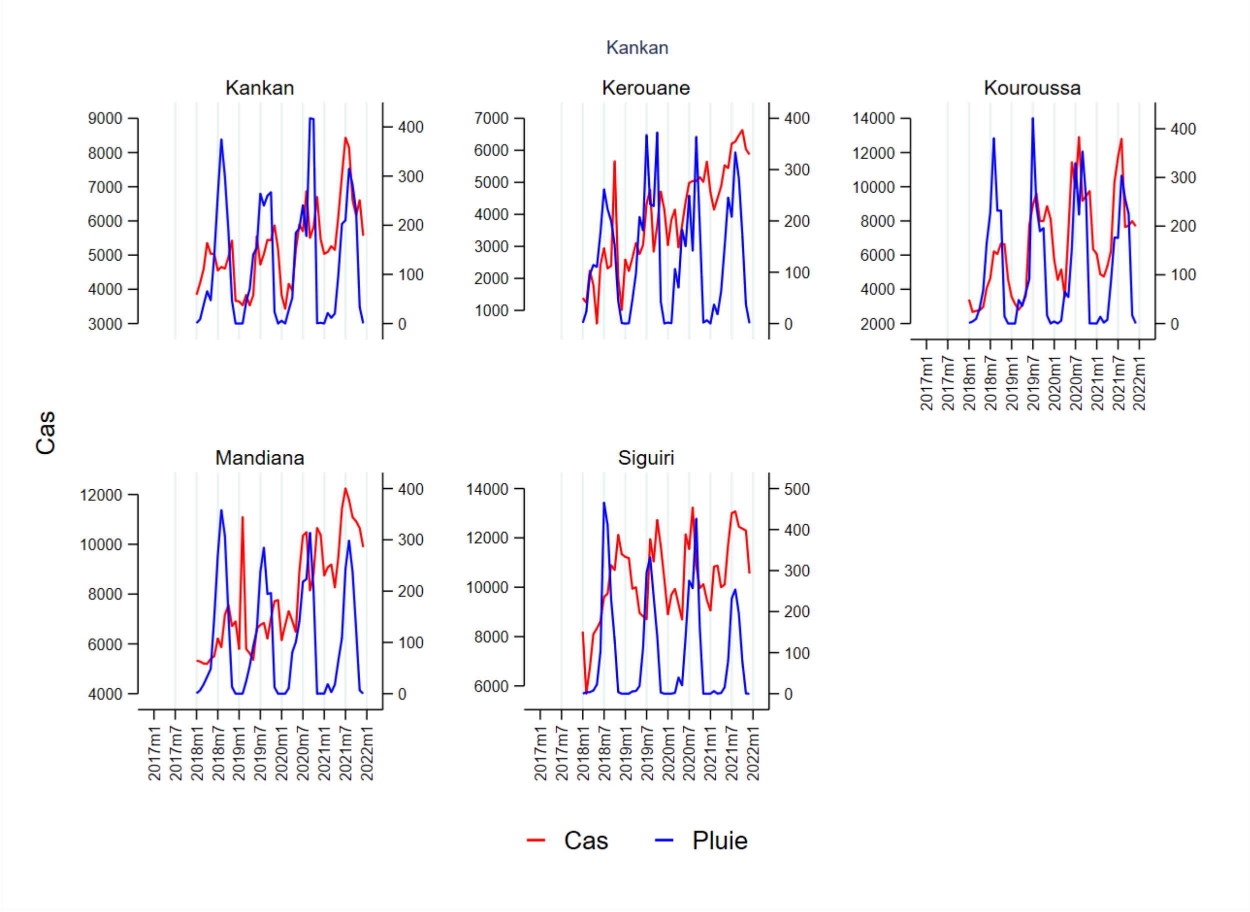

Figure S1.16. Monthly malaria cases (red) and rainfall (blue) in the region of Kankan.

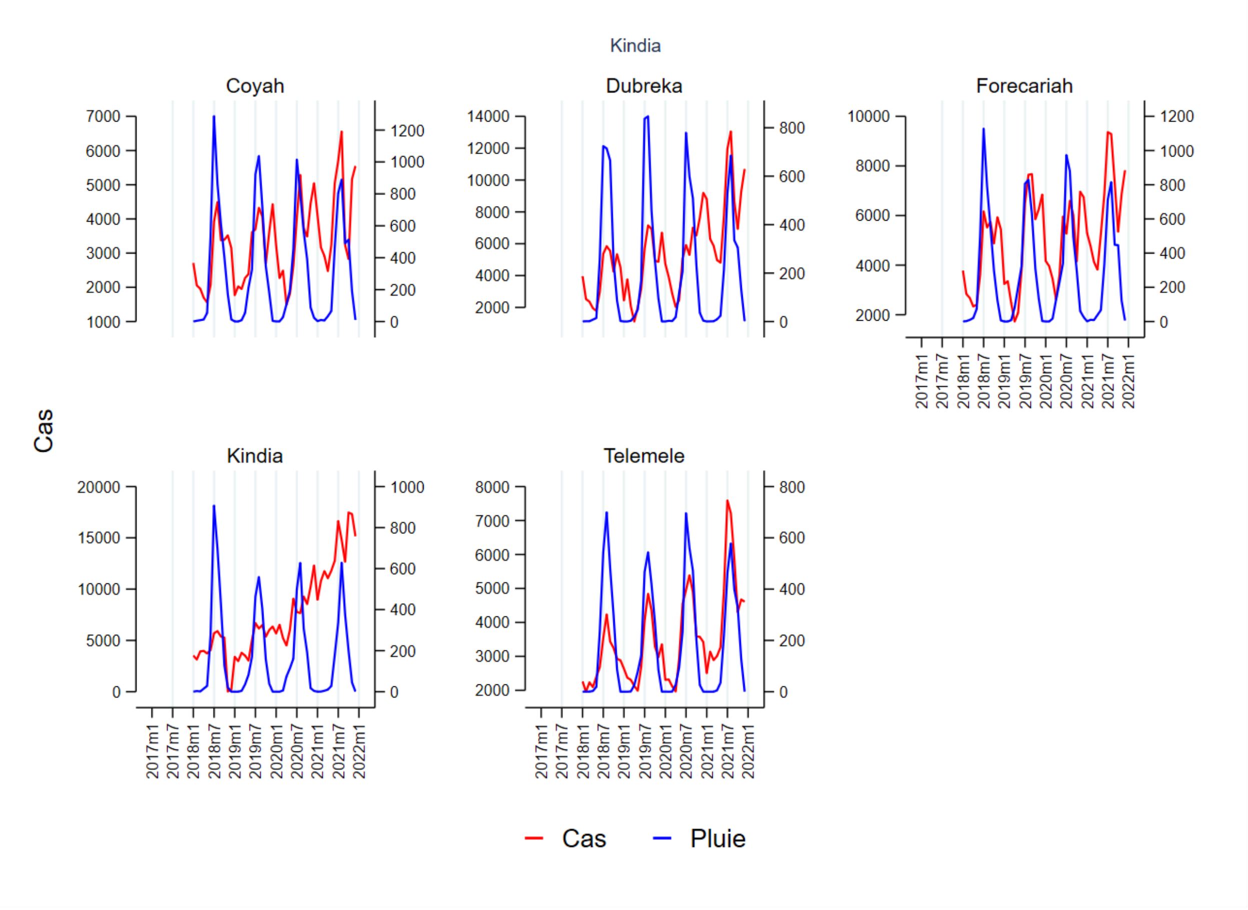

Figure S1.17. Monthly malaria cases (red) and rainfall (blue) in the region of Kindia.

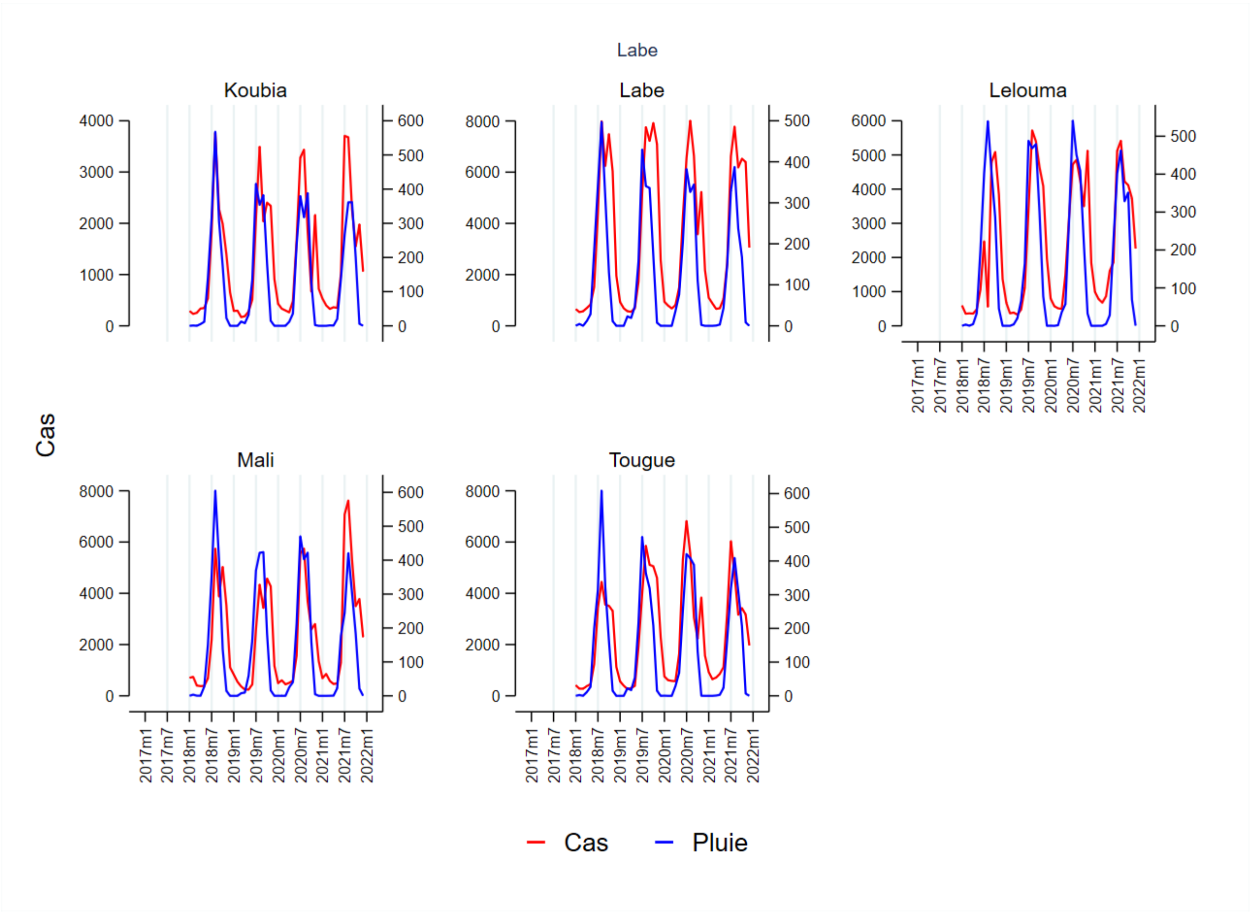

Figure S1.18. Monthly malaria cases (red) and rainfall (blue) in the region of Labé.

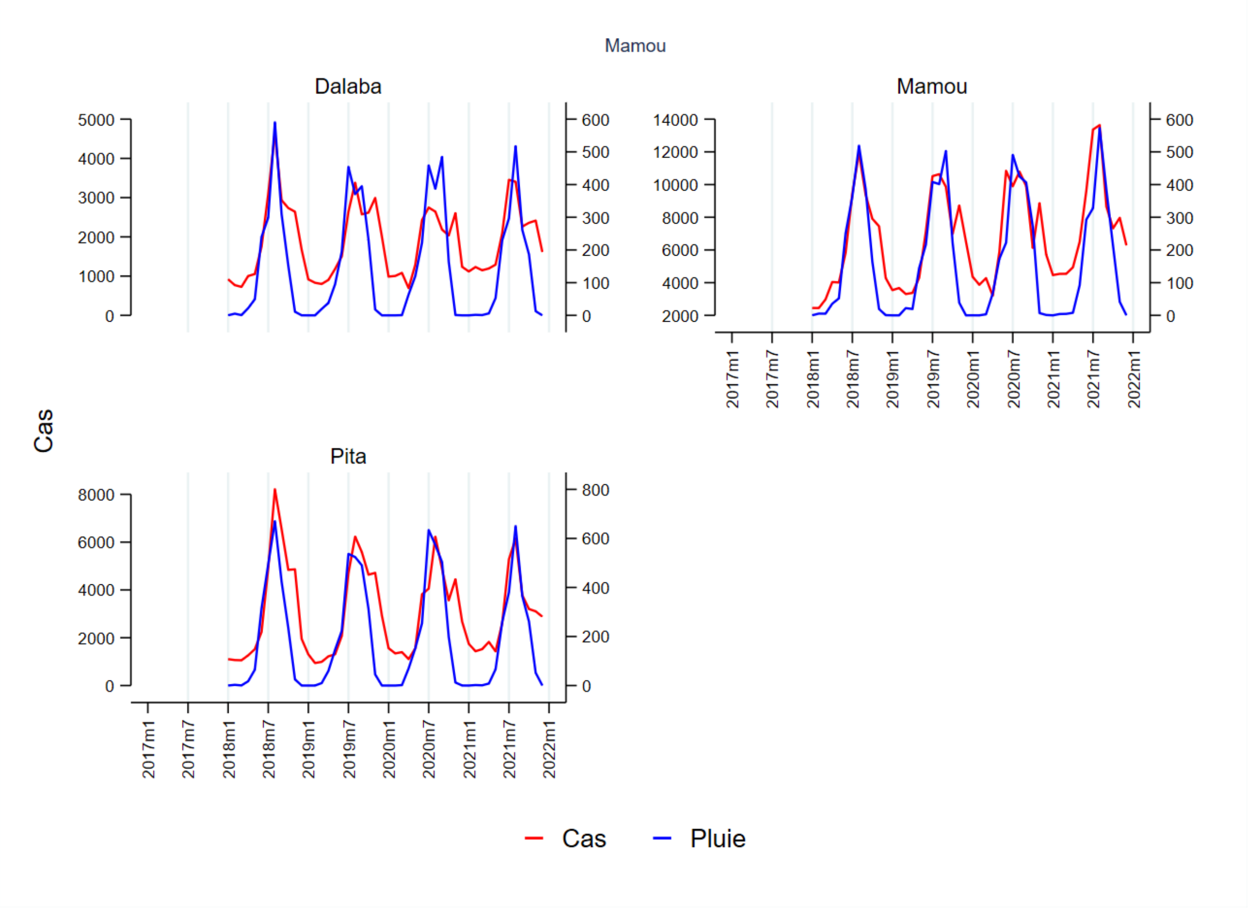

Figure S1.19. Monthly malaria cases (red) and rainfall (blue) in the region of Mamou.

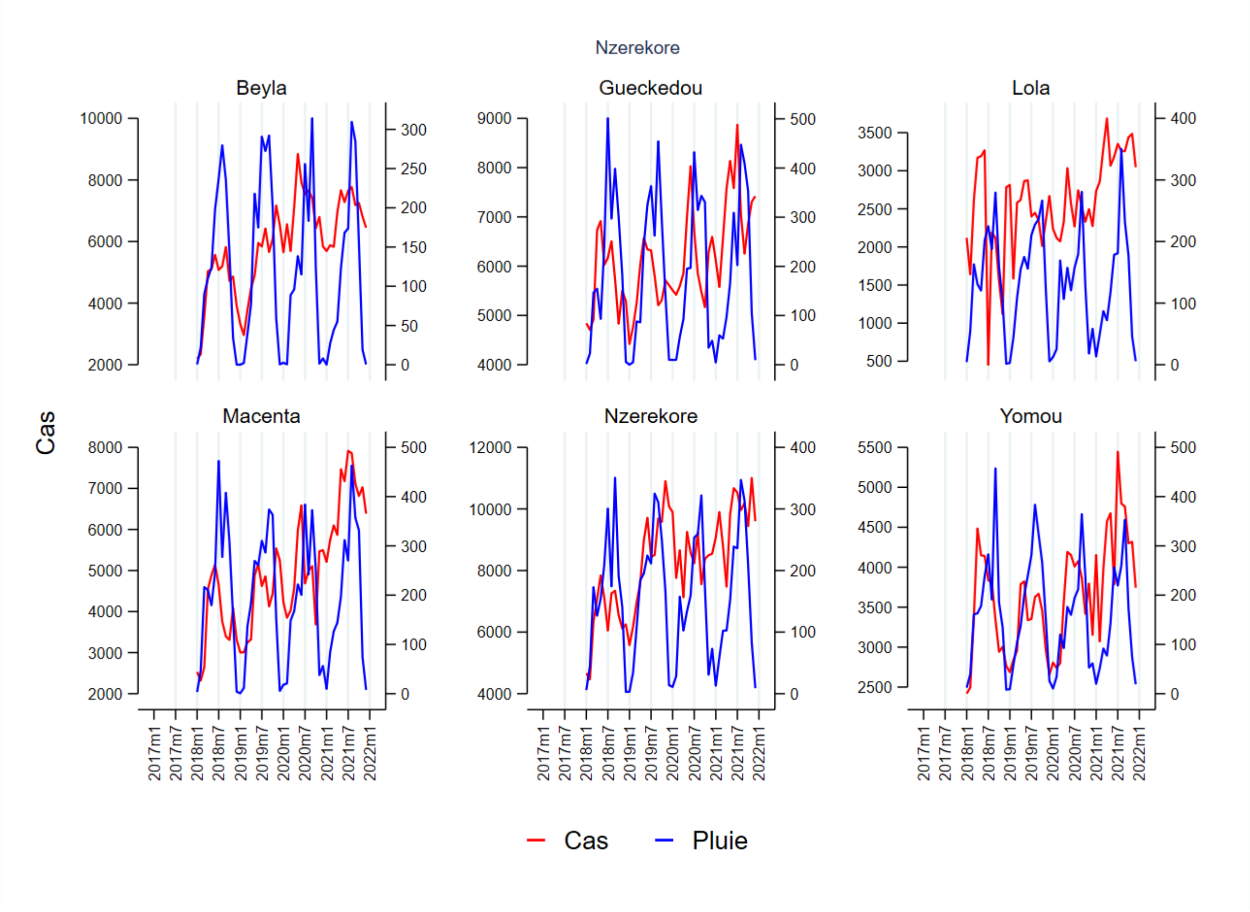

Figure S1.20. Monthly malaria cases (red) and rainfall (blue) in the region of N'zérékoré.

#### Case seasonality and the SMC calendar

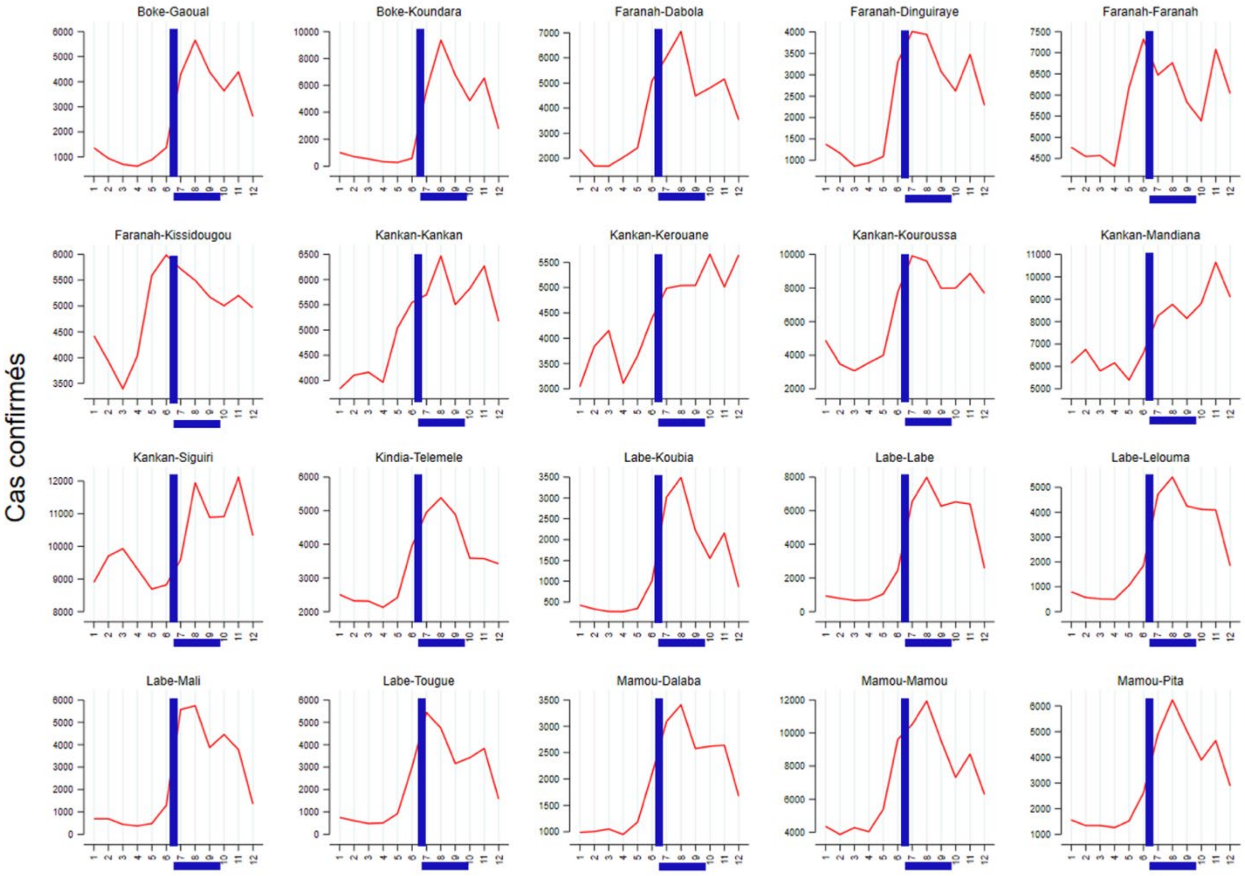

Figure S.21. Timing of confirmed cases in relation to SMC in July and October (blue horizontal bar), by district, according to routine data

### References

Beavogui AH, Camara A, Delamou A, Diallo MS, Doumbouya A, Kourouma K, Bouedouno P, Guilavogui T, dos Santos Souza S, Kelley J, Talundzic E. Efficacy and safety of artesunate–amodiaquine and artemether–lumefantrine and prevalence of molecular markers associated with resistance, Guinea: an open-label two-arm randomised controlled trial. Malaria Journal. 2020 Dec;19(1):1-9.

Diallo OO, Ozodiegwu ID, Camara A, Galatas B, Gerardin J. Factors associated with the ownership and use of insecticide-treated nets in Guinea: an analysis of the 2018 Demographic and Health Survey. Malaria Journal. 2023 Dec;22(1):1-3.

World Health Organization. World Malaria Report 2023. World Health Organization: Geneva, 2023.
