## Supplementary material for "Subnational tailoring of malaria interventions to prioritize the malaria response in Guinea": Supp File 2: Mathematical Modeling

Supplementary file 2: Mathematical modeling methods and results

#### Overview

We use EMOD v2.20, an agent-based model commonly used for modeling *Plasmodium falciparum* (*Pf*) transmission and disease, to simulate impact of malaria intervention across a range of varying scenarios (Bershteyn 2018, IDM EMOD 2023). The model includes a compartmental vector model that captures the mosquitoes' behavior and climate-dependent population dynamics; individual humans with asexual and sexual stage parasite infections, uncomplicated and severe symptoms, and acquired immunity; and all standard and many novel malaria interventions including treatment with artemisinin-based combination therapy (ACTs), chemoprevention with sulfadoxine-pyrimethamine (SP) with or without amodiaquine, pre-erythrocytic vaccines, indoor residual spray, and insecticide-treated bednets.

Key model parameters related to parasite dynamics, exposure-dependent infections and symptoms, and acquisition of immunity were previously calibrated to data from nine reference sites across sub-Saharan Africa (McCarthy 2015, Gerardin 2015b, Selvaraj 2018). Intervention parameters, such as ACTs and RTSS, were calibrated to clinical trial data in previous studies (details below).

Our modeling framework is designed to adapt to local contexts within countries and provide future projections of malaria and intervention impact. We simulate each health district in Guinea as a closed population, without considering movement between districts, and aggregate the outcomes to provide national estimates.

#### Basic model setup

Three vector species were included in the model: *Anopheles arabiensis, Anopheles funestus,* and *Anopheles coluzzii*. Vector biting behavior (indoor biting rate and anthropophily), were obtained from West African research studies, and we assumed this vector behavior to be the same in Guinea as in other West African countries (Table 1). For each vector species we assumed the same seasonality (see later sections for determination of seasonality). We based relative vector abundance on geostatistical model estimates from Sinka et al. (Sinka, 2016).

For modeling purposes, we initialized each district with a population of 1000 humans. Birth rate was fixed at 0.1 per day, similar to Guinea's crude birth rate of 35.9 per 1000 person per year in 2018 (The World Bank, 2024) and death rate was set to the same value, such that the population size was stable. The initial age structure of the population was the equilibrium structure from the birth and death rates.

The simulation results were scaled to the district population to obtain district-level estimates.

Table 1: Vector parameters

| Parameters | Assumed values | Data source |
|---|---|---|
| Vector species | *Anopheles arabiensis, Anopheles funestus,* and *Anopheles coluzzii* | Sinka et al. 2016 |
| Vector relative abundance | Varies by district | Sinka et al. 2016 |
| Vector bionomics<br>*An. arabiensis*<br>Indoor feeding rate<br>Anthropophily<br>*An. funestus*<br>Indoor feeding rate<br>Anthropophily<br>*An. coluzzi*<br>Indoor feeding rate<br>Anthropophily | <br><br>0.50<br>0.88<br><br>0.86<br>0.50<br><br>0.90<br>0.74 | Guelbeogo et al. 2014.<br>Nguyen et al., 2017<br>Hien et al., 2020<br>Note: we chose 0.50 for parameters with wide-ranging numbers across multiple literatures. |
| Seasonality of vector abundance | Varies by archetypes | Calibrated to routine data |

#### Intervention parameterization

Multiple sources of data were used to inform parameterization of interventions (Table 2). Key data sources included the Demographic and Health Surveys (DHS) and Malaria Indicator Surveys (MIS), which were available for 2012, 2018 and 2021 (Institut national de la Statistique (INS) [Guinée], et ICF, 2012, 2018 and 2021).

Table 2: Model intervention parameters and sources

| Type | Parameters | Resolution | Data source |
|---|---|---|---|
| Case management | Fraction of uncomplicated malaria cases that are treated | district | Fever care-seeking rate and ACT usage within 2 weeks prior to survey in children under the age of 5 years from DHS/MIS data. Assumed to be the 50% less for adults. |
| | Fraction of severe malaria cases that are treated | district | Changing in relation to uncomplicated case management |

| | | | |
|---|---|---|---|
| | Efficacy | national | Previously calibrated to clinical trial data for AL (Gerardin 2015a) |
| Insecticide-treated net | Usage | district | Overall usage based on MAP estimates, age group and seasonal usage based on DHS/MIS data |
| | Retention | national | Physical attrition of bed net described by net survival based on estimates from MAP (Bertozzi-Villa 2021) |
| | Waning of insecticides | default | Previously calibrated (Eckhoff 2013) |
| | Initial efficacy given resistance | district | Based on resistance estimates from (Hancock 2020) and statistical model between permethrin and experimental hut trial data (Nash 2021) to inform killing and blocking rates of pyrethroids nets, calibrated to control group of Toe et al. 2018. IG2 efficacy was informed by relationship between standard and PBO nets (Sherrard Smith, 2022) as well as experimental hut studies conducted in West African countries (Bayili 2017, N'Guessan 2016, Ketoh 2018, Toe 2018) |
| | Insecticide/ ITN type | district | Programme National de Lutte Contre le Paludisme (PNLP) |
| PMC | Coverage | region | Proportion of children getting an effective dose of SP at specified ages. Based on reported vaccination coverage from 2018 Guinea DHS adjusted for an EPI to PMC scaling factor from an implementation study in Sierra Leone (Lahuerta 2021) per region |
| | Efficacy | national | Previously parameterized to Ghana IPTi trial (Runge 2023) |
| SMC | Coverage | district | Estimated from Loua report using mean cycle received per district |
| | Efficacy | national | Parameterized to SMC trials in Burkina Faso (Zongo 2015, Chandramohan 2020) |
| Malaria vaccine (RTS,S) | Coverage | region | Based on 2018 Guinea DHS reported measles coverage per region |
| | Efficacy | national | Previously calibrated to Phase 3 trial data (Penny 2016). |

#### Case management

All modeled treatment of symptomatic malaria was assumed to occur with the ACT artemether-lumefantrine (Gerardin 2015a) and no ACT resistance. Case management began in the model in 2005 when Guinea established a policy of ACTs for first-line treatment (Cherif 2021). A successful case management is defined as a febrile individual with parasitemia seeking medical care and being effectively treated with an ACT-based treatment.

In the model, all malaria cases are treated with artemether-lumefantrine within 3 days of clinical disease onset, and within 2 days of severe disease onset.

##### Uncomplicated malaria

We estimated the case management coverage using DHS/MIS data. We assumed that the coverage is the fever care-seeking rate multiplied by the ACT usage rate for malaria treatment. The care-seeking rate of a household cluster was calculated as the fraction of children under the age of 5 years who sought care when they had a febrile illness in the two weeks prior to the survey. A Bayesian geospatial model was used to interpolate the cluster-level estimates of care-seeking rate for febrile children under 5 years of age to the entire country at a 2 x 2km resolution. We did not use any spatial covariates for the model as they are not predictive of the response (Nguyen 2023). The Bayesian geospatial model was run using INLA method in R (Rue 2009, Martins 2013). All pixels within a health district were aggregated, weighted by the total population in each pixel, to obtain district-level mean and 95% intervals (Fig 1). The population rasters were obtained from WorldPop for the corresponding years of the DHS/MIS data (WorldPop 2018, Lloyd 2019).

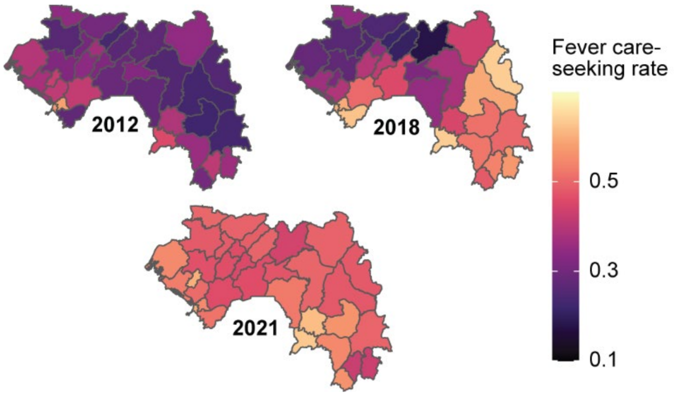

*Fig 1: Fever care-seeking rates for each district, estimated using DHS/MIS datasets and a Bayesian geospatial model.*

The ACT usage rate was calculated as the proportion of children who were treated with any form of antimalarial for their febrile illness who received ACT. The ACT usage could be estimated on either the national or the regional level. The PNLP was presented with both options (Fig 2) for ACT usage and their downstream effect in the case management coverage assumptions (Fig 3, Fig 4). The PNLP chose the first option (Fig 2A) as the district and even regional-level estimates were deemed highly uncertain due to the small sample sizes of febrile children who sought care.

For years without any DHS/MIS, we used linear interpolation to obtain the case management coverage assuming a coverage of 0 in 2005 and the same as in 2021 from 2022 onwards (Fig 4). The treatment rate for individuals over 5 years was assumed to be 50% of the treatment rate for children under 5 years.

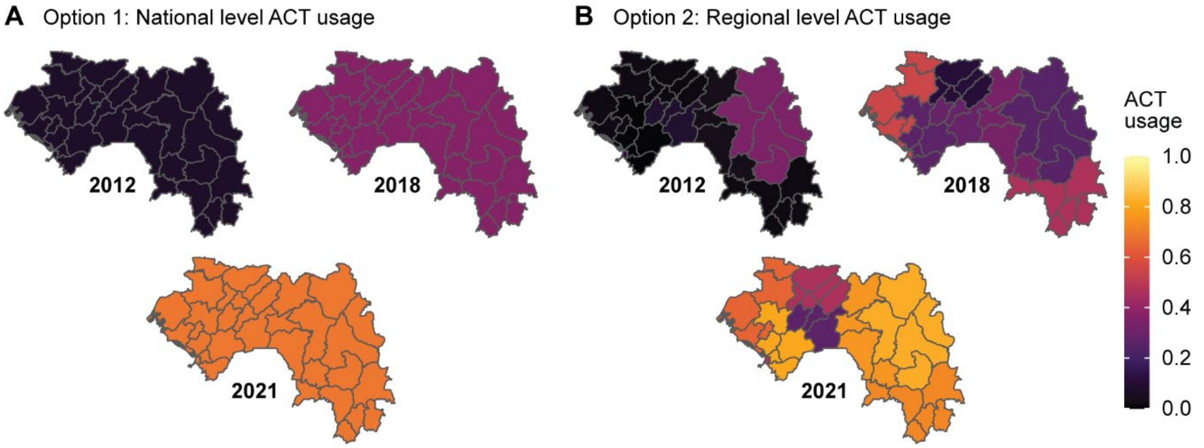

*Fig 2: PNLP was presented with two options: (A) to estimate ACT usage at national level and assume that all districts have the same ACT usage, or (B) to estimate ACT usage at regional level, whereby districts within a region share the same ACT usage.*

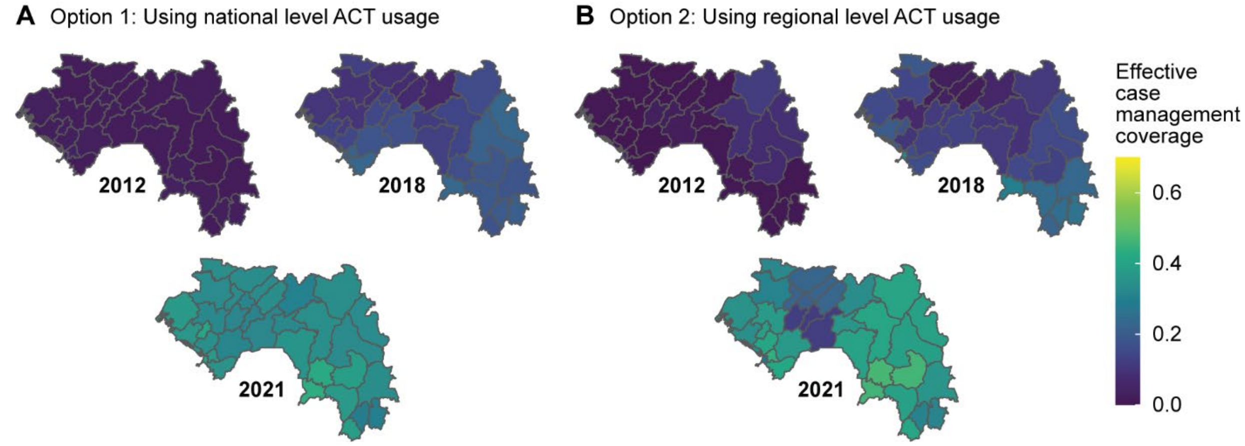

*Fig 3: Mean coverage of effective case management for each district, calculated using ACT usage at (A) national level, or at (B) regional level. They are estimated from 2012, 2018 and 2021 DHS/MIS data.*

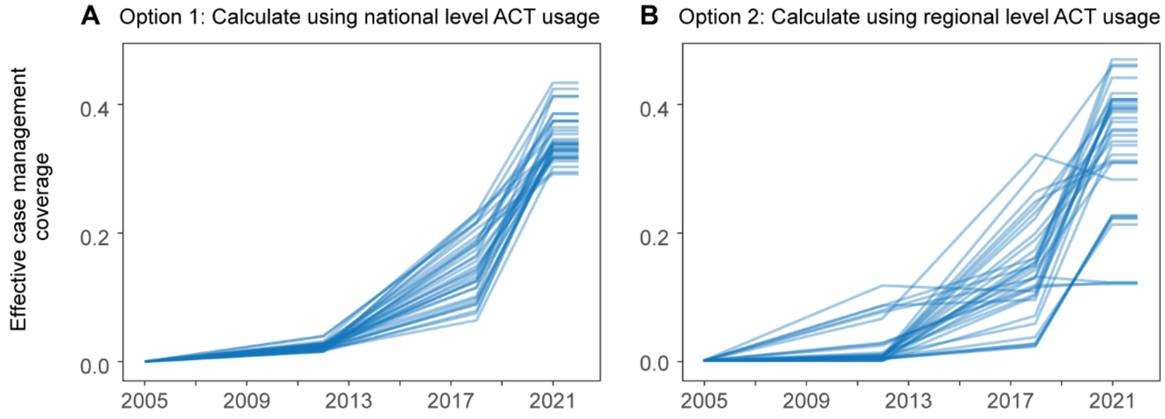

*Fig 4: Mean coverage of effective case management from 2005 to 2022, calculated using ACT usage at (A) national level, or at (B) regional level. Coverages were interpolated in years without DHS/MIS data. Each line represents one district.*

##### Severe malaria

The treatment rate for severe malaria for all ages was assumed to be the greater of either 60%, or a relative 60% higher than the treatment rate for children under 5 years of age with uncomplicated malaria.

The model assumed a case-fatality rate of 9.7% for treated severe malaria cases for all ages, and 33% for untreated severe malaria cases (von Seidlein 2012).

##### Non-malarial fevers

Given that a substantial fraction of malaria infections can come from RDT-positive individuals seeking care for a non-malaria illness (Dalrymple 2019), we assumed that each day a random 0.38% of the entire population would get tested for malaria and treated if they tested positive. This is equivalent to about 1.4 non-malaria fever episodes per person per year.

#### Insecticide-treated nets (ITNs)

##### ITN coverage

Multiple data sources were used to parameterize the coverage and effect of long-lasting ITNs (Table 2). In the model, the timing of mass distribution of ITNs followed the PNLP's implementation. Mass distributions began in 2013 and occur every 3 years.

In the model, individuals who have access to a net use the net with a maximum 90% probability each night, modulated by a seasonal usage factor, until they discard the net. Seasonal usage was estimated by inferring the month effect from usage data in the 2012 and 2018 DHS using rainfall and temperature as covariates.

We use ITN usage data (fraction of people who slept under a net the night before) to inform the initial coverage of ITNs given during mass distribution events in the model.

There were two potential sources for estimating overall ITN usage for each district: (1) aggregating net usage data of all clusters from DHS/MIS within a district, or (2) aggregating net usage values of all pixels (weighted by population) within a district according to the usage maps created by MAP (Bertozzi-Villa 2021). The two options (Fig 5) were presented to the PNLP, who decided to use the second option, as MAP incorporated DHS/MIS data on a larger scale for estimation. We extracted the mean and 95% interval from MAP during the mass distribution years. The mean and 95% interval were used as the proposed distribution for the initial coverage of ITN during the year of mass distribution (See section on Setting modeled transmission intensity). Since MAP's estimation stopped at 2020, we assumed subsequent deployment of ITNs to have the same usage level as in 2020 (Fig 6).

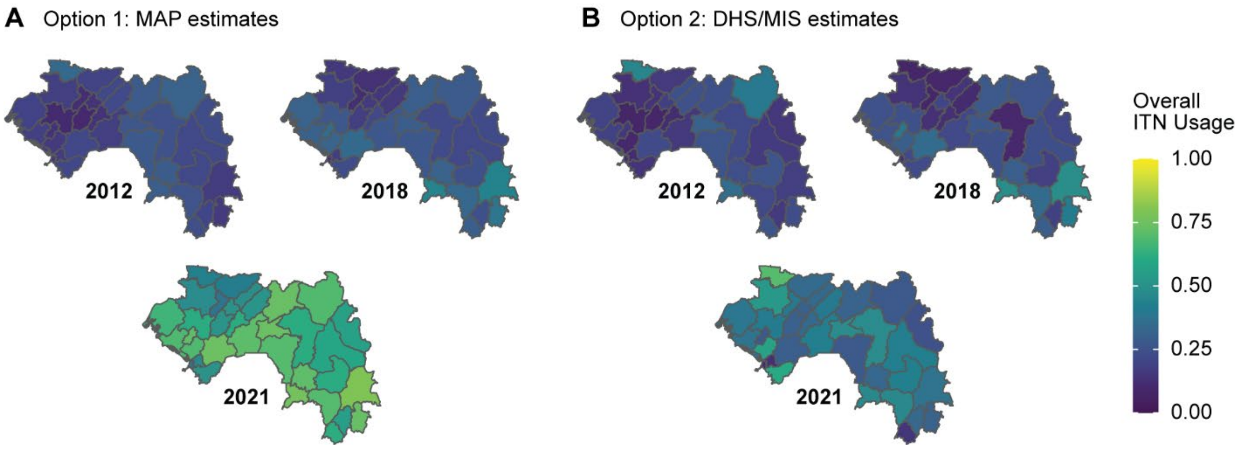

*Fig 5: The PNLP was presented with two possible sources of estimating ITN usage, using (A) MAP or (B) DHS/MIS dataset. Since MAP only estimated ITN usage up to 2020, the 2021 usage level in A is based on 2020 estimates. The years presented here are based on the DHS/MIS data availability.*

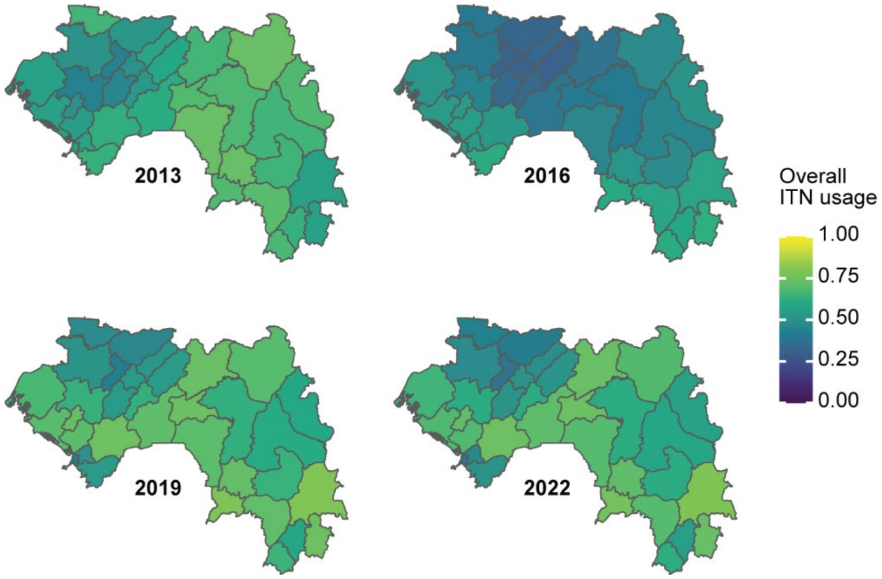

*Fig 6: Mean all-age ITN usage for each district and each ITN distribution year used in the model. Usage levels were estimated from (Bertozzi-Villa 2021).*

To account for variation in net usage by age, we adjusted the initial coverage of ITN based on age-specific ITN usage estimated from DHS/MIS. Regional estimates were used for age-specific ITN usage because DHS/MIS is powered at this level. In each region, we calculated the ITN usage by aggregating the individual response for each of these four age groups: under the age of five, five to ten years old, ten to twenty years old, and above twenty years old. Then the usages were normalized to the usage of those under the age of five (Fig 7). The initial net coverage of any age group is obtained by multiplying the ITN coverage of those under the age of five with the corresponding normalized usage. For each ITN mass distribution year, we use the age-specific ITN usage derived from the closest DHS/MIS (e.g., 2013 usage was based on DHS 2012, etc.).

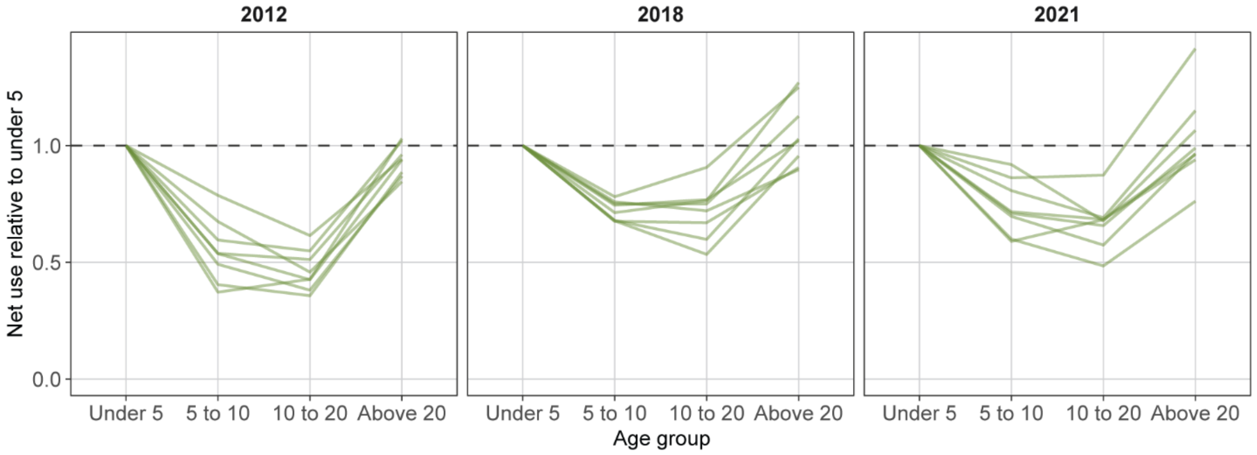

*Fig 7: Regional ITN usage level for each age group, expressed as a ratio to the corresponding usage level for age under 5 years. Each line represents a region.*

#### Net retention

In the model, the usage, and protective effects of ITN are highest at the time of deployment of nets, and the effects wane over time due to net retention, net durability, and insecticide-killing effect decay. For the net retention time, we first fitted a Weibull distribution to the Burkina retention data of the New Nets Project (NNP) Interim results report (PATH 2022). This resulted in a shape parameter ($k$) of 2.3 and a scale parameter ($L$) of 2.5, corresponding to 50% of nets retained for 2.5 years in the simulated population. In the final set of simulations, the scale parameter was modified to 2.39 years based on (Bertozzi-Villa 2021) which suggests that the median retention rate of ITNs in Guinea was 96% of that of Burkina Faso.

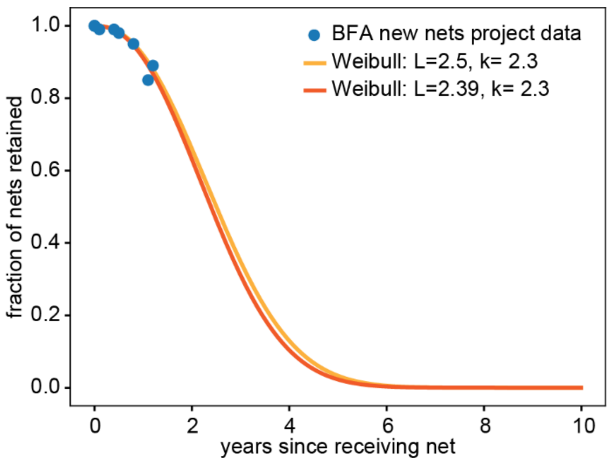

*Fig 8: Decline of ITN ownership over time used in the simulations compared to observed retention in Burkina Faso (BFA).*

#### ITN efficacy parameterization for pyrethroid nets

In EMOD, efficacy of ITNs is described by the killing and the blocking rate, that define probabilities of a mosquito to be blocked by an ITN from reaching the human host, and the subsequent probability to die after having been exposed to the insecticide. At full susceptibility of mosquitoes against pyrethroids, the initial blocking and killing of pyrethroid-treated nets have been estimated at 0.6 and 0.9 respectively (Eckhoff 2013), based on calibration to early ITN trials summarized in a 2004 Cochrane review (Lengeler 2004). To account for pyrethroid resistance in the effect sizes of pyrethroids and dual-insecticide-nets, an adjustment method had been implemented in the modeling analysis for Nigeria (Ozodiegwu 2023). In the present study for Guinea, we modified the previous ITN efficacy adjustment methods using newer studies on ITN impact and a revised approach as described below.

In 2021, Nash and colleagues published a systematic review on the impact of ITNs that included statistical relationships between observed mosquito mortality in experimental hut trials (EHT) and entomological indicators of ITN impact, such as successful blood feeding rates and exit rates (Nash 2021). From these, the authors derived mosquitoes kill rates, deterrence, hut exit rates with or without being blood-fed for a range of bioassay survival rates. We used these rates to determine ITN efficacy for EMOD at given resistance levels across geographical areas, assuming that the relationships between bioassay survival rates and EMOD parameters of killing and blocking are the same functional form as the Nash et al. relationships between bioassay survival rates and kill rates and one minus blood fed rates.

Since EMOD mosquitoes can only die when first being blocked, the Nash et al. category 'blood fed and dead' was completely merged into the "blood fed and alive" category (Fig 9).

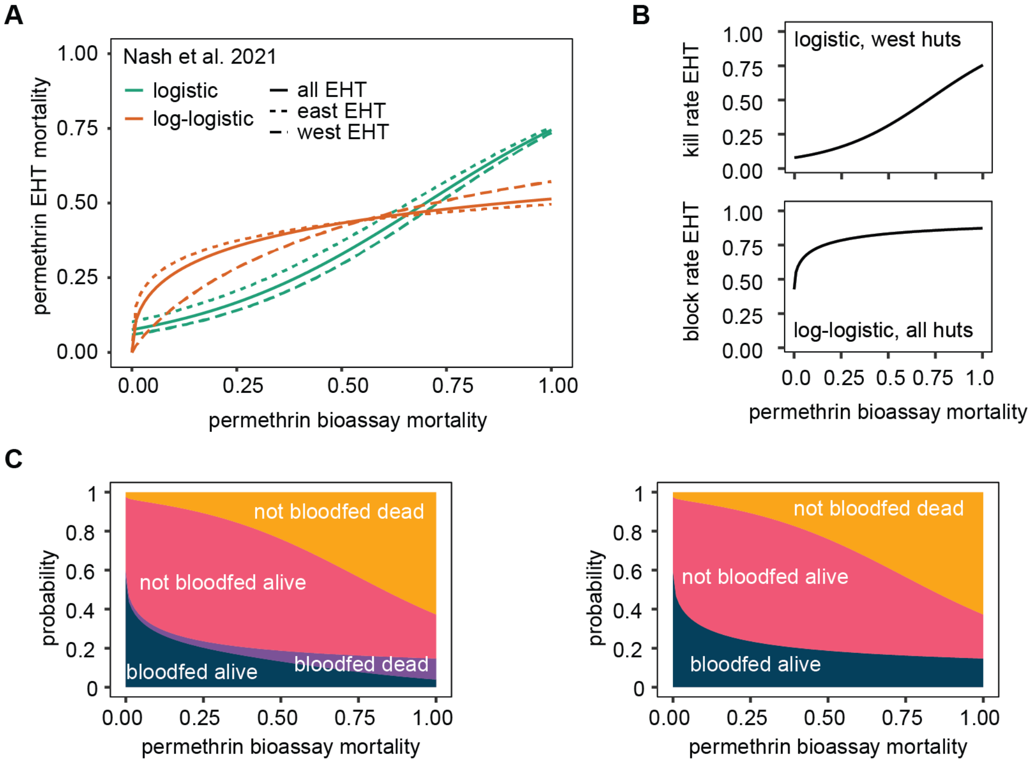

*Fig 9: A) Relationships between bioassay and experimental hut (EHT) mortality reproduced from Nash et al 2021. B) Relationship between bioassay mortality and EHT blood feeding or mortality rates. C) Probability of mosquitoes being bloodfed and alive, bloodfed and dead, or not bloodfed and alive or not bloodfed and dead by bioassay mortality, in EHT's or in EMOD. Initial script in R with statistical functions obtained from Ozodiegwu 2023 (Monique Ambrose, IDM personal communication) and modified for present analysis.*

We began with the prior assumption that for each level of bioassay mortality, the EMOD kill rate parameter is the same value as the Nash et al. estimate of the kill rate, and that the EMOD block rate parameter should be the same value as one minus the Nash et al. estimate of the total blood fed rate.

These prior assumptions were subsequently adjusted so that for a known level of local bioassay mortality, the epidemiological impact of pyrethroid net distribution matched an observed epidemiological impact from clinical trial.

Two data points were used to anchor the adjustment: 1) the no resistance context from Eckhoff 2013, where 100% bioassay mortality was associated with EMOD kill rate of 0.6 and EMOD block rate of 0.9; and 2) a high resistance context: the control arm of an Olyset Duo trial conducted in Banfora District in the Cascades Region of Burkina Faso, with 15.2% bioassay mortality (sample size weighted average in the Cascades Region from geospatial estimates in Hancock 2020) and 12.7% reduction in clinical incidence after deployment of pyrethroid nets (Tiono 2018).

We used the Nash et al. logistic (west Africa) kill rate model and log-logistic blocking rate model to set the prior at 15.2% bioassay mortality and to set the functional form for the relationship between parameter value and bioassay mortality (Table 4), as this combination was most successful at being able to capture both our anchor observations. Other combinations systematically under- or over-estimated incidence reductions.

**Table 4:** Estimated crude ITN effect parameters for permethrin bioassay mortality of 15.2% based on relationships shown in Fig 9. For kill rate, the logistic model for West African experimental hut studies were used and for blocking the loglogistic model based on all experimental hut trials as these seemed to reflect the relationships best (shaded in blue).

| **Model**<br>**(Nash et al 2021)** | **Nash EHT**<br>**mortality** | **Nash EHT**<br>**bloodfed** | **EMOD**<br>**kill rate** | **EMOD**<br>**blocking** |
|---|---|---|---|---|
| Logistic – west huts only | 0.101 | 0.476 | **0.101** | 0.524 |
| Logistic – all hut trials | 0.124 | 0.450 | 0.124 | 0.550 |
| Loglogistic – all hut trials | 0.303 | 0.274 | 0.303 | **0.726** |

Fixing the prior blocking rate in EMOD at 0.726, we simulated the Banfora trial control arm to recalibrate the kill rate parameter, sampling values above and below the prior of 0.101:

1. A simulation was run, with a) no ITN deployment after 2013 (date of previous mass distribution in Banfora district), or b) with ITN deployment in 2014 at 95% coverage (coverage obtained in the trial). The ITN kill rate was varied between 0 and 0.6 for nets deployed in 2013 and 2014. The simulation ran with 3 seeds and 32 kill rate values (equally spaced and including 0.101) and a single blocking value (0.726).
2. The 7 months following the 2014 mass campaign (June to December 2014) were used to calculate relative reduction in clinical cases compared to the counterfactual scenario, where no new nets were deployed in 2014.
3. The relationship between initial killing and reduction in clinical incidence followed a clear linear trend. A linear regression model was generated from the simulation data and used to calculate the initial kill rate value that corresponds to the reference effect size of 12.6% (Fig 10A): this value was 0.067.

Finally, we rescaled the statistical relationship for killing in Nash et al. to go through the two anchor points (Fig 10B). For blocking, we decided to directly use the Nash et al. statistical relationship without rescaling, as it corresponded well to the initial reference ITN blocking parameter and estimated blocking for untreated nets (Ngufor 2014, Bayili 2017, Bayili 2019, Toe 2018) (Fig 10C).

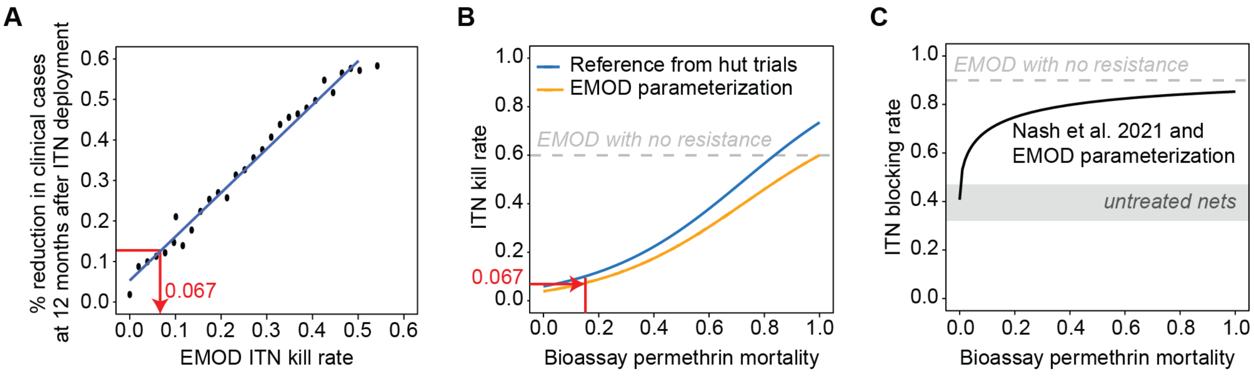

*Fig 10: A) Fitting kill rate in EMOD to achieve a reference effect size of 12.6% reduction in incidence. Linear regression model: kill rate = -0.05391 + 0.95254 * relative reduction. B) Re-scaled relationship between bioassay mortality and kill rate in EMOD (blue to orange line). The grey dashed line shows the reference ITN parameter from previous calibration in the context of no resistance (Eckhoff 2013). C) Relationship between bioassay mortality and blocking rate in EMOD. The grey dashed line shows the reference ITN parameter from previous calibration in the context of no resistance (Eckhoff 2013). The grey shaded rectangle shows estimated blocking for untreated nets based on Ngufor 2014, Bayili 2017, Bayili 2019, and Toe 2018.*

Finally, we compared modeled effect sizes against reported effect sizes from other studies. Another simulation was run using the maximum kill rate parameter values to compare the reduction in clinical cases to effect sizes from two Cochrane reviews (Chaplin 2021 and Pryce 2018), assessing standard ITNs in the early 2000s. The simulated ITN coverage was 95%. The modeled impact of 56.5-64.7%, during a 1 year follow up period, was slightly higher than reported in the Cochrane reviews (42-62%). Varying factors, such as case management, transmission intensity, follow up period, or pyrethroid resistance were not matched between the simulation and studies included in the review.

#### ITN efficacy for dual-insecticide nets

Efficacy of EMOD's ITN parameters were adjusted for PBO nets, as a first step to determine IG2 efficacy and for comparison, since stronger evidence exists on the impact of PBO nets compared to standard nets than for IG2. We adjusted the killing and blocking rate of PBO nets upward when compared to that of standard pyrethroid net, according to the statistical relationship in Sherrard-Smith et al. 2022, on which we applied the same scaling factors as determined in the calibration described above (Fig 11A).

The blocking effect of IG2 nets is mostly attributable to the pyrethroid component and the killing effect almost entirely from the chlorfenapyr, as demonstrated in EHTs in Tanzania and Cameroon (Tungu 2021, Tchouakui 2023). As with PBO, we adjusted the IG2 net's blocking rate upward when compared to that of standard pyrethroid net. The killing rate was optimistically assumed to be unaffected by pyrethroid resistance and chosen to be constant at 75% based on two EHT studies from West Africa (Bayili 2017, Bayili 2019, N'Guessan 2016, Ketoh 2018, Toe 2018) (Fig 11B).

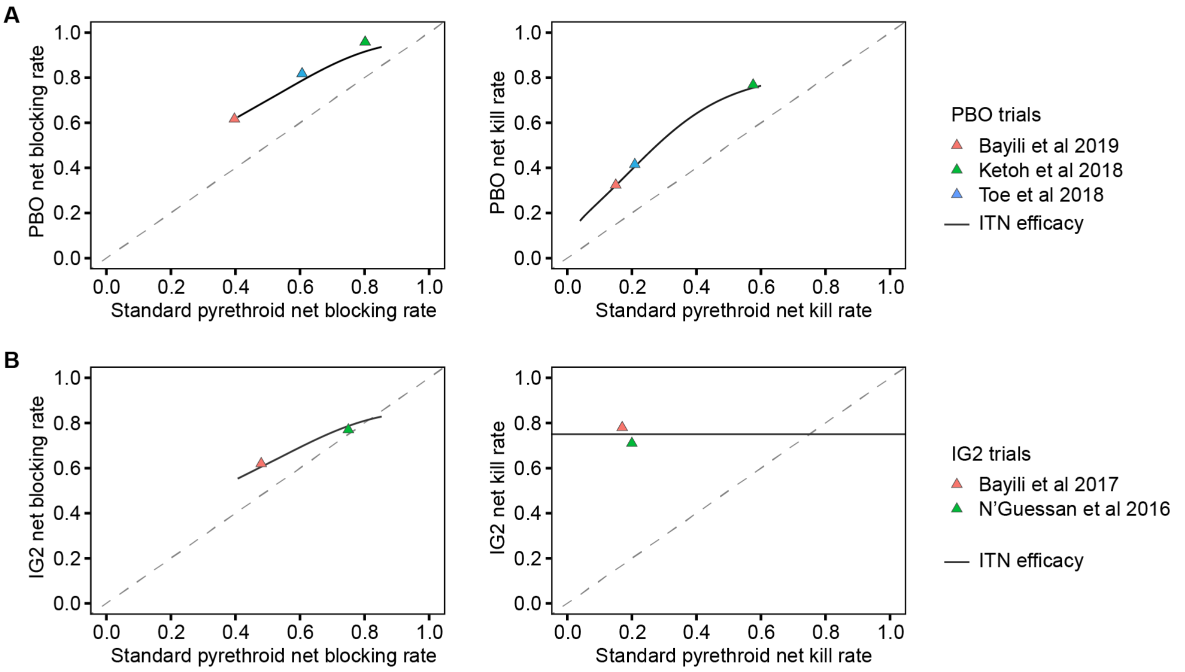

*Fig 11: Comparison of ITN effect parameter for standard LLINs to PBO and IG2 nets. A) A) standard ITN parameters on the x axis against PBO parameter on the y axis PBO ITN effect parameter scaled using statistical function from Sherrard-Smith et al 2022. The black solid line shows the ratio in efficacy obtained from the rescaled statistical relationship. The triangles reference points from West African PBO trials. B) standard ITN parameters on the x axis against IG2 parameter on the y axis. The black solid line shows the ratio and the triangles reference points from IG2 trials in Burkina Faso, Benin.*

#### ITN efficacy in Guinea

Mean permethrin mortality estimates for Guinea for 2005-2017 were estimated per district and year of distribution, accounting for resistance to pyrethroid resistance.

Permethrin resistance estimates were obtained from a geospatial model of insecticide resistance by Hancock and colleagues (Hancock 2020). The mean estimates were smoothed to remove any extreme fluctuations in resistance over time. Since estimates were only available up to 2017, and entomological data only available for a few study sites (WHO Malaria Threats Map,2022), three options for continuation of the trend 2017-2022 were presented to the PNLP (Fig 12). The first option was to assume the resistance level of 2017 to continue without further change, the second option were to assume a continued linear trend as seen between 2010 to 2017. Since the second option showed rapid increase in resistance in some areas, a third option was provided with the mean of the first two options, showing either no change or a slight decrease in permethrin mortality between 2017 and 2022. The PNLP selected the third option and for the years after 2022, 2022 resistance levels were assumed.

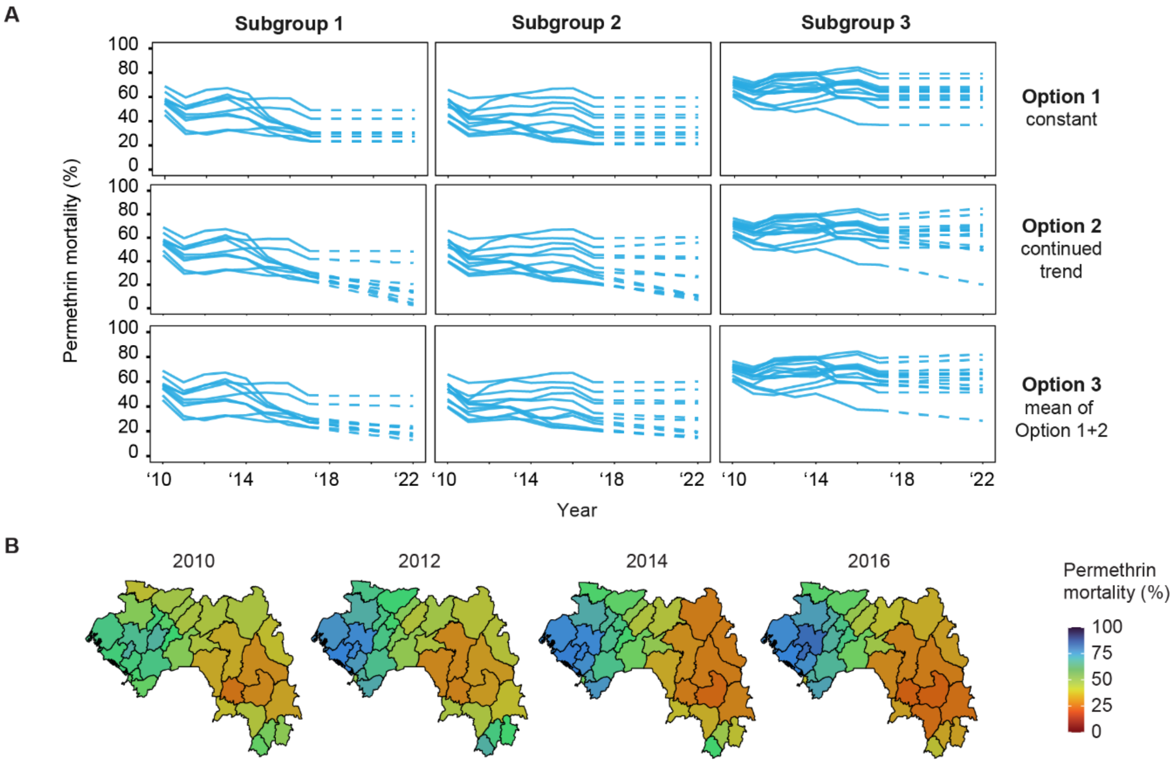

*Fig 12: **A)** Options of adjustments for permethrin resistance trends over time. Each line corresponds to one district. **B)** Mosquito mortality to permethrin per district corresponding to the selected Option 3 of A).*

At estimated resistance levels for 2022, pyrethroid-based nets block an average of 78% (range in districts 71-84%, depending on local resistance level) of mosquitoes, depending on deltamethrin resistance level, and the mortality rate was 21% (6-48%). IG2-based nets blocked an average of 80% (range in districts 77-83%) of mosquitoes, with the kill rate fixed at 75%. These blocking and killing parameters are the values required in the model to have the correct epidemiological impact according to the clinical trials of the nets and should not be interpreted as meaningful outside this context (Fig 13).

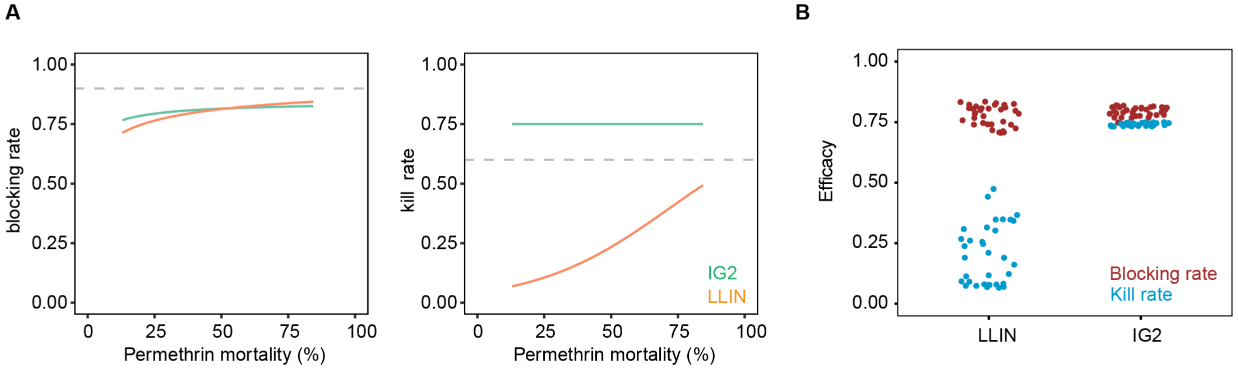

*Fig 13: Net efficacy and resistance. A) Conversion from permethrin mortality to ITN efficacy parameters for IG2 nets compared with pyrethroid LLINs. B) ITN efficacy parameter values per district (points) and ITN type extrapolated for the year 2022.*

#### SMC

##### SMC efficacy

In the model, SMC with SP and amodiaquine (SPAQ) initially provides nearly complete protection against clinical malaria, that wanes over a period of 6 weeks, averting half of all clinical cases at 4-5 weeks and zero protection at 6 weeks. The initial effect size, defined as the reduction in force of infection after a child received a drug, as well as the waning over time, consisting of a constant effect duration parameter and parameter defining exponential decay were fit to a SMC trial conducted in Burkina Faso (Lena district) between August 2009 and January 2010 (Zongo 2015).

In that study, malaria transmission is highly seasonal and the rainy season to occur between July and October. Four rounds of SMC were administered to children aged 3 to 59 months in monthly intervals starting on August 1$^{st}$. A EMOD malaria model with constant transmission intensity (no vectors) was used to simulate a single round of SMC in absence of other interventions except for treatment of clinical and severe malaria cases. The simulated seasonality was informed by the SMC trial while transmission levels were varied to obtain and compare transmission specific efficacy parameters.

We used a sequential Monte Carlo procedure, which is an approximate Bayesian computation, to search for the best parameters that fit to the study's efficacy. First, 500 sets of parameters (also known as particles) were sampled from prior distributions for all three parameters: uniform distributions with a range of 0.8-1 for the initial effect size, 7-20 days for the constant effect duration, and 2-40 days for the exponential decay parameter. The particles were used to run simulations and were ranked according to the mean square errors between simulated and observed weekly trial efficacy. The best 50 particles were selected for the next round, with 450 new particles created by perturbation based on the 50 particles. The perturbation was done with a multivariate normal sampling with mean of each of the 50 particles, and covariance of 2 times the 50-particle sample covariance. The process was repeated for at least four rounds to arrive at the best 10 particles. The 10 best particles were further tested across varying transmission levels and the best fitting one at moderate-high transmission (annual EIR=13) was selected.

In the parameterization used, only the prophylactic effect of SMC was simulated, keeping the efficacy constant before exponential decline. Final model parameters were an initial efficacy of 98% against infection for 13 days, followed by an exponential decrease in efficacy with a half-life of 28 days. After fitting, the cumulative efficacy was compared to results from a meta-analysis including 7 studies after 28 days and after 42 days (Cairns 2021 and Wilson 2011). The calibration and validation simulations further show variation in efficacy depending on transmission intensity and highlight uncertainty in the potential rebound in incidence after the effect of SMC waned completely (Fig 14).

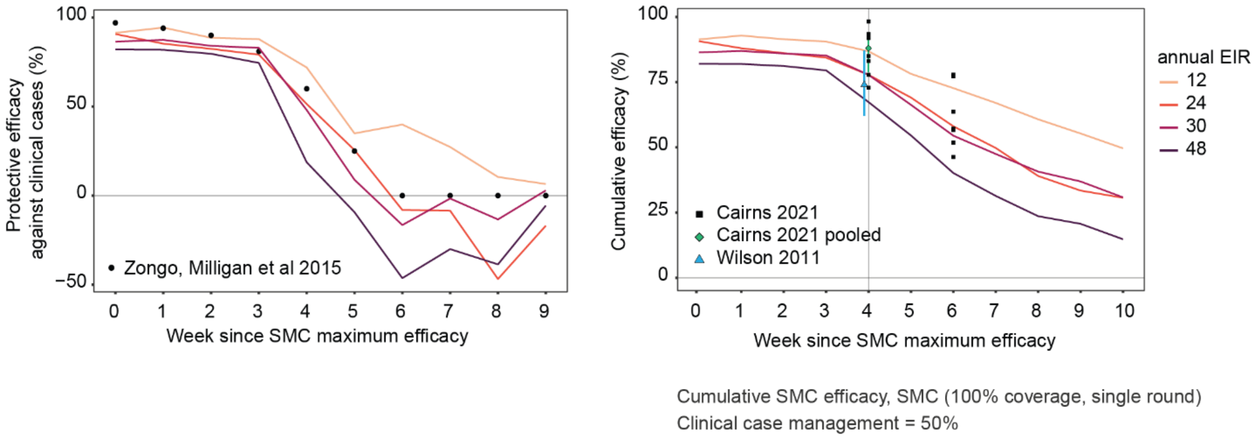

*Fig 14: Assumed SMC efficacy curves against clinical malaria in the simulation, after calibration to Zongo 2015 and validation of cumulative protection against pooled estimates (Cairns 2021, Wilson 2011).*

#### SMC coverage

The SMC coverages were based on estimates of the mean number of SMC cycles per child per year in each SMC district from 2015 to 2020 obtained from (Loua and Milligan 2019, 2020, 2021). These studies also provided estimates of percentage of children who received 0 to 4 cycles of SMC in each year in each group of districts. In this study, the percentage of children receiving 4 cycles tended to be the highest, following by 0 cycle in most districts, indicating likely semi-correlated and non-random SMC coverage within the population, with some having excellent access to SMC and others no access to SMC at all.

We used numerical optimization to determine (a) the fraction of the population who received no SMC dose, and (b) the per-cycle SMC coverage of the population with access to SMC, assuming that coverage in each cycle is independent (Fig 15 and 16). For each district in each year, we found the combination of the two coverage parameters that minimized the square errors for mean number of SMC cycles received, and the fraction of children receiving 0 to 4 cycles in Loua and Milligan 2021.

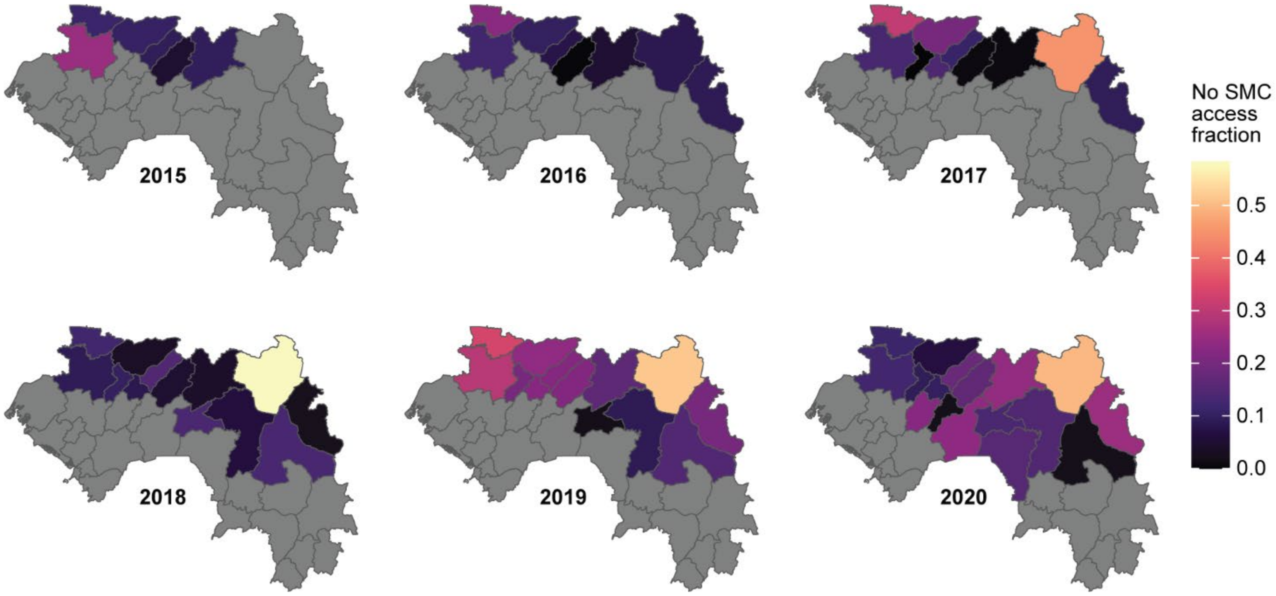

*Fig 15: Fraction of people who had no access to SMC, estimated using numerical optimization and data from Loua and Milligan (2021).*

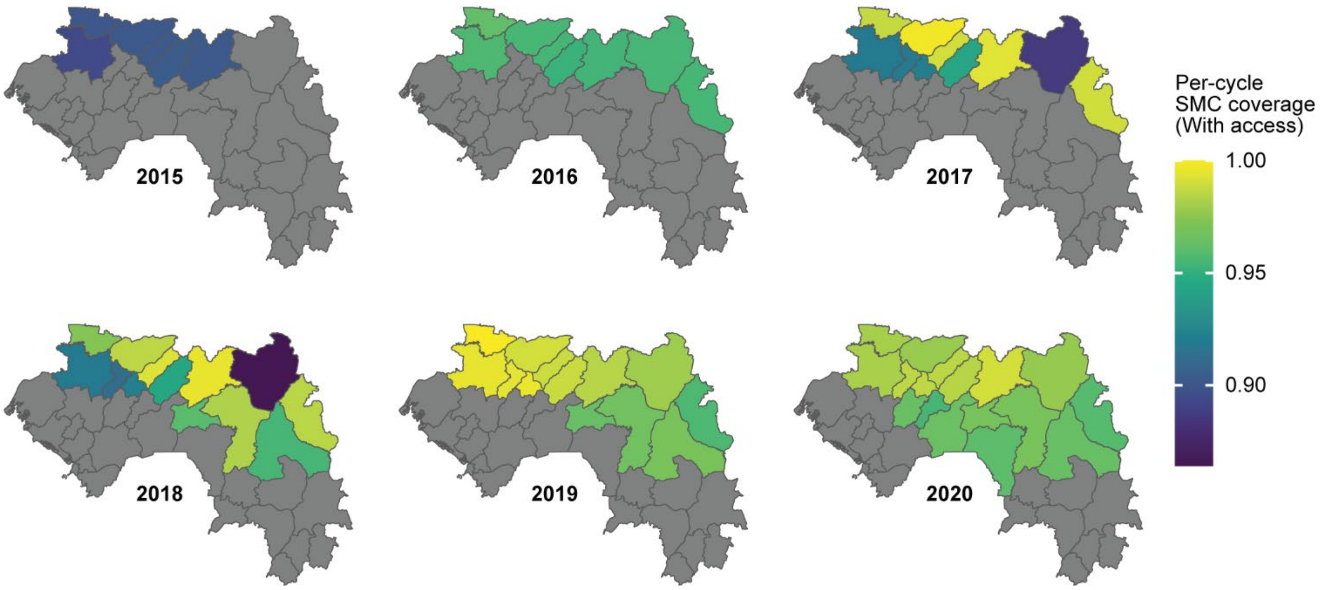

*Fig 16: Coverage for each SMC cycle among people with access to the drugs, estimated using numerical optimization and Loua and Milligan (2021).*

SMC doses may occasionally go to children above 5 years old who are ineligible for SMC. In the model, SMC is assumed to "leak" to children from 5 to 10 years old. Loua and Milligan (2021) provides nationwide estimates of mean number of cycles received per year, and the SMC coverage per cycle per year for children aged 6 to 7 years at the survey. Unlike children under 5, we assumed that the entire population of 5 to 10 years old has access to SMC, which is consistent with the study estimates. For each year, the ratio between mean number of SMC cycles received between over and under 5 years old is calculated. Then for each district, the per-cycle coverage for children from 5 to 10 years old is assumed to be the mean coverage of children under 5 multiplied by the ratio of the corresponding year (Fig 17).

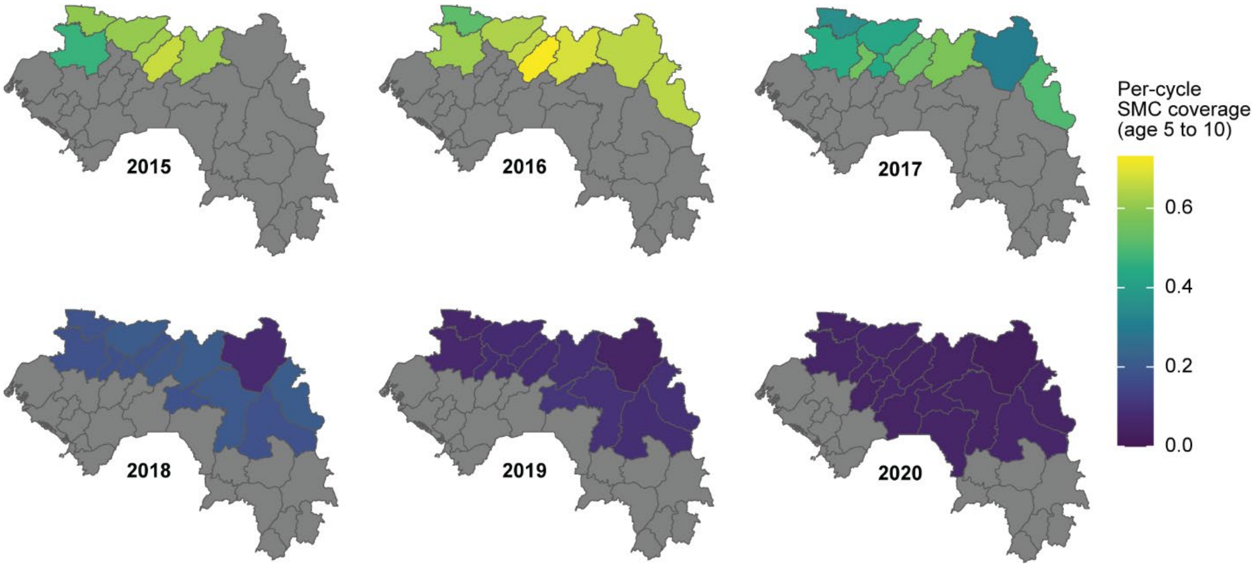

*Fig 17: SMC coverage for age 5 to 10 for each district and year assumed in the model.*

#### PMC

##### PMC Efficacy

PMC initially provides 80% protection against new infections with the protection waning exponentially reaching 40% protection after 4-5 weeks and zero protection eight weeks after administration of the drug. This study used the PMC parameterization from Runge 2023. In that study, using EMOD, effect parameters were fitted to the estimated efficacy curve of a single dose of SP on clinical cases based on a randomized controlled trial conducted in Ghana in the early 2000s (Chandramohan 2005, Cairns 2008), assuming constant efficacy fifteen days before reaching peak efficacy to compensate lack of parasite clearance.

##### PMC Coverage

The vaccination coverages for diphtheria, tetanus, and pertussis (DTP) 2, DTP3 and measles, administered at the ages of 10, 14 weeks and 9 months through the Expanded Program of Immunization (EPI) were retrieved from the Guinea DHS 2018 according to the DHS guide for calculating vaccination coverage (DHS Program, 2023). After internal inspection of the coverage values and comparison to coverage from routine data in Guinea, regional values were selected for use as input for the model. Based on the experience of PMC implementation in Sierra Leone (Lahuerta 2021), PMC coverage can be lower than vaccine coverage, hence was downscaled for the model per dose, using 0.838, 0.9556, and 0.697 as scaling factors, which were the per-dose ratios of PMC coverage to vaccine coverage observed in Lahuerta 2021.

In the model, PMC was administered independently of individuals' malaria infection or treatment status, and each dose was distributed regardless of whether the child had received a previous dose. Children received each dose of PMC exactly as planned, without delay.

The resulting coverages ranged between 40% and 56% for the first dose, between 30% and 52% for the 2$^{nd}$ dose, and 32 to 50% for the 3$^{rd}$ dose, and between 34% to 53% for all three doses (Fig 18).

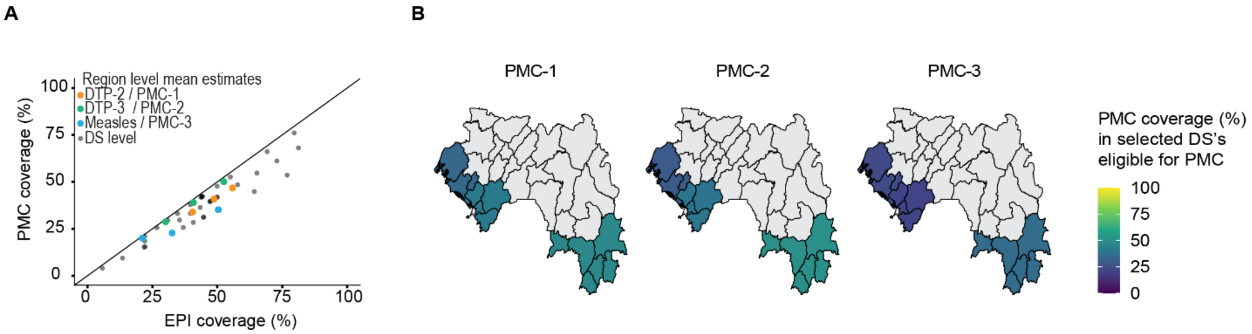

*Fig 18: Assumed PMC coverage in eligible areas in Guinea (n=13) with a) scatter plot showing the coverage and EPI-to-PMC conversion, and b) coverage map showing regional coverage values used for each PMC eligible DS.*

#### Malaria vaccine (RTS,S)

##### RTS,S Efficacy

The malaria vaccine RTS,S was modeled with a proportionate reduction in the force of infection that wanes exponentially. The initial efficacy was 80% after completion of the primary sequence at 9 months of age that waned over 13.5 months. These parameters were previously obtained for EMOD based on calibration to phase-3 trial data (RTS,S Clinical Trials Partnership 2015) published by Penny and colleagues (Penny, 2016). The booster dose at 24 months of age had the same initial efficacy and waning profile as the priming sequence.

##### RTS,S Coverage

The primary sequence of RTS,S was simulated assuming the same schedule as used in the efficacy calibration (Penny, 2016), with the 3rd dose at 9 months of age. Hence, the vaccination coverage for the measles vaccine, administered at 9 months of age, was used to inform the coverage of the primary sequence of RTS,S in the model. The measles vaccination coverage data was obtained from the 2018 DHS in Guinea (since the 2021 national survey was an MIS and did not include questions on vaccination) at the district and regional levels following the same approach as for PMC. The regional level estimates were used as they better agreed with coverages obtained from routine data. In the absence of sufficient data on vaccination uptake in the second year of life, eighty percent of the children that received the primary sequence were assumed to receive a booster at exactly 24 months of age. This aligns with findings from the RTS,S phase 3 trial data in Ghana, in which the booster dose had substantially lower coverage compared to the primary sequence (Yeboah 2022). Across Guinea's regions, the assumed RTS,S primary sequence coverage ranged from 18% to 51%, with a mean of 36% (Fig 19).

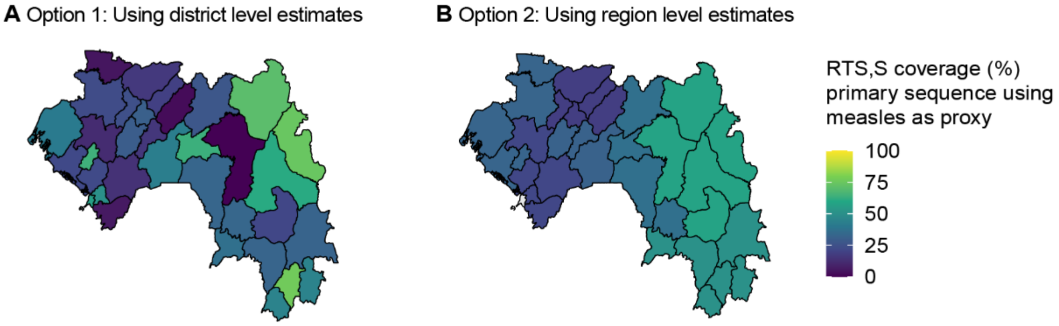

*Fig 19: Options for assumed RTS,S coverage of primary sequence completed at 9 month of age per A) district and B) region level. Regional estimates were used based on internal discussion and comparison to routine data.*

#### Setting modeled seasonality

Archetypes were determined by clustering the districts based on their monthly rainfall, monthly temperature suitability index for anophelines, and relative abundance of three vector species. Monthly rainfall was obtained from CHIRPS, and temperature suitability and relative vector abundance from the Malaria Atlas Project. Similar to the approach in Ozodiegwu 2023, we used clustering algorithm CLARA to identify candidate sets of archetypes ranging from 3-12 archetypes; these were presented to the PNLP. After discussion, the PNLP selected the clustering with 9 archetypes as most representative of transmission seasonality in Guinea (Fig 20).

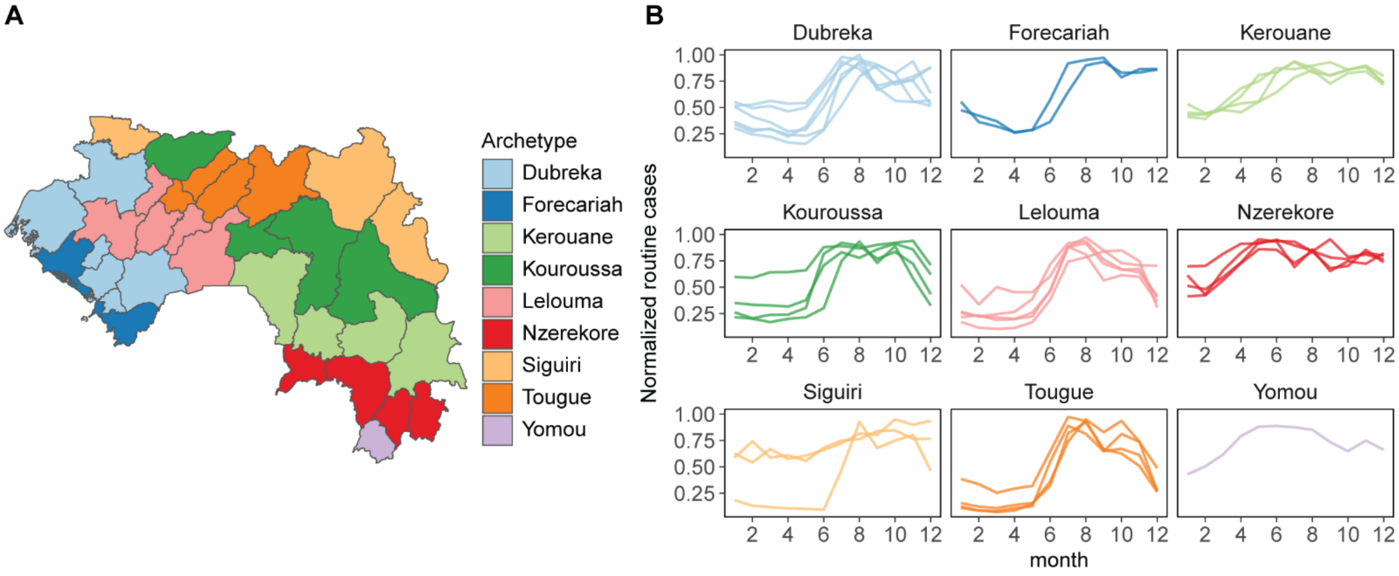

*Fig 20: (A) The 34 districts in Guinea are grouped into nine seasonality archetypes for seasonality calibration purpose. The archetypes are named after one of the districts within the archetype. (B) The mean monthly routine cases, normalized by the maximum number of cases in the corresponding district and year. The routine cases shown here are the number of confirmed and suspected cases. Each line represents a district within the archetype.*

For each archetype, we summed the mean monthly routine cases for people over 5 years of age from 2018 to 2022 in all districts. We chose routine case data for those over 5 years old, as incidence in children under 5 would presumably be influenced by the presence of SMC. The mean monthly cases were the mean of the number of cases observed in all January months, the mean in all February months, etc. for the 12 months of the year. For each archetype, we scaled the mean monthly cases to the maximum number of monthly cases. This resulted in, for each archetype, a 12-month case series where the shape of the series was informative of average seasonality of malaria cases in the archetype, but due to the rescaling, no information about the intensity of cases was preserved.

We adjusted the monthly availability of modeled mosquito habitat, by trial and error, such that the scaled clinical incidence in individuals over 5 years of age in the model tracked that of the archetype monthly average (Fig 21). We also ensured that the resulting simulation had an annual incidence that was in the same order of magnitude as that of the adjusted incidence (adjustment 3 using the non-treatment-seeking rate from DHS, see main text Methods for explanation of adjustment 3 and

Supplementary File 1 for maps of incidence under adjustment 3 when using the non-treatment-seeking rate from DHS) of the districts within the archetype. Modeled seasonality can vary substantially if the overall transmission level deviates too much.

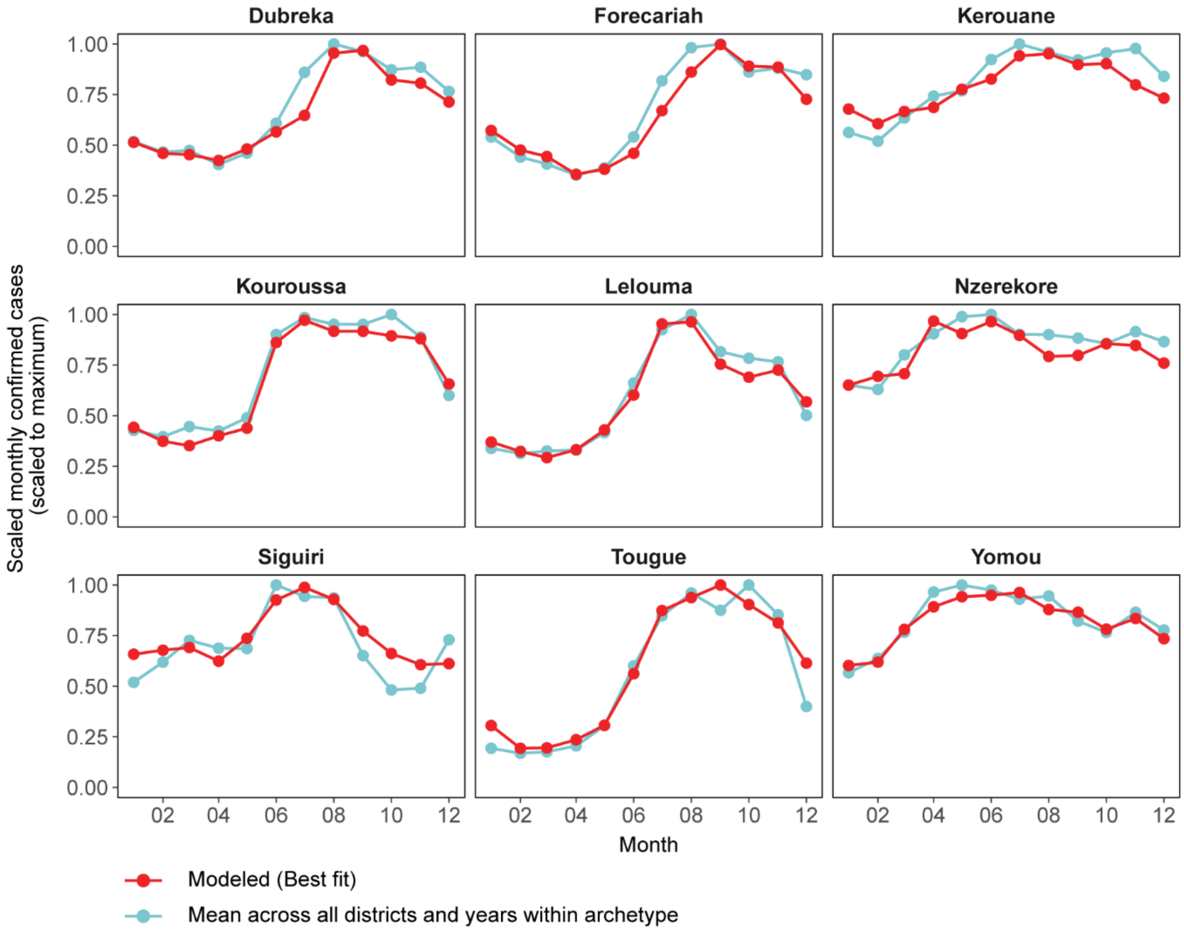

*Fig 21: Seasonality calibration outcomes for the nine archetypes. The monthly availability of modeled mosquito habitat was adjusted such that the modeled scaled monthly cases are similar to that of the average of routine cases for those above 5 years old.*

#### Setting modeled transmission intensity

Each district's modeled transmission intensity was fit to prevalence data from DHS/MIS and incidence from routine surveillance (adjustment 3, using the non-treatment-seeking rate from DHS).

Transmission intensity in the model is determined by the amount of habitat available for mosquito larvae. This amount was sampled at 50 candidate levels, evenly in log space, such that mean mosquito biting rate per human was bounded between 0.1 and 20 per day, and was a scale factor on top of the monthly habitat availability derived from the seasonality fitting. Interventions were included following the schedule, coverage, and effect sizes described above. To account for uncertainty in intervention coverage, prior distributions for case management coverage (at 2012, 2018, and 2021 timepoints), and ITN usage (2013, 2016, and 2019 timepoints) were derived and sampled from simultaneously with the habitat availability. The priors for the intervention coverages were normal on the logistic scale, approximated according to the 95% interval determined in the geostatistical model (see Case Management section). All priors were independent. Latin Hypercube Sampling was used to construct 1000 sets of parameters, each set including 1 habitat availability, 3 case management coverages, and 3 ITN usages. Three stochastic realizations were simulated for each parameter set for the period 1960 to 2020, resulting in 3000 simulations per district.

For each district, the monthly modeled prevalence was compared with the aggregated monthly prevalence in children under 5 in DHS/MIS from 2012 to 2021 across all clusters falling within the district boundaries. The modeled annual incidence in children under 5 was compared with the annual incidence in children under 5 (adjustment 3) from 2018 to 2021. The goodness of fit for each parameter set was evaluated using a binomial negative log-likelihood (per person) for prevalence and the mean square errors for incidence. The final score was a weighted sum of the likelihood and error measures with a weight of 30 to 1. The weight was chosen to offset the scale difference between the measures and to reflect greater confidence in the prevalence data compared to the adjusted incidence data. For each district, the 20 parameter sets with the lowest score were selected and used for impact projections.

The calibration process for the district Lelouma is shown below as a representative example. The transmission intensity in the model is controlled by a habitat multiplier parameter. The level of larval habitat availability is scaled up or down by the parameter, and stayed the same as that used in the seasonal calibration step if the multiplier is 1. Fig 22 shows the marginal distributions of the habitat multiplier and the intervention coverages before and after the parameter selection process. In Fig 23, the monthly prevalence and annual incidence trajectory for each of the 20 selected parameter sets are presented. Because of the final goodness of fit favors the prevalence data, these trajectories are close to the DHS/MIS prevalence data, but somewhat different from the adjusted incidence data (Fig 24.

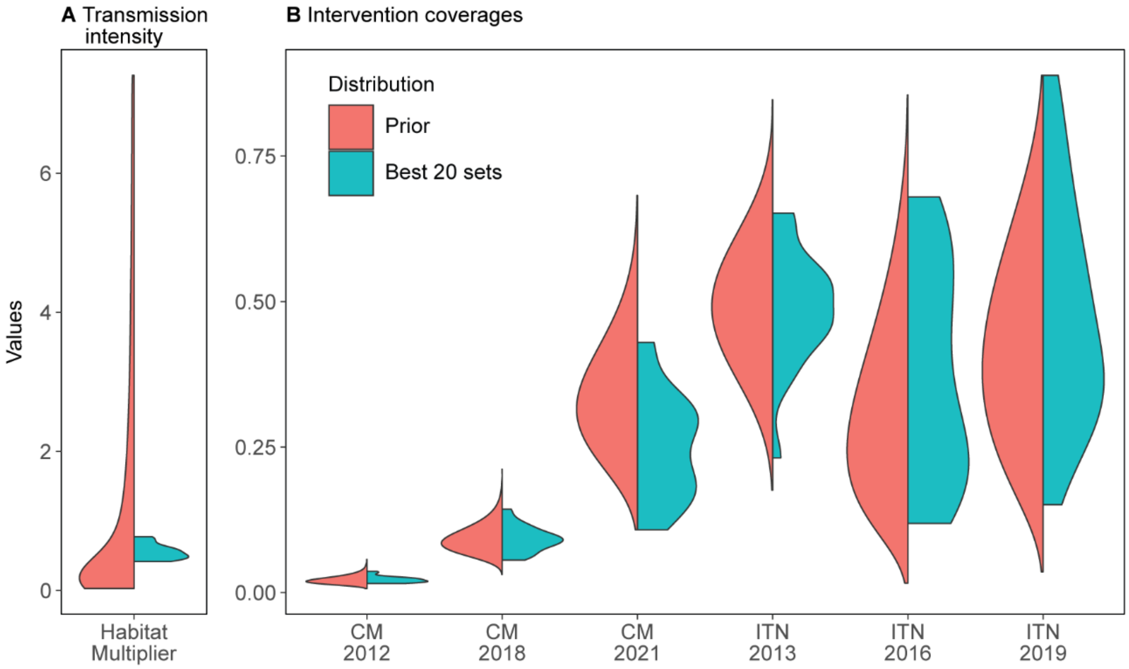

*Fig 22: The marginal distribution of the transmission intensity and intervention coverages parameters before and after the parameter selection in the Lelouma district. CM = Case management, ITN = Insecticide-treated net. Prior distribution of the habitat multiplier is uniform in log space, and the prior distributions for intervention coverages are normal in logistic space. All priors are assumed to be independent.*

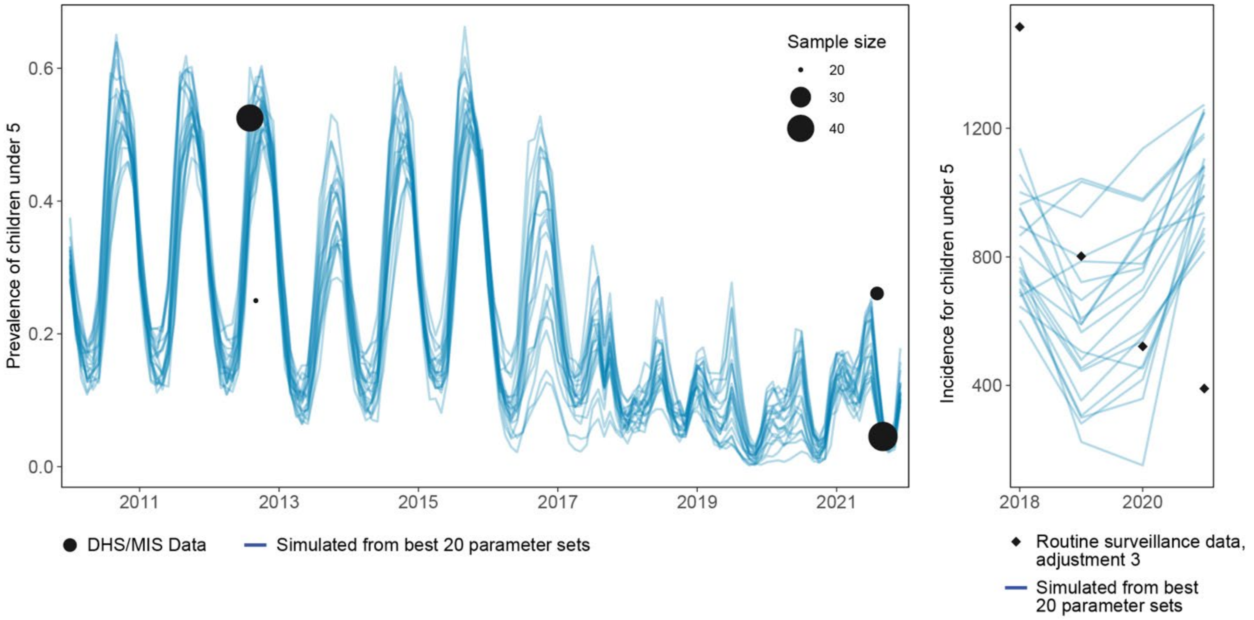

*Fig 23: Simulated prevalence and incidence of children under 5 in Lelouma district vs data from DHS/MIS (for prevalence) and routine case data (for adjusted incidence). Each line represents the average of one parameter set and 20 best parameter sets are presented here.*

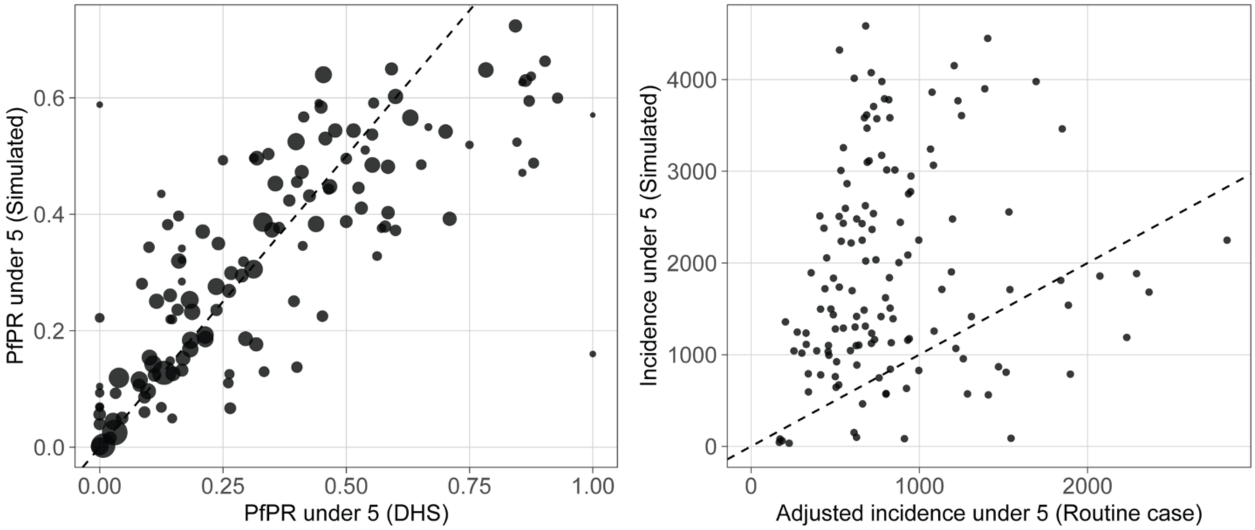

*Fig 24: Comparison of simulated and measured epidemiological indicators. Left: Prevalence in children under 5 at the district level, all surveys combined. Right: Incidence in children under 5 years of age for 2018-2021.*

#### Aggregating district-level output

For each district, 15 realizations were run for each of the 20 selected parameter sets. Thus, we obtained 300 runs for each district. To obtain projections on national level or on specific zones, we aggregate the district level output.

We created 1000 sets of aggregates: for each set, we randomly selected one of the 300 runs for each district. We then calculated the mean clinical incidence and deaths across the district, weighted by the district populations. Based on the 1000 sets of aggregate, we obtained the mean and 95% PI. To ensure comparability and consistency across all scenarios, each set of aggregate across all the different scenarios was created by the same parameter set, random number seed and district combination.

### References

Bayili K, N'do S, Namountougou M, Sanou R, Ouattara A, Dabiré RK, Ouédraogo AG, Malone D, Diabaté A. Evaluation of efficacy of Interceptor® G2, a long-lasting insecticide net coated with a mixture of chlorfenapyr and alpha-cypermethrin, against pyrethroid resistant *Anopheles gambiae sl* in Burkina Faso. Malaria Journal. 2017 Dec;16(1):1-9.

Bayili K, N'Do S, Yadav RS, Namountougou M, Ouattara A, Dabiré RK, Ouédraogo GA, Diabaté A. Experimental hut evaluation of DawaPlus 3.0 LN and DawaPlus 4.0 LN treated with deltamethrin and PBO against free-flying populations of *Anopheles gambiae s. l.* in Vallée du Kou, Burkina Faso. PloS one. 2019 Dec 23;14(12):e0226191.

Bershteyn A, Gerardin J, Bridenbecker D, Lorton C, et al. on behalf of the Institute for Disease Modeling. Implementation and applications of the EMOD individual-based disease modeling platform: software design and development processes to enable multi-scale modeling. Pathog Dis. 2018; 76(5).

Bertozzi-Villa A, Bever CA, Koenker H, Weiss DJ, Vargas-Ruiz C, Nandi AK, Gibson HS, Harris J, Battle KE, Rumisha SF, Keddie S. Maps and metrics of insecticide-treated net access, use, and nets-per-capita in Africa from 2000-2020. Nature communications. 2021 Jun 11;12(1):3589.

Cairns M, Carneiro I, Milligan P, Owusu-Agyei S, Awine T, Gosling R, Greenwood B, Chandramohan D. Duration of protection against malaria and anaemia provided by intermittent preventive treatment in infants in Navrongo, Ghana. PLoS One. 2008 May 21;3(5):e2227.

Cairns M, Ceesay SJ, Sagara I, Zongo I, Kessely H, Gamougam K, Diallo A, Ogboi JS, Moroso D, Van Hulle S, Eloike T. Effectiveness of seasonal malaria chemoprevention (SMC) treatments when SMC is implemented at scale: Case–control studies in 5 countries. PloS medicine. 2021 Sep 8;18(9):e1003727.

Chandramohan D, Owusu-Agyei S, Carneiro I, Awine T, Amponsa-Achiano K, Mensah N, Jaffar S, Baiden R, Hodgson A, Binka F, Greenwood B. Cluster randomized trial of intermittent preventive treatment for malaria in infants in area of high, seasonal transmission in Ghana. BMJ. 2005 Sep 29;331(7519):727-33.

Chandramohan D, Zongo I, Sagara I, Cairns M, Yerbanga RS, Diarra M, Nikiema F, Tapily A, Sompougdou F, Issiaka D, Zoungrana C. Seasonal malaria vaccination with or without seasonal malaria chemoprevention. New England Journal of Medicine. 2021 Sep 9;385(11):1005-17.

Chaplin M, Choi L, Ranson H. Piperonyl butoxide (PBO) combined with pyrethroids in insecticide-treated nets to prevent malaria in Africa. Cochrane Database of Systematic Reviews. 2021(5).

Cherif MS, Dahal P, Beavogui AH, Delamou A, Lama EK, Camara A, Diallo MP. Malaria epidemiology and anti-malarial drug efficacy in Guinea: a review of clinical and molecular studies. Malaria Journal. 2021 Jun 16;20(1):272.

Dalrymple U, Cameron E, Arambepola R, Battle KE, Chestnutt EG, Keddie SH, Twohig KA, Pfeffer DA, Gibson HS, Weiss DJ, Bhatt S. The contribution of non-malarial febrile illness co-infections to *Plasmodium falciparum* case counts in health facilities in sub-Saharan Africa. Malaria Journal. 2019 Dec;18(1):1-2.

The DHS Program. Percentage of children age 12-23 months and children age 24-35 months who received specific vaccines at any time before the survey according to vaccination card, according to mother's report, according to either vaccination card or mother's report, and percentage who received specific vaccines by appropriate age. [Internet]. Accessed 25 November 2023. https://dhsprogram.com/data/Guide-to-DHS-Statistics/Vaccination.htm

Eckhoff P. Mathematical models of within-host and transmission dynamics to determine effects of malaria interventions in a variety of transmission settings. The American journal of tropical medicine and hygiene. 2013 May 5;88(5):817.

Gerardin J, Eckhoff P, Wenger EA. Mass campaigns with antimalarial drugs: a modelling comparison of artemether-lumefantrine and DHA-piperaquine with and without primaquine as tools for malaria control and elimination. BMC infectious diseases. 2015 Dec;15(1):1-4.

Gerardin J, Ouédraogo AL, McCarthy KA, Eckhoff PA, Wenger EA. Characterization of the infectious reservoir of malaria with an agent-based model calibrated to age-stratified parasite densities and infectiousness. Malaria Journal. 2015 Dec;14(1):1-3.

Guelbeogo, W. M., Sagnon, N., Liu, F., Besansky, N. J. & Costantini, C. Behavioural divergence of sympatric *Anopheles funestus* populations in Burkina Faso. Malar. J. 2014; 13, 65.

Hancock PA, Hendriks CJ, Tangena JA, Gibson H, Hemingway J, Coleman M, Gething PW, Cameron E, Bhatt S, Moyes CL. Mapping trends in insecticide resistance phenotypes in African malaria vectors. PLoS Biology. 2020 Jun 25;18(6):e3000633.

Hien AS, Soma DD, Sawadogo SP, Poda SB, Namountougou M, Ouédraogo GA, et al. Effect of Bendiocarb (Ficam® 80% WP) on Entomological Indices of Malaria Transmission by Indoor Residual Spraying in Burkina Faso, West Africa. AE. 2020;08(04):158–78.

Institut national de la Statistique (INS) [Guinée] et ICF. 2018. Enquête demographique et de santé et à indicateurs multiples (EDS-MICS 2012). 2012. Conakry, Guinée, et Rockville, Maryland, USA : INS et ICF.

Institut national de la Statistique (INS) [Guinée] et ICF. 2018. Enquête Démographique et de Santé en Guinée 2018. Conakry, Guinée, et Rockville, Maryland, USA : INS et ICF.

Institut national de la Statistique (INS) [Guinée], et ICF. 2021. Enquête sur les indicateurs du paludisme et de l'anémie en Guinée 2021. Rockville, Maryland, USA : INS et ICF.

Institute for Disease Modeling (IDM). Epidemiological Modeling Software (EMOD) [Internet]. 2023. Available from: https://www.idmod.org/tool/emod/

Ketoh GK, Ahadji-Dabla KM, Chabi J, Amoudji AD, Apetogbo GY, Awokou F, Glitho IA. Efficacy of two PBO long lasting insecticidal nets against natural populations of *Anopheles gambiae s. l.* in experimental huts, Kolokopé, Togo. PLoS one. 2018 Jul 11;13(7):e0192492.

Lahuerta M, Sutton R, Mansaray A, Eleeza O, Gleason B, Akinjeji A, Jalloh MF, Toure M, Kassa G, Meshnick SR, Deutsch-Feldman M. Evaluation of health system readiness and coverage of intermittent preventive treatment of malaria in infants (IPTi) in Kambia district to inform national scale-up in Sierra Leone. Malaria Journal. 2021 Dec;20(1):1-3.

Lengeler C. Insecticide-treated bed nets and curtains for preventing malaria. Cochrane database of systematic reviews. 2004(2).

Lloyd, C.T., Chamberlain, H., Kerr, D., Yetman, G., Pistolesi, L., Stevens, F.R., Gaughan, A.E., Nieves, J.J., Hornby, G., MacManus, K., Sinha, P., Bondarenko, M., Sorichetta, A., Tatem, A.J. Global spatio-temporally harmonised datasets for producing high-resolution gridded population distribution datasets. Big Earth Data. 2019 Apr 3;3(2):108–39.

Loua, Kovana Marcel; Milligan, Paul; (2019) Seasonal Malaria Chemoprevention Coverage Survey Guinea, 2018. Project Report. LSHTM. DOI: https://doi.org/10.17037/PUBS.04654302

Loua, Kovana Marcel; Milligan, Paul; (2020) Seasonal Malaria Chemoprevention Coverage in Guinea in 2019. Technical Report. London School of Hygiene & Tropical Medicine, London and Universite Gamal Abdel Nasser, Conakry. DOI: https://doi.org/10.17037/PUBS.04663124

Loua, Kovana Marcel; Milligan, Paul; (2021) Seasonal Malaria Chemoprevention in Guinea in 2020: coverage survey results. Technical Report. London School of Hygiene & Tropical Medicine, London and Universite Gamal Abdel Nasser, Conakry. DOI: https://doi.org/10.17037/PUBS.04663123

Martins, T. G., Simpson, D., Lindgren, F. & Rue, H. Bayesian computing with INLA: New features. Computational Statistics & Data Analysis. 2013, 67, 68–83.

McCarthy KA, Wenger EA, Huynh GH, Eckhoff PA. Calibration of an intrahost malaria model and parameter ensemble evaluation of a pre-erythrocytic vaccine. Malaria Journal. 2015 Dec;14(1):1-0.

Nash RK, Lambert B, N'Guessan R, Ngufor C, Rowland M, Oxborough R, Moore S, Tungu P, Sherrard-Smith E, Churcher TS. Systematic review of the entomological impact of insecticide-treated nets evaluated using experimental hut trials in Africa. Current research in parasitology & vector-borne diseases. 2021 Jan 1;1:100047.

N'Guessan R, Odjo A, Ngufor C, Malone D, Rowland M. A chlorfenapyr mixture net Interceptor® G2 shows high efficacy and wash durability against resistant mosquitoes in West Africa. PLoS One. 2016 Nov 16;11(11):e0165925.

Ngufor C, Tchicaya E, Koudou B, N'Fale S, Dabire R, Johnson P, Ranson H, Rowland M. Combining organophosphate treated wall linings and long-lasting insecticidal nets for improved control of pyrethroid resistant *Anopheles gambiae*. PLoS one. 2014 Jan 7;9(1):e83897.

Nguyen M, Dzianach PA, Castle PE, Rumisha SF, Rozier JA, Harris JR, Gibson HS, Twohig KA, Vargas-Ruiz CA, Bisanzio D, Cameron E. Trends in treatment-seeking for fever in children under five years old in 151 countries from 1990 to 2020. PLoS Global Public Health. 2023 Aug 23;3(8):e0002134.

Ozodiegwu ID, Ambrose M, Galatas B, Runge M, Nandi A, Okuneye K, Dhanoa NP, Maikore I, Uhomoibhi P, Bever C, Noor A. Application of mathematical modelling to inform national malaria intervention planning in Nigeria. Malaria journal. 2023 Dec;22(1):1-9.

PATH. New Nets Project Interim Report: Output 3: Evidence of effectiveness and cost-effectiveness of dual-AI ITNs created and disseminated. July 2022.

Penny MA, Verity R, Bever CA, Sauboin C, Galactionova K, Flasche S, White MT, Wenger EA, Van de Velde N, Pemberton-Ross P, Griffin JT. Public health impact and cost-effectiveness of the RTS, S/AS01 malaria vaccine: a systematic comparison of predictions from four mathematical models. The Lancet. 2016 Jan 23;387(10016):367-75.

Pryce J, Richardson M, Lengeler C. Insecticide-treated nets for preventing malaria. Cochrane Database of Systematic Reviews. 2018(11).

RTS,S Clinical Trials Partnership. Efficacy and safety of RTS, S/AS01 malaria vaccine with or without a booster dose in infants and children in Africa: final results of a phase 3, individually randomised, controlled trial. Lancet. 2015 Jul 4;386(9988):31-45.

H. Rue, S. Martino, N. Chopin. Approximate Bayesian inference for latent Gaussian models using integrated nested Laplace approximations (with discussion). Journal of the Royal Statistical Society. 2009 Series B, 71(2):319{392.

Runge M, Stahlfeld A, Ambrose M, Toh KB, Rahman S, Omoniwa OF, Bever CA, Oresanya O, Uhomoibhi P, Galatas B, Tibenderana JK. Perennial malaria chemoprevention with and without malaria vaccination to reduce malaria burden in young children: a modelling analysis. Malaria Journal. 2023 Dec;22(1):1-3.

Selvaraj P, Wenger EA, Gerardin J. Seasonality and heterogeneity of malaria transmission determine success of interventions in high-endemic settings: a modeling study. BMC Infectious Diseases. 2018 Dec;18(1):1-4.

Sherrard-Smith E, Winskill P, Hamlet A, Ngufor C, N'Guessan R, Guelbeogo MW, Sanou A, Nash RK, Hill A, Russell EL, Woodbridge M. Optimising the deployment of vector control tools against malaria: a data-informed modelling study. The Lancet Planetary Health. 2022 Feb 1;6(2):e100-9.
Sinka ME, Golding N, Massey NC, Wiebe A, Huang Z, Hay SI, Moyes CL. Modelling the relative abundance of the primary African vectors of malaria before and after the implementation of indoor, insecticide-based vector control. Malaria Journal. 2016 Dec;15(1):1-0.

Tchouakui M, Thiomela RF, Nchoutpouen E, Menze BD, Ndo C, Achu D, et al. High efficacy of chlorfenapyr-based net Interceptor® G2 against pyrethroid-resistant malaria vectors from Cameroon. Infectious Diseases of Poverty. 2023 Aug 29;12(1):81.

The World Bank. World Bank Open Data. [Internet]. Birth rate, crude (per 1,000 people) - Guinea. Available from: https://data.worldbank.org/indicator/SP.DYN.CBRT.IN?locations=GN.
Tiono AB, Ouédraogo A, Ouattara D, Bougouma EC, Coulibaly S, Diarra A, Faragher B, Guelbeogo MW, Grisales N, Ouédraogo IN, Ouédraogo ZA. Efficacy of Olyset Duo, a bednet containing pyriproxyfen and permethrin, versus a permethrin-only net against clinical malaria in an area with highly pyrethroid-resistant vectors in rural Burkina Faso: a cluster-randomized controlled trial. The Lancet. 2018 Aug 18;392(10147):569-80.

Toe KH, Müller P, Badolo A, Traore A, Sagnon N, Dabiré RK, Ranson H. Do bednets including piperonyl butoxide offer additional protection against populations of *Anopheles gambiae sl. t*hat are highly resistant to pyrethroids? An experimental hut evaluation in Burkina Fasov. Medical and veterinary entomology. 2018 Dec;32(4):407-16.

Tungu PK, Michael E, Sudi W, Kisinza WW, Rowland M. Efficacy of interceptor® G2, a long-lasting insecticide mixture net treated with chlorfenapyr and alpha-cypermethrin against *Anopheles funestus*: experimental hut trials in north-eastern Tanzania. Malar J. 2021 Apr 9;20(1):180.

Von Seidlein L, Olaosebikan R, Hendriksen IC, Lee SJ, Adedoyin OT, Agbenyega T, Nguah SB, Bojang K, Deen JL, Evans J, Fanello CI. Predicting the clinical outcome of severe falciparum malaria in african children: findings from a large randomized trial. Clinical infectious diseases. 2012 Apr 15;54(8):1080-90.

Wilson AL, IPTc Taskforce. A systematic review and meta-analysis of the efficacy and safety of intermittent preventive treatment of malaria in children (IPTc). PLoS one. 2011 Feb 14;6(2):e16976.

WHO Malaria Threat Map – Vector Insecticide Resistance Map. World Health Organization. 2024. https://apps.who.int/malaria/maps/threats/.

WorldPop (www.worldpop.org - School of Geography and Environmental Science, University of Southampton; Department of Geography and Geosciences, University of Louisville; Departement de Geographie, Universite de Namur) and Center for International Earth Science Information Network (CIESIN), Columbia University (2018). Global High Resolution Population Denominators Project - Funded by The Bill and Melinda Gates Foundation (OPP1134076). https://dx.doi.org/10.5258/SOTON/WP00645

Yeboah D, Owusu-Marfo J, Agyeman YN. Predictors of malaria vaccine uptake among children 6–24 months in the Kassena Nankana Municipality in the Upper East Region of Ghana. Malaria Journal. 2022 Nov 16;21(1):339.

Zongo I, Milligan P, Compaore YD, Some AF, Greenwood B, Tarning J, Rosenthal PJ, Sutherland C, Nosten F, Ouedraogo JB. Randomized noninferiority trial of dihydroartemisinin-piperaquine compared with sulfadoxine-pyrimethamine plus amodiaquine for seasonal malaria chemoprevention in Burkina Faso. Antimicrobial agents and chemotherapy. 2015 Aug;59(8):4387-96.
